# Transdiagnostic alterations in white matter microstructure associated with suicidal thoughts and behaviours in the ENIGMA Suicidal Thoughts and Behaviours consortium

**DOI:** 10.1101/2024.11.07.24316876

**Authors:** Laura S. van Velzen, Lejla Colic, Zuriel Ceja, Maria R. Dauvermann, Luca M. Villa, Hannah S. Savage, Yara J. Toenders, Niousha Dehestani, Alyssa H. Zhu, Adrian I. Campos, Lauren E. Salminen, Ingrid Agartz, Nina Alexander, Rosa Ayesa-Arriola, Elizabeth D. Ballard, Nerisa Banaj, Carlotta Barkhau, Zeynep Başgöze, Jochen Bauer, Francesco Benedetti, Klaus Berger, Bianca Besteher, Katharina Brosch, Manuel Canal-Rivero, Simon Cervenka, Romain Colle, Colm G. Connolly, Emmanuelle Corruble, Philippe Courtet, Baptiste Couvy-Duchesne, Benedicto Crespo-Facorro, Kathryn R Cullen, Udo Dannlowski, Jeremy Deverdun, Ana M. Diaz-Zuluaga, Lorielle M.F. Dietze, Jennifer W Evans, Negar Fani, Kira Flinkenflügel, Naomi P. Friedman, Ian H. Gotlib, Nynke A. Groenewold, Dominik Grotegerd, Tomas Hajek, Alexander S. Hatoum, Marco Hermesdorf, Ian B. Hickie, Yoshiyuki Hirano, Tiffany C. Ho, Yuki Ikemizu, Frank Iorfino, Jonathan C. Ipser, Yuko Isobe, Andrea P. Jackowski, Fabrice Jollant, Tilo Kircher, Melissa Klug, Sheri-Michelle Koopowitz, Anna Kraus, Axel Krug, Emmanuelle Le Bars, Elisabeth J. Leehr, Meng Li, Elizabeth T.C. Lippard, Carlos Lopez-Jaramillo, Ivan I. Maximov, Andrew M. McIntosh, Katie A. McLaughlin, Sean R. McWhinney, Susanne Meinert, Elisa Melloni, Philip B. Mitchell, Benson Mwangi, Igor Nenadić, Stener Nerland, Emilie Olie, Victor Ortiz-García de la Foz, Pedro M. Pan, Fabricio Pereira, Fabrizio Piras, Federica Piras, Sara Poletti, Andrew E. Reineberg, Gloria Roberts, Rafael Romero-García, Matthew D. Sacchet, Giovanni A. Salum, Anca-Larisa Sandu, Carl M. Sellgren, Eiji Shimizu, Harry R. Smolker, Jair C. Soares, Gianfranco Spalletta, J. Douglas Steele, Frederike Stein, Dan J. Stein, Benjamin Straube, Lea Teutenberg, Florian Thomas-Odenthal, Paula Usemann, Romain Valabregue, Johanna Valencia-Echeverry, Gerd Wagner, Gordon Waiter, Martin Walter, Heather C. Whalley, Mon-Ju Wu, Tony T. Yang, Carlos A. Zarate, Andre Zugman, Giovana B. Zunta-Soares, Kees van Heeringen, Sanne J.H. van Rooij, Nic van der Wee, Steven van der Werff, Paul M. Thompson, Hilary P. Blumberg, Anne-Laura van Harmelen, Miguel E. Rentería, Neda Jahanshad, ENIGMA Suicidal Thoughts and Behaviours Consortium, Lianne Schmaal

## Abstract

Previous studies have suggested that alterations in white matter (WM) microstructure are implicated in suicidal thoughts and behaviours (STBs). However, findings of diffusion tensor imaging (DTI) studies have been inconsistent. In this large-scale mega-analysis conducted by the ENIGMA Suicidal Thoughts and Behaviours (ENIGMA-STB) consortium, we examined WM alterations associated with STBs.

Data processing was standardised across sites, and resulting WM microstructure measures (fractional anisotropy, axial diffusivity, mean diffusivity and radial diffusivity) for 25 WM tracts were pooled across 40 cohorts. We compared these measures among individuals with a psychiatric diagnosis and lifetime history of suicide attempt (*n*=652; mean age=35.4±14.7; female=71.8%), individuals with a psychiatric diagnosis but no STB (i.e., clinical controls; *n*=1871; mean age=34±14.8; female=59.8%), and individuals with no mental disorder diagnosis and no STB (i.e., healthy controls; *n*=642; mean age=29.6±13.1; female=62.9%). We also compared these measures among individuals with recent suicidal ideation (*n*=714; mean age=36.3±15.3; female=66.1%), clinical controls (*n*=1184; mean age=36.8±15.6; female=63.1%), and healthy controls (*n*=1240; mean age= 31.6±15.5; female=61.0%).

We found subtle but statistically significant effects, such as lower fractional anisotropy associated with a history of suicide attempt, over and above the effect of psychiatric diagnoses. These effects were strongest in the corona radiata, thalamic radiation, fornix/stria terminalis, corpus callosum and superior longitudinal fasciculus. Effect sizes were small (Cohen’s *d* < 0.25). Recent suicidal ideation was not associated with alterations in WM microstructure.

This large-scale coordinated mega-analysis revealed subtle regional and global alterations in WM microstructure in individuals with a history of suicide attempt. Longitudinal studies are needed to confirm whether these alterations are a risk factor for suicidal behaviour.

## Introduction

Suicide is a worldwide public health concern, with more than 700,000 deaths by suicide occurring worldwide annually [1]. For each death by suicide, it is estimated that there are more than 20 attempts, which has a cascading effect that negatively impacts families, friends, and communities [1]. Despite widespread international efforts to reduce deaths by suicide, the number of deaths by suicide continues to increase in several regions of the world [2]. Risk and protective factors for suicidal thoughts and behaviours (STBs) have been identified [3]. Our current understanding of the underlying pathophysiological mechanisms is limited. Therefore, it is crucial that we increase our knowledge and understanding of these neurobiological mechanisms so that we can better target prevention and intervention efforts for individuals with high suicide risk.

Many prior magnetic resonance imaging (MRI) studies have sought to identify neural correlates of suicidal ideation and suicide attempt [for a review, please see 4, 5]. However, identifying robust and reliable patterns of neural alterations associated with STBs has been hampered by methodological heterogeneity across studies. Furthermore, due to the high clinical heterogeneity, associations with brain alterations are likely subtle; hence, large samples are needed to increase statistical power and identify neural correlates of STBs. The ENIGMA Suicidal Thoughts and Behaviours (ENIGMA-STB) consortium was established to address these issues by pooling data across international research groups to identify neural correlates of STBs. In our first study of subcortical and cortical grey matter morphology and STBs in young people between 8 and 25 years of age, we identified a subtle association between lifetime history of suicide attempt and the surface area of the frontal pole, a region in the prefrontal cortex [6], in a well-phenotyped subsample that was enriched for STBs and more severe symptoms of major depressive disorder (MDD) or bipolar disorder (BD). In addition to grey matter morphology, alterations in the microstructure of white matter (WM) tracts connecting brain regions may also contribute to risk for STBs. Identifying these WM alterations could reveal new treatment targets and increase the accuracy of monitoring or treatment response prediction.

Previous neuroimaging studies have used diffusion tensor imaging (DTI) to examine microstructural alterations related to STBs [for a review, please see 7]. These studies have focused primarily on fractional anisotropy (FA), which measures the coordinated directionality of water diffusion in WM fibre tracts and may reflect the coherence and myelination of neuronal fibres [8, 9]. A lifetime history of suicide attempt has been associated with lower FA values in various regions and WM tracts, including in the prefrontal cortex (PFC) [10–13], corpus callosum [14–16], cingulum [11], internal capsule [11, 17], uncinate fasciculus [16, 18], and inferior fronto-occipital fasciculus [16]. However, other studies found higher FA in individuals with a history of suicide attempt [19, 20]. Fewer studies have examined associations between FA measures and suicidal ideation, but findings from these studies suggest that suicidal ideation is associated with lower FA in the corpus callosum [21, 22], uncinate fasciculus [23], and corona radiata [21].

Thus, few studies have examined WM microstructural alterations related to suicidal ideation, and findings on suicide attempt are inconsistent across studies. Prior studies are hampered by small sample sizes [4], which decreases the likelihood of identifying true effects, increases the probability of false-negative findings, and may cause inflation of effect sizes [24]. In addition, large samples may be needed to identify subtle associations between STBs and WM microstructure. Finally, most prior studies focused on FA and did not examine other WM diffusivity measures, including axial diffusivity (AD), which is associated with axonal number and organisation; mean diffusivity (MD), which may be an estimate of membrane density; and radial diffusivity (RD), which can provide insights into myelination [25, 26]. Previous studies have also focused only on the presence or absence of suicidal ideation or suicide attempt, and have not examined specific aspects of suicide-related thoughts, such as the severity of suicidal ideation.

To address these limitations, we pooled data from 40 cohorts from the ENIGMA-STB consortium to examine associations between measures of WM microstructure (FA, AD, MD and RD) for 25 WM tracts and STBs in a large transdiagnostic sample. We examined WM microstructure alterations in people with a psychiatric diagnosis and lifetime history of suicide attempt compared to individuals with a diagnosis but no history of suicide attempt (i.e., clinical controls; CLC) and individuals with no disorder and no history of suicide attempt (healthy controls). In addition, we examined WM alterations in people with recent suicidal ideation (within the last six months) but no history of suicide attempt, compared to CLC and HC.

Based on previous findings concerning suicidal behaviour, we expected that a lifetime history of suicide attempt would be associated with lower FA in WM tracts that connect the frontal lobe with limbic regions or connect different limbic regions, such as the cingulum, corpus callosum, internal capsule, and uncinate fasciculus [11, 14, 17, 18, 23].

In a subsample of participants for whom more in-depth assessment of STBs from the Columbia Suicide Severity Rating Scale (CSSRS) was available, we investigated associations between WM microstructure and the severity of suicidal ideation. Finally, in this sample, we were able to examine associations between WM microstructure and suicide attempt, and also distinguish among interrupted, aborted, and actual suicide attempt.

## Patients and methods

### Cohorts

We pooled data from 40 international cohorts from 15 countries (see Supplementary Figure 1) to investigate the association between STBs and WM microstructure in a transdiagnostic sample, including individuals diagnosed with major depressive disorder, obsessive-compulsive disorder, bipolar disorder, post-traumatic stress disorder, psychotic disorders, generalised anxiety disorder, panic disorder, or social anxiety disorder. Demographic characteristics of the different samples are presented in **Table 1** and **Table 2**. The inclusion and exclusion criteria for the different studies are shown in **Supplemental Table S1**. All cohorts obtained ethics approval from their local institutional review boards and ethics committees. Participants who were 18 years old and over provided written informed consent; those under the age of 18 years provided written informed assent in addition to written informed consent from a parent/guardian at the local institution.

**Table 1.** Descriptive statistics for sites included in the lifetime suicide attempt analyses. Presented here are age (mean and standard deviation) and sex for the three groups (HC: healthy controls, CLC: clinical controls, group with lifetime suicide attempt) for the different sites included in the analysis on lifetime history of suicide attempt.

| Site | Age<br>HC | Age<br>CLC | Age<br>suicide<br>attempt<br>group | %<br>female<br>HC | %<br>female<br>CLC | %<br>female<br>suicide<br>attempt<br>group | Total<br>N HC | Total<br>N<br>CLC | Total N<br>suicide<br>attempt<br>group |
| --- | --- | --- | --- | --- | --- | --- | --- | --- | --- |
| BiDirect | - | 48.6<br>(7.3) | 49.5<br>(7.3) | - | 56.3 | 66.9 | 0 | 316 | 124 |
| CHU<br>Montpellier<br>BICS | 42.2<br>(11.7) | 45.3<br>(14.4) | 44.2<br>(13.8) | 71.4 | 86.4 | 90.0 | 21 | 22 | 20 |
| CHU<br>Montpellier<br>IMPACT | - | 38.3<br>(12.9) | 38.6<br>(9.8) | - | 94.4 | 85.7 | 0 | 18 | 7 |
| CHU<br>Montpellier<br>UF8555 | 37.3<br>(7.9) | 35.4<br>(8.4) | 37.9<br>(9.5) | 100.0 | 100.0 | 100.0 | 28 | 47 | 42 |
| DCHS | 30.9<br>(5.6) | 29.7<br>(6.4) | 32.3<br>(6.0) | 100.0 | 100.0 | 100.0 | 22 | 20 | 9 |
| MOODS team<br>DEP-ARREST<br>CLIN | 35.5<br>(13.0) | 33.2<br>(10.2) | 34.8<br>(14.3) | 67.7 | 70.8 | 63.0 | 31 | 24 | 27 |
| ETPB-STB | 33.9<br>(11.3) | 34.3<br>(7.9) | 34.8<br>(11.9) | 65.0 | 50.0 | 63.6 | 20 | 12 | 11 |
| Fondazione<br>Santa Lucia _<br>Neuropsychiatr<br>y Lab /<br>(FSL_NPL)-<br>OCD sample | 39.3<br>(10.5) | 39.9<br>(11.1) | 38.1<br>(7.6) | 35.4 | 34.9 | 36.4 | 82 | 43 | 11 |
| FSL_NPL -<br>Schizophrenia<br>sample | - | 40.7<br>(11.7) | 43.9<br>(9.9) | - | 32.7 | 44.4 | 0 | 49 | 18 |
| Ghent | 33.6<br>(13.4) | 38.2<br>(11.7) | 34.8<br>(11.9) | 94.1 | 53.3 | 69.2 | 17 | 15 | 13 |
| GIPSI BD | - | 31.5<br>(10.3) | 34.4<br>(11.3) | - | 25.0 | 25.0 | 0 | 24 | 8 |
| GIPSI SZ | - | 39.7<br>(11.9) | 40.0<br>(11.4) | - | 62.2 | 68.0 | 0 | 45 | 25 |
| Grady Trauma<br>Project Emory<br>University | 39.4<br>(13.0) | 39.3<br>(12.1) | 36.1<br>(10.9) | 97.8 | 100.0 | 100.0 | 46 | 51 | 14 |
| Halifax | 40.4<br>(13.7) | 42.8<br>(17.8) | 51.0<br>(16.5) | 64.3 | 41.2 | 57.1 | 42 | 17 | 7 |
| Houston BD | 24.1<br>(14.7) | 29.1<br>(16.4) | 30.2<br>(11.2) | 43.1 | 48.4 | 64.3 | 51 | 64 | 14 |
| LTS Colorado<br>site 1 | 28.7<br>(1.1) | 28.6<br>(0.9) | 28.5<br>(0.5) | 63.6 | 37.0 | 76.9 | 55 | 92 | 13 |
| LTS Colorado<br>site 2 | 28.6<br>(0.6) | 28.7<br>(0.8) | 28.6<br>(0.5) | 73.3 | 62.5 | 66.7 | 30 | 72 | 9 |
| McGill<br>University | 35.8<br>(8.0) | 44.5<br>(11.3) | 39.4<br>(10.6) | 53.3 | 50.0 | 77.8 | 15 | 14 | 9 |
| MR-IMPACT | - | 15.0<br>(1.4) | 15.2<br>(1.2) | - | 67.1 | 91.4 | 0 | 70 | 35 |
| PAFIP | - | 29.8<br>(8.5) | 27.3<br>(7.8) | - | 36.4 | 26.7 | 0 | 77 | 15 |
| San Raffaele<br>Hospital | - | 47.7<br>(10.9) | 46.7<br>(10.2) | - | 63.5 | 74.7 | 0 | 288 | 79 |
| Sydney Bipolar<br>Kids and<br>Siblings | 20.9<br>(3.6) | 23.0<br>(4.8) | 24.1<br>(4.0) | 52.2 | 65.5 | 75.0 | 67 | 84 | 24 |
| Sydney Brain<br>and Mind<br>Centre | - | 19.5<br>(3.5) | 18.1<br>(2.6) | - | 65.1 | 68.8 | 0 | 169 | 32 |
| UCSF<br>Adolescent<br>MDD | 15.4<br>(1.4) | 15.7<br>(1.3) | 16.0<br>(1.1) | 60.0 | 55.8 | 71.4 | 40 | 43 | 14 |
| University of<br>Minnesota<br>Adolescent<br>MDD | 16.5<br>(2.2) | 15.5<br>(1.8) | 16.9<br>(2.1) | 72.7 | 78.9 | 71.4 | 11 | 38 | 7 |
| University of<br>Texas- Austin -<br>Bipolar Seed<br>Program | 20.5<br>(1.7) | 21.0<br>(2.3) | 22.4<br>(2.0) | 68.4 | 73.3 | 57.1 | 19 | 15 | 7 |
| University of<br>Washington/<br>Harvard | 12.0<br>(1.8) | 12.4<br>(2.6) | 14.4<br>(1.8) | 46.7 | 35.5 | 60.0 | 45 | 31 | 5 |
| Yale School of<br>Medicine | - | 19.3<br>(3.4) | 19.0<br>(3.2) | - | 60.4 | 73.6 | 0 | 111 | 53 |
| **Total** | **29.6**<br>**(13.1)** | **34.0**<br>**(14.8)** | **35.4**<br>**(14.7)** | **62.9** | **59.8** | **71.8** | **642** | **1871** | **652** |

**Table 2.**
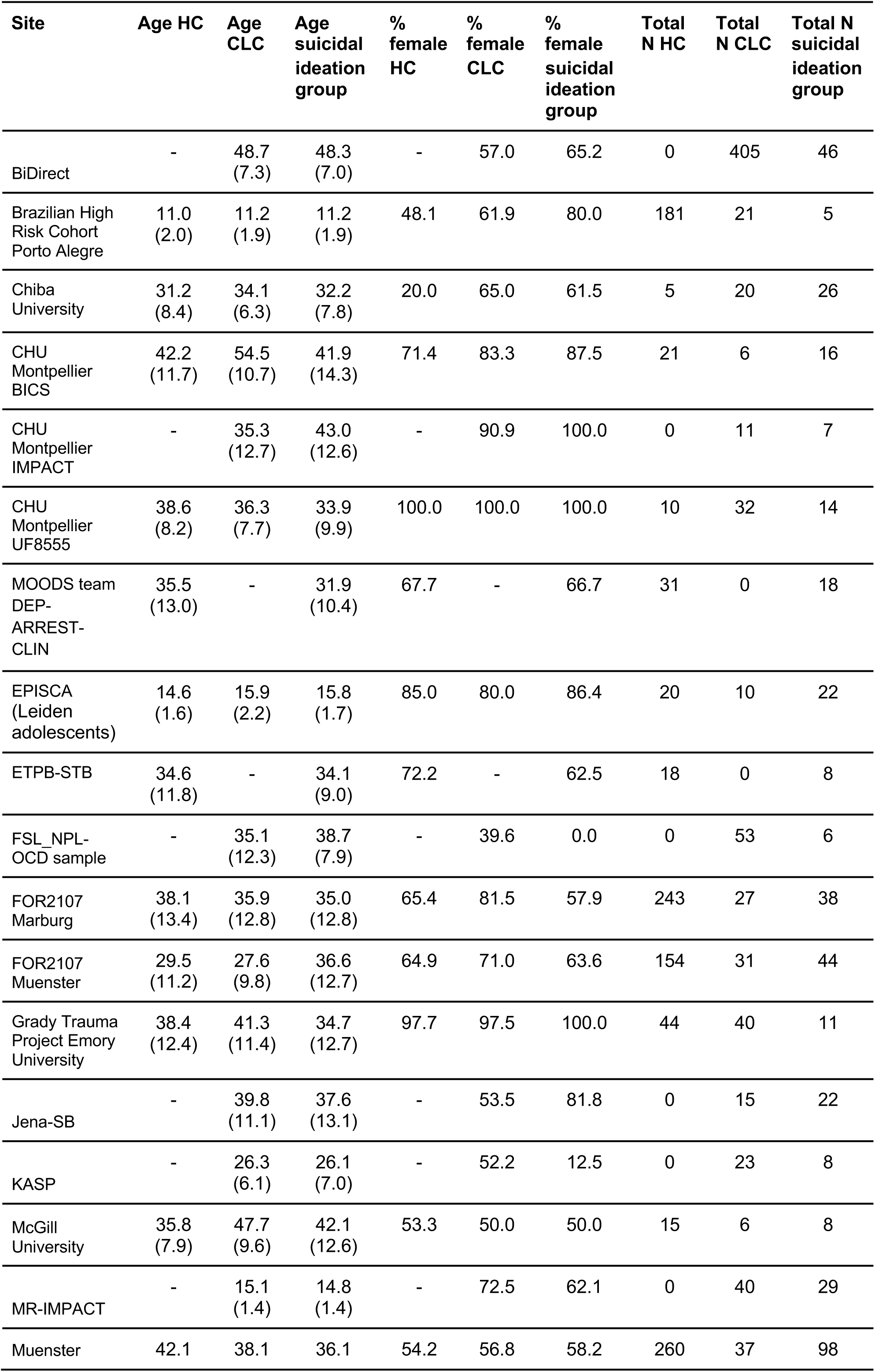
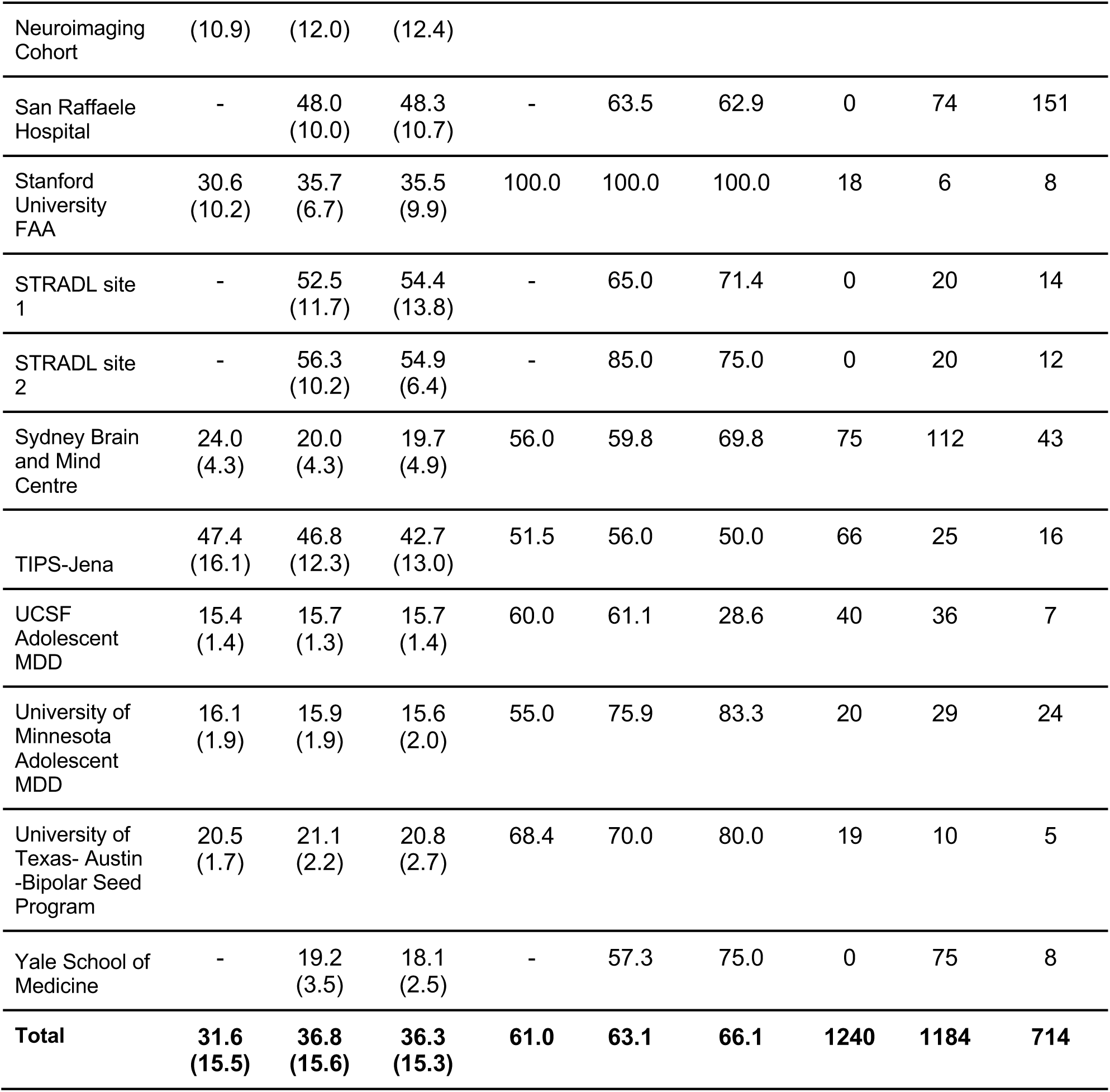
Descriptive statistics for studies included in the recent suicidal ideation analysis. Presented here are age (mean and standard deviation) and sex for the three groups (HC: healthy controls, CLC: clinical controls, group with recent suicidal ideation) for the different sites included in the analysis on suicidal ideation.

### Image processing

Scanner characteristics and acquisition parameters for all cohorts are provided in Supplemental **Table S2**. Each site performed local preprocessing of diffusion-weighted images, including diffusion tensor fitting. The pre-processed images were then processed using the ENIGMA-DTI protocol, including quality control procedures. This protocol is freely available on the ENIGMA-GitHub webpage (https://github.com/ENIGMA-git#enigma-dti-imaging) and NITRC (http://www.nitrc.org/projects/enigma_dti). For 24 tracts of interest, FA, AD, MD and RD measures were extracted (please see Supplemental **Table S3**). We combined regions of interest (ROIs) across hemispheres by calculating the mean of the left and right hemispheres, weighted by the number of voxels, in order to reduce the number of statistical tests. In addition, a global anisotropy or diffusivity measure was created, leading to a total of 25 measures for FA, AD, MD, and RD.

The WM microstructure measures were harmonised across sites using the ComBat algorithm in R [27, 28]. An empirical Bayes approach was used to adjust for variability between scanners while preserving biological variability related to age, sex, and diagnosis. In line with our previous study [6], all DTI measures included in the analyses were ComBat-corrected. Finally, within-site outliers (measures greater than three standard deviations away from the mean of that region) were visually inspected and, if necessary, excluded from the analysis.

### Statistical analysis

The main aim of this study was to examine associations among WM microstructure, lifetime history of suicide attempt, and recent (in the last six months) suicidal ideation in a large transdiagnostic sample. The cohorts included in this multi-study analysis administered different instruments to assess recent suicidal ideation and lifetime history of attempt. We used a similar approach to our previous study [6] to harmonise these measures across studies (see Supplemental **Table S4**). Lifetime history of suicide attempt (coded yes/no) was determined using clinical interviews (e.g., the Kiddie Schedule for Affective Disorders and Schizophrenia (K-SADS; [29]) or Structured Clinical Interview for DSM-5 (SCID; [30])) or detailed clinical scales on STBs (e.g., the CSSRS [31] or Self-Injurious Thoughts and Behaviours Interview (SITBI; [32])). Recent suicidal ideation (yes/no in the past six months or more recent) was assessed using items from depression severity rating scales (e.g. the Hamilton Depression Rating Scale (HDRS; [33]), Beck Depression Inventory (BDI; [34, 35]), detailed clinical scales on STBs (e.g., CSSRS or Beck Scale for Suicidal Ideation (SSI; [36, 37]) and clinical interviews (e.g. SCID). Similar to our previous study [6], we conducted separate analyses for suicidal ideation and suicide attempt to optimise the sample size for each analysis, as only 16 out of the 40 cohorts had information on both suicidal ideation and suicide attempt. In total, we included data from 28 cohorts in the analysis on lifetime suicide attempt and from 28 cohorts in the analysis on recent suicidal ideation.

We assessed differences among groups in global and regional anisotropy or diffusivity measures using multiple linear regression models in R [38]. We included a group variable to compare individuals with a lifetime history of suicide attempt (*N*=652) to either CLC (*N*=1871) or HC (*N*=642) (two group comparisons). A group variable was also included to compare individuals with recent suicidal ideation (*N*=714) to CLC (*N*=1184) and HC (*N*=1240) in separate analyses. In supplementary analyses, we also compared the HC and CLC groups (please see Supplemental **note 1** for a description of the methods and results). Consistent with previous ENIGMA-DTI analyses [39], the analyses were corrected for age, sex, and their linear and non-linear interactions (age-by-sex interaction, age^2^ and age^2^-by-sex interaction). Effect sizes were assessed using the Cohen’s *d* metric. All p-values were corrected for multiple testing (for the 25 tracts per anisotropic/diffusivity measure) using the Benjamini-Hochberg correction in R, resulting in FDR<.05.

Finally, in supplementary analyses, we examined interactions between group and type of lifetime psychiatric diagnosis in a subsample of participants, as data on lifetime psychiatric diagnosis were not available for all participants (please see Supplemental **note 2** for a description of the methods and results).

#### Analysis in the CSSRS sample

We also examined associations between WM microstructure and more detailed phenotypes of suicide attempt and suicidal ideation in a subsample of 7 cohorts that used the CSSRS to assess STBs. The CSSRS was specifically developed to assess the intensity and severity of suicidal thoughts and suicidal behaviour [31]. C-SSRS has good validity, high sensitivity and specificity for suicide attempts [40].

For analyses of suicidal ideation, we examined how the nominal measures of recent or lifetime severity of suicidal ideation (coded 0-5; 0: no ideation; 1: passive ideation; 2: non-specific active ideation; 3: active ideation with a method, but no plan or intent; 4: active ideation with intent, but no plan; 5: active ideation with a plan and intent) were associated with WM microstructure. For these analyses, the standardised beta was calculated as an effect size.

In line with our previous study, for analyses of suicide attempt, we compared anisotropy/diffusivity measures between individuals with a lifetime history of any attempt (actual, aborted, or interrupted attempt) and individuals with no lifetime history of any attempt [6]. In addition, we compared these measures between individuals with a lifetime history of an actual (non-interrupted and non-aborted) attempt and those without any lifetime suicide attempt. Finally, these measures were compared between individuals with a history of suicidal ideation (but no history of an actual suicide attempt), and those with a lifetime history of an actual attempt. Cohen’s *d* metric was calculated as an effect size for these analyses.

Similar to the main analyses, all analyses in the CSSRS sample included age, sex, age-by-sex, age^2^, and age^2^-by-sex as covariates, and all resulting p-values were corrected for multiple testing (for the 25 tracts per anisotropic/diffusivity measure) using the Benjamini-Hochberg correction in R to lead to FDR<.05.

## Results

### Lifetime suicide attempt

#### Individuals with a lifetime history of suicide attempt versus HC

Global FA and regional FA were lower in the attempt group (*N*=652) compared to HC (*N*=642) in the following tracts: anterior limb of the internal capsule (ALIC), body of the corpus callosum (BCC), corpus callosum (CC), cingulate gyrus of the cingulum bundle (CGC), corona radiata (CR), external capsule (EC), fornix/*stria terminalis* (FXST), genu of the corpus callosum (GCC), inferior fronto- occipital fasciculus (IFO), posterior *corona radiata* (PCR), posterior thalamic radiation (PTR), retrolenticular part of the internal capsule (RLIC), splenium of the corpus callosum (SCC), superior *corona radiata* (SCR), superior longitudinal fasciculus (SLF), sagittal stratum (SS), and uncinate fasciculus (UNC) (Cohen’s *d* range: 0.125-0.226; **Figure 1.A** and Supplemental **table S6**). In addition, MD was higher in the UNC in the attempt group compared to HC (Cohen’s *d*=0.220; Supplemental **table S7**). Finally, RD was higher in the BCC, CC, CR, FXST, PCR, SCC, SLF, and UNC in the attempt group compared to HC (Cohen’s *d* range: 0.148-0.170; **Figure 1.A** and Supplemental **table S8**). There were no significant differences in AD between the attempt group and HC (Supplemental **table S5**).

**Figure 1.**
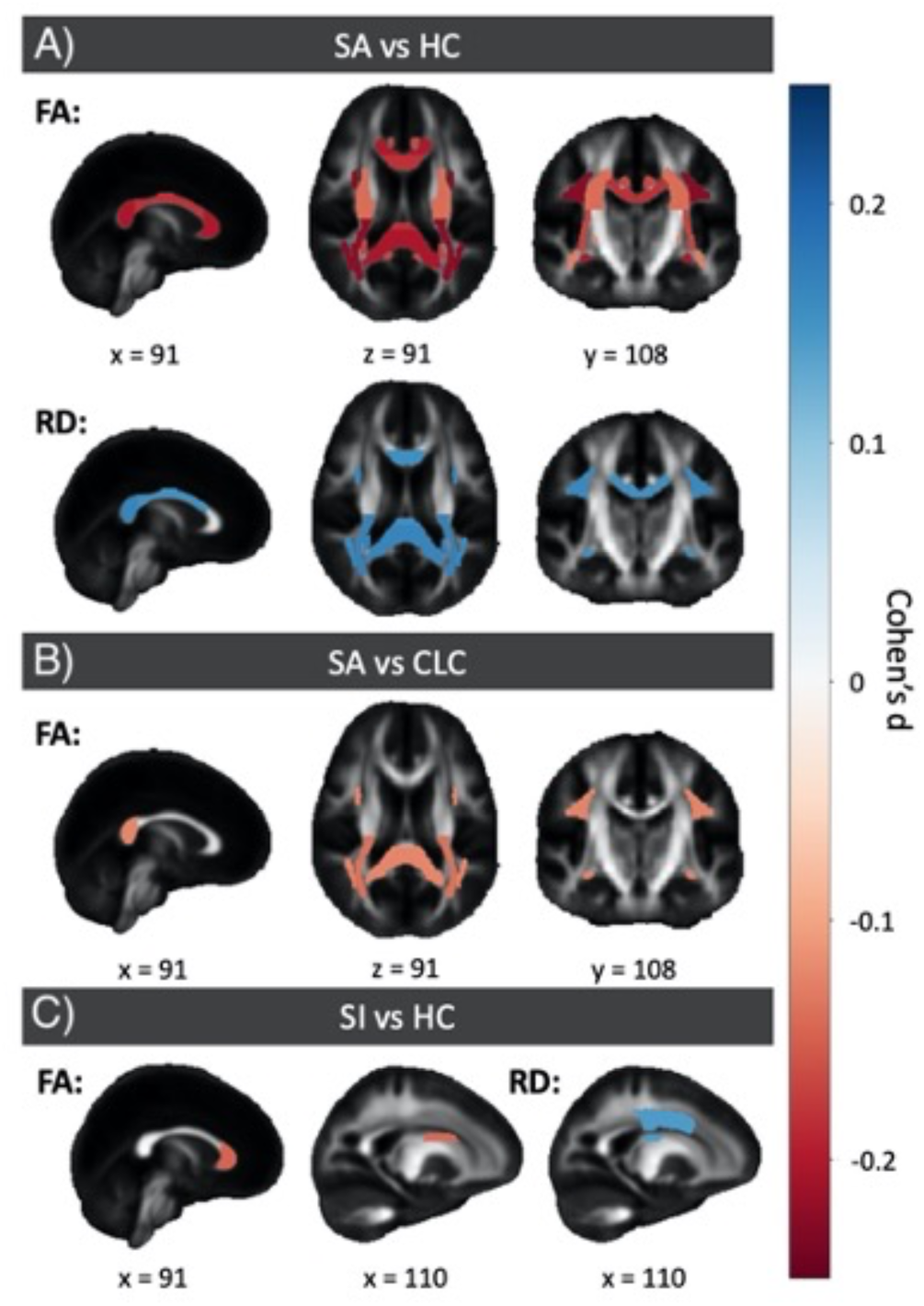
Cohen’s *d* map represents significant ROIs without the global covariate and threshold of pFDR < 0.05. From left to right, middle slices of sagittal, axial, and coronal views were used, except for C, where only the sagittal view was used. The figure includes three sections: A) FA and RD for individuals with a lifetime history of suicide attempt versus HC, B) FA for individuals with a lifetime history of suicide attempt versus CLC and C) FA and RD for individuals with recent suicidal ideation versus HC. <u>Abbreviations</u>: CLC= clinical control group ; FA= fractional anisotropy; HC= healthy control group; pFDRR= p value corrected using Benjamini Hochberg correction; RD= radial diffusivity; ROI= region of interest; SA= individuals with a lifetime history of suicide attempt; SI= individuals with recent suicidal ideation.

#### Individuals with a lifetime history of suicide attempt versus CLC

Global and regional AD, MD, and RD did not differ between individuals with a lifetime history of suicide attempt (*N*=652) and CLC (*N*=1871) (**Figure 1.B**, Supplemental **tables S9**, **S11,** and **S12**).

Regional FA was lower in the CR, FXST, PCR, PTR, SCC and SLF in the suicide attempt group compared to CLC (Cohen’s *d* range: 0.123-0.138; Supplemental **table S10**).

### Recent suicidal ideation

#### Individuals with recent suicidal ideation versus HC

Global and regional AD and MD did not differ between the suicidal ideation group (*N*=714) and HC (*N*=1240) (Supplemental **tables S13 and S15**). Global FA and regional FA in the CR, GCC and superior fronto-occipital fasciculus (SFO) were lower in the suicidal ideation group compared to HC (Cohen’s *d* range: 0.133-0.158; **Figure 1.C**, Supplemental **table S14**). Global RD and regional RD in the SCR were higher in the suicidal ideation group compared to HC (Cohen’s *d*: 0.142-0.150; Supplemental **table S16**).

#### Individuals with recent suicidal ideation versus CLC

There were no significant differences between the suicidal ideation group (*N*=714) and CLC (*N*=1184) (Supplemental **tables S17-20**).

### Deeper phenotyping in the C-SSRS sample

There were no significant associations of regional FA, AD, MD, RD, with severity of lifetime suicidal ideation (*N*=299; Supplemental **tables S21-24**) or severity of recent suicidal ideation (*N*=338; Supplemental **tables S25-28**). In addition, there were no significant differences in WM microstructure between people with a lifetime history of an actual suicide attempt (*N*=134) and those without a history of any suicide attempt (no interrupted, aborted, or actual suicide attempt; *N*=244) (Supplemental **tables 29-32**). Moreover, there were no significant differences between people with an actual suicide attempt (*N*=134) and those with lifetime suicidal ideation, but without a history of an actual suicide attempt (*N*=122; Supplemental **tables 33-36**). Finally, there were no differences in FA or diffusivity measures between people with a lifetime history of an interrupted, aborted, or actual suicide attempt (*N*=177) compared to individuals without a lifetime history of any subtype of suicide attempt (*N*=244; Supplemental **tables S37-40**).

## Discussion

In this large-scale multi-cohort transdiagnostic analysis, we examined associations between global and regional measures of WM microstructure and STBs in a pooled sample from 40 international cohorts. A lifetime history of suicide attempt was associated with subtle differences in global and regional FA and diffusivity (higher regional MD and RD), and recent suicidal ideation was associated with lower global and regional FA and higher global and regional RD.

Individuals with a lifetime history of suicide attempts had lower regional FA than did clinical controls. This analysis was undertaken to ascertain that the observed differences are not merely attributable to microstructural variances associated with psychiatric disorders, but rather, are significantly influenced by lifetime suicide attempt. The WM tracts that showed this difference between individuals with a lifetime history of suicide attempt and CLC included the *corona radiata* (including the posterior *corona radiata*), part of the fornix (fornix/*stria terminalis*), thalamic radiation (specifically the posterior thalamic radiation), splenium of the corpus callosum, and the superior longitudinal fasciculus. The *corona radiata* and the thalamic radiation connect the cortex with subcortical brain structures, including the brainstem and thalamus. The *corona radiata* is part of the thalamic-cortical circuitry and has been associated with perceptual, motor, emotional and cognitive function, including behavioural regulation, and lower FA in the *corona radiata* has been associated with poorer executive functioning [41–44]. In addition, previous single-cohort studies have reported that lower FA in the corona radiata is associated with suicidal behaviour [16, 45], albeit inconsistently [20]. Moreover, lower FA in the *corona radiata* predicted suicidal behaviour on average two years later [46]. The (posterior) thalamic radiations also connect the thalamus with cortical regions, including the parietal and occipital lobes. These are important for cortical arousal and consciousness but may also be associated with cognitive control [47]. While lower FA in this tract has not been associated with suicidal behaviour before, FA in this tract was lower in individuals with BD [48], who are at higher risk of suicidal behaviour [49]. The splenium of the corpus callosum connects the occipital-parietal and temporal cortex from both hemispheres and has been associated with visuospatial functioning, reading, language processing, and consciousness [50]. Lower FA in the splenium of the corpus callosum has been associated with a higher number of suicide attempts in BD and MDD [14]. The fornix (including the fornix/*stria terminalis*) connects the amygdala to the hypothalamus and plays an important role in threat monitoring, regulation of the hypothalamic-pituitary-adrenal (HPA) axis and behavioural inhibition [51]. In addition, previous investigators have speculated that impaired structural connectivity in this tract may contribute to feelings of anhedonia [52], which may drive suicidality [53]. Finally, the superior longitudinal fasciculus is an association tract that connects the parietal and temporal cortex with the frontal cortex and may play a role in speech, spatial awareness, processing speed, and attention [54]. FA in this region has inconsistently been associated with suicidality (see [20, 55] and has been consistently linked to brooding and rumination [56], which are risk factors for suicidal behaviour [57].

In this large sample, we were unable to replicate previous findings of lower FA in other WM tracts, including the cingulum [11], internal capsule [11, 17], uncinate fasciculus [16, 18], and inferior fronto- occipital fasciculus [16]. This lack of replication may be related to the small sample size of these previous studies, related to sample characteristics, or be specific to certain psychiatric disorders.

We also found differences in global and regional RD between individuals with a history of suicide attempt and HC but not CLC, suggesting that these findings were driven by psychiatric illness and not by suicidal behaviour *per se*. Similarly, differences in FA and RD were identified between individuals with recent suicidal ideation and HC but not CLC, which may also be related to mental illness in general and not to suicidal ideation specifically. Consistent with this finding, similar effects for FA were observed comparing CLC to HC; however, these findings were weaker and most did not survive correction for multiple testing (see Supplemental **note 1**). We hypothesise that mental illness may be more severe in the suicidal ideation group than in the CLC group, which may explain why differences in FA were more pronounced.

Our findings provide evidence consistent with previous ENIGMA reports about differences in WM microstructure in major depressive disorder [39], bipolar disorder [58] and schizophrenia [59] with DTI data, where associations with global and regional FA were reported. The MDD study included 1305 individuals diagnosed with MDD and 1602 HC; the results showed lower global FA and regional FA in 16 out of 25 WM tracts and higher RD in adult individuals with recurrent MDD; the largest FA differences were observed in the corpus callosum and *corona radiata* [39]. On the other hand, the schizophrenia study included 1963 individuals diagnosed with schizophrenia and 2359 HC, and the results showed lower global FA and regional FA in 20 out of 25 ROIs; the larger differences were reported in anterior *corona radiata* and corpus callosum [59]. Finally, the study on bipolar disorder included 1482 individuals diagnosed with BD and 1551 HC and showed lower global FA and regional FA in 29 out of 43 tracts in patients, most prominently in the corpus callosum and cingulum [58].

In a subsample of 7 cohorts, which had more deeply phenotyped suicidal thoughts and behaviours using the CSSRS, we were able to examine the associations with more detailed phenotypes of suicide attempt and suicidal ideation. In our previous study [6] on grey matter morphology and STB in young people, lower surface area of the frontal pole was associated with a history of actual (non- interrupted or non-aborted) suicide attempt in a subsample of studies that had used the CSSRS to assess suicidal behaviour (4 out of 7 cohort overlap, 53% total sample overlap with the CSSRS sample in the current study). In this study, we did not find any associations between WM anisotropy or diffusivity measures and severity of suicidal ideation, nor did we find differences in these measures related to suicide attempt in general (actual, interrupted or aborted suicide attempt), or actual suicide attempt specifically (non-interrupted or non-aborted suicide attempts). It is possible that this sample was not large enough to identify subtle associations between suicide attempt and WM microstructure.

This study increases our understanding of the mechanisms underlying suicidal behaviour by confirming findings from previous smaller studies that lower FA is implicated in suicidal behaviour and showing which tracts are involved. However, the effect sizes for significant group differences observed in this study were small (all d<0.25), especially for differences between the suicidal ideation/suicide attempt groups and the CLC. High clinical heterogeneity may mask larger effects in individual persons, which we were not able to detect with the group average approach used in the current study. Nonetheless, the currently observed differences are too subtle to be clinically useful in terms of prediction of risk for suicidal behaviour at the individual level. In addition, it is unclear whether alterations in WM microstructure represent a risk factor for suicidal behaviour or are a consequence of a previous suicide attempt. Previous longitudinal studies have shown that lower regional FA at baseline predicts suicidal behaviour at follow-up in individuals with mood disorders [11, 46]. Further, FA has been found to be highly heritable [60], suggesting that alterations in WM microstructure may indeed represent a risk factor for suicidal behaviour. However, cellular brain damage following hypoxia/anoxia during a suicide attempt may also cause WM damage and affect WM microstructure [61].

A strength of this study is the large sample size, which allowed the examination of more detailed phenotypes in subsamples. A second strength is the use of harmonised protocols for diffusion image processing and quality control. However, we should also acknowledge several limitations of this study, including heterogeneity across samples in how STBs were assessed. We have tried to minimise this effect by using a detailed process to harmonise these measures across studies (in line with the approach used in our previous study). Finally, we could not control for severity of psychiatric symptoms, as this was a transdiagnostic analysis and the different cohorts had used different severity rating scales; therefore, we cannot rule out the possibility that higher symptom severity in the suicide attempt group affected our findings. The scope of this study was limited to cross-sectional findings. However, this presents an opportunity for future research to delve into longitudinal analyses, shedding light on the evolution of the differences reported.

To conclude, in this large-scale multi-cohort study we found an association between a history of suicide attempt and regional FA, above and beyond the effect of psychiatric diagnosis. FA in several tracts that have been associated with (among others) behavioural regulation/inhibition, executive functioning, and brooding/rumination show a subtle association with suicidal attempt. Future longitudinal studies are needed to examine if altered FA may represent a risk factor for suicidal behaviour, for instance by contributing to lower resilience to severely stressful life events.

## Data Availability

All data produced in the present study could be made available upon reasonable request to the authors, but access to raw data is limited, as approval from each participating cohort is required for this.

## Acknowledgments

This work was supported by the MQ Brighter Futures Award MQBFC/2 (LS, LC, MD, LvV, ALvH, HB) and the National Institute of Mental Health of the National Institutes of Health under Award Number R01MH117601 (LS, LvV, NJ). ALvH was funded by a MQ Brighter Futures Award MQBFC/2, a Royal Society Dorothy Hodgkin Fellowship (DH15017), an MRC MRF emerging leaders award, the Leiden University Social Resilience and Security fund, and an NWO VIDI award. HPB was supported by the MQ Brighter Futures Award MQBFC/2, the National Center for Advancing Translational Science (Grant Number: UL1TR000142), the National Institute of Mental Health (Grant Numbers: RC1MH088366, R01MH69747), Brain and Behavior Research Foundation, International Bipolar Disorders Foundation, American Foundation for Suicide Prevention SRG-1-10-119, and the John and Hope Furth Endowment. EDB is supported by the National Institute of Mental Health Intramural Research Program (ZIA MH002857). NB is supported by the Italian Ministry of Health, grant Ricerca Corrente RC 23. The University of Minnesota adolescent MDD study is supported by the National Institute of Mental Health (K23MH090421), the National Alliance for Research on Schizophrenia and Depression, the University of Minnesota Graduate School, the Minnesota Medical Foundation, and the Biotechnology Research Center (P41 RR008079 to the Center for Magnetic Resonance Research), University of Minnesota, and the Deborah E. Powell Center for Women’s Health Seed Grant, University of Minnesota. FB is supported by the Italian Ministry of Health, grant PNRR-MAD-2022-12375859. BiDirect was funded by grants from the German Federal Ministry of Education and Research (BMBF; Grants FKZ-01ER0816 and FKZ-01ER1506) to KB. BB and LC are supported by the Interdisciplinary Center of Clinical Research of the Medical Faculty Jena. M.C.-R. acknowledges funding support from the Consejería de Salud y Familias (Junta de Andalucía) 2020 grant, which covers his salary (RH-0081-2020). Funding for the DEPARRESTCLIN cohort was provided by a national grant (ANR SAMENTA 2012) of the Agence Nationale de la Recherche (ANR). BCD is funded by a NHMRC CJ Martin fellowship (1161356). BCF acknowledges this work was supported by the Instituto de Salud Carlos III (PI14/00639 and PI14/00918) and Fundación Instituto de Investigación Marqués de Valdecilla (NCT0235832 and NCT02534363). UD acknowledges this work was funded by the German Research Foundation (DFG), Udo Dannlowski (co- speaker FOR2107, DA 1151/5-1, DA 1151/5-2, grant DA1151/9-1, DA1151/10-1 and DA1151/11-1) and the Interdisciplinary Center for Clinical Research (IZKF) of the medical faculty of Münster (grant Dan3/022/22 to UD). MuensterNeuroimagingCohort: This work was funded by the German Research Foundation (SFB- TRR58, Project C09 to UD). JWE is supported by the National Institute of Mental Health Intramural Research Program (ZIA MH002857). NPF acknowledges the LTS Colorado data acquisition was supported by grants from the National Institutes of Health (NIH) in the United States (AG046938 and MH063207). NPF was supported by NIH grants AG046938,, MH117131, MH124846, DA042742, DA046413, DA046064, and DA051018. IHG is supported by the National Institute of Mental Health Grant R37MH101495. TH acknowledges this study was supported by funding from the Canadian Institutes of Health Research (103703, 106469 and 142255, 180449, 186254), Nova Scotia Health Research Foundation, Dalhousie Clinical Research Scholarship to T. Hajek, Brain & Behavior Research Foundation (formerly NARSAD); 2007 Young Investigator and 2015 Independent Investigator Awards to T. Hajek. This work was also supported by the AMED Brain/MINDS Beyond Program (JP18dm0307002) and JSPS KAKENHI (JP19K03309 and JP22H01090). TCH is supported in part by the National Institutes of Health (K01MH117442, R21MH130817, R01MH127176). YI was supported by JSPS KAKENHI (JP23K07004). FI was supported by an NHMRC EL1 Investigator Grant (GNT2018157) and the Bill and Patricia Richie Foundation Fellowship. YI was supported by JSPS KAKENHI (JP23K02956). FJ acknowledges this study was supported by an operating grant from the Canadian Institutes for Health Research (CIHR #119288). TK acknowledges this work was funded by the German Research Foundation (DFG grants FOR2107 KI588/14-1, and KI588/14-2, and KI588/20-1, KI588/22-1 to Tilo Kircher, Marburg, Germany). Biosamples and corresponding data were sampled, processed and stored in the Marburg Biobank CBBMR. This work was funded by the German Research Foundation (DFG grants KR 3822/5-1, KR 3822/7-2 to Axel Krug). ETCL s funded in part by the American Foundation for Suicide Prevention SRG-0-112-20. CLJ is supported by PRISMA U.T. AMM is supported by the Wellcome Trust Grants (220857/Z/20/Z, 226770/Z/22/Z, 104036/Z/14/Z, 216767/Z/19/Z), and the European Union Horizon 2020 Grant (Grant Agreement 847776). KAM acknowledges this study was funded by the National Institute of Mental Health (R01-MH103291). PBM acknowledges this study was funded by the Australian National Medical and Health Research Council (Program Grant 1037196 (PBM and MB); Investigator Grant 1177991 (PBM)), the Lansdowne Foundation, Good Talk, and the Keith Pettigrew Family Bequest (PBM). IN work is funded by the Deutsche Forschungsgemeinschaft (DFG) grants NE2254/1-2, NE2254/2-1, NE2254/3-1, NE2254/4-1. PMP acknowledges BHRCS was supported with grants from the National Institute of Development Psychiatric for Children and Adolescent (INPD) and the Grant Fapesp 2014/50917-0 - 2021/05332-8 CNPq 465550/2014-2. FP acknowledges support for the team in Nimes, CINES grants access to HPC facilities (A0100311413). FaP is supported by the Italian Ministry of Health, grant RF-2019-12370182. FeP is supported by the Italian Ministry of Health, grant Ricerca Corrente RC 23. SP is supported by the Italian Ministry of Health, grant PNRR-MAD-2022-12375716, RF-2018-12367249. GR acknowledges this study was funded by the Australian National Medical and Health Research Council (Program Grant 1037196 (PBM and MB); Investigator Grant 1177991 (PBM)), the Lansdowne Foundation, Good Talk, and the Keith Pettigrew Family Bequest (PBM). RRG was supported by EMERGIA Junta de Andalucía program (EMERGIA20_00139), the VII Plan Propio of the University of Seville, the Plan de Consolidación (CNS2023-143647) and the Proyectos de Generación de Conocimiento (PID2021-122853OA- I00) from the Spanish Ministry of Science and Innovation. MDS is supported by the National Institute of Mental Health (Project Number R01MH125850); Brain and Behavior Research Foundation (Grant Number 28972). HRS was supported by a grant from the National Institutes of Health in the United States (DA051018). JCS acknowledges this work was partially supported by NIMH (1R01MH085667-01A1), John S. Dunn Foundation (Houston, Texas), and Pat Rutherford Chair in Psychiatry (UTHealth Houston). GS is supported by the Italian Ministry of Health, grant Ricerca Corrente RC 23. JDS is supported by the grants MRC (MR/S010351/1, MR/W002388/1, MR/W002566/1) and EPSRC (EP/Y017544/1). The DCHS cohort is funded by the Bill & Melinda Gates Foundation [INV-006732]. DJS is funded by the South African Medical Research Council. BS acknowledges this work was funded by the German Research Foundation (DFG grant as part of FOR2107: STR1146/18-1 to Benjamin Straube, Marburg, Germany). MW is supported by German Center for Mental Health (DZPG); FKZ: 01EE2103; and NeuroMarKet: Neuroimaging and Blood Markers as Indicators of Ketamine Efficacy in Treatment Resistant Depression; BMBF-EU-EraNet-Neuron, FKZ: 01EW2010A. HCW acknowledges this work is supported by the Chief Scientist Office of the Scottish Government Health Directorates [CZD/16/6], Scottish Funding Council [HR03006], Wellcome Trust [216767/Z/19/Z] & Wellcome Trust (Wellcome Trust Strategic Award “STratifying Resilience and Depression Longitudinally” (STRADL) Reference 104036/Z/14/Z). CAZ is supported by the National Institute of Mental Health Intramural Research Program (ZIA MH002857). TTY is supported by the National Center for Complementary and Integrative Health (NCCIH) R21AT009173, R61AT009864, and R33AT009864 to TTY; by the National Center for Advancing Translational Sciences (CTSI), National Institutes of Health, through UCSF-CTSI UL1TR001872 to TTY; by the American Foundation for Suicide Prevention (AFSP) SRG-1-141- 18 to TTY; by UCSF Weill Institute for Neurosciences to TTY; by UCSF Research Evaluation and Allocation Committee (REAC) and J. Jacobson Fund to TTY; by the National Institute of Mental Health (NIMH) R01MH085734 and the Brain and Behavior Research Foundation (formerly NARSAD) to TTY. PMT is supported by NIH grants R01AG058854, R01MH116147 and R01MH129742. NJ is supported by R01MH134004. MER receives Fellowship support from the Rebecca L Cooper Medical Research Foundation (F20231230).

## ENIGMA Suicidal Thoughts and Behaviours Consortium

Laura S. van Velzen^1,2^, Lejla Colic^3,4,5^, Zuriel Ceja^6,7^, Maria R. Dauvermann^8,9^, Luca M. Villa^3^, Hannah S. Savage^2,10^, Yara J. Toenders^1,2,11^, Niousha Dehestani^10,12^, Alyssa H. Zhu^13^, Adrian I. Campos^14^, Lauren E. Salminen^13^, Ingrid Agartz^15,16,17^, Nina Alexander^18,50^, Rosa Ayesa-Arriola^19^, Elizabeth D. Ballard^20^, Nerisa Banaj^21^, Carlotta Barkhau^22^, Zeynep Başgöze^24^, Jochen Bauer^23,^ Francesco Benedetti^25^, Klaus Berger^26^, Bianca Besteher^4,5^, Katharina Brosch^18,50,86^, Manuel Canal- Rivero^27,28^, Simon Cervenka^17,29^, Romain Colle^30^, Colm G. Connolly^31^, Emmanuelle Corruble^30^, Philippe Courtet^32,33^, Baptiste Couvy-Duchesne^34,35^, Benedicto Crespo-Facorro^27,28,36^, Kathryn R Cullen^24^, Udo Dannlowski^22^, Jeremy Deverdun^37,38^, Ana M. Diaz-Zuluaga^87,88^, Lorielle M.F. Dietze^39^, Jennifer W Evans^20^, Negar Fani^40^, Kira Flinkenflügel^22^, Naomi P. Friedman^41^, Ian H. Gotlib^42^, Nynke A. Groenewold^43^, Dominik Grotegerd^22^, Tomas Hajek^39^, Alexander S. Hatoum^44^, Marco Hermesdorf^26^, Ian B. Hickie^89^, Yoshiyuki Hirano^90,91^, Tiffany C. Ho^92,93^, Yuki Ikemizu^90,94^, Frank Iorfino^89^, Jonathan C. Ipser^43^, Yuko Isobe^90,91^, Andrea P. Jackowski^45,46^, Fabrice Jollant^47,48,49,30^, Tilo Kircher^18,50^, Melissa Klug^22^, Sheri-Michelle Koopowitz^43^, Anna Kraus^22^, Axel Krug^18,51^, Emmanuelle Le Bars^37,38^, Elisabeth J. Leehr^22^, Meng Li^4,5^, Elizabeth T.C. Lippard^52,53^, Carlos Lopez-Jaramillo^88^, Ivan I. Maximov^15,54^, Andrew M. McIntosh^55^, Katie A. McLaughlin^56,57^, Sean R. McWhinney^39^, Susanne Meinert^22,58^, Elisa Melloni^25^, Philip B. Mitchell^59^, Benson Mwangi^60,95^, Igor Nenadić^18,50^, Stener Nerland^15,16^, Emilie Olie^32,33^, Victor Ortiz-García de la Foz^19^, Pedro M. Pan^46^, Fabricio Pereira^61,62^, Fabrizio Piras^21^, Federica Piras^21^, Sara Poletti^25^, Andrew E. Reineberg^63^, Gloria Roberts^59^, Rafael Romero-García^9,27^, Matthew D. Sacchet^64^, Giovanni A. Salum^65,66^, Anca-Larisa Sandu^67^, Carl M. Sellgren^17,68^, Eiji Shimizu^90,91,94^, Harry R. Smolker^69^, Jair C. Soares^60,95^, Gianfranco Spalletta^21,70^, J. Douglas Steele^71^, Frederike Stein^18,50^, Dan J. Stein^72^, Benjamin Straube^18,50^, Lea Teutenberg^18,50^, Florian Thomas-Odenthal^18,50^, Paula Usemann^18,50^, Romain Valabregue^73,74^, Johanna Valencia-Echeverry^88^, Gerd Wagner^4,75^, Gordon Waiter^67^, Martin Walter^4,5,76,77^, Heather C. Whalley^55,78^, Mon-Ju Wu^60,95^, Tony T. Yang^79^, Carlos A. Zarate^20^, Andre Zugman^80^, Giovana B. Zunta-Soares^60,95^, Kees van Heeringen^96^, Sanne J.H. van Rooij^40^, Nic van der Wee^81,82^, Steven van der Werff^81,82^, Paul M. Thompson^13^, Hilary P. Blumberg^3,83,84^, Anne-Laura van Harmelen^9,85^, Miguel E. Rentería^6,7^, Neda Jahanshad^13^, Lianne Schmaal^1,2^

## References

1. WHO. Suicide worldwide in 2019: global health estimates. 2021. 2021.

2. Martínez-Alés G, Jiang T, Keyes KM, Gradus JL. The Recent Rise of Suicide Mortality in the United States. Annu Rev Public Health. 2022;43:99–116.

3. Wasserman D, Carli V, Iosue M, Javed A, Herrman H. Suicide prevention in childhood and adolescence: a narrative review of current knowledge on risk and protective factors and effectiveness of interventions. Asia Pac Psychiatry. 2021;13:e12452.

4. Schmaal L, van Harmelen A-L, Chatzi V, Lippard ETC, Toenders YJ, Averill LA, et al. Imaging suicidal thoughts and behaviors: a comprehensive review of 2 decades of neuroimaging studies. Mol Psychiatry. 2020;25:408–427.

5. Auerbach RP, Pagliaccio D, Allison GO, Alqueza KL, Alonso MF. Neural Correlates Associated With Suicide and Nonsuicidal Self-injury in Youth. Biol Psychiatry. 2021;89:119– 133.

6. van Velzen LS, Dauvermann MR, Colic L, Villa LM, Savage HS, Toenders YJ, et al. Structural brain alterations associated with suicidal thoughts and behaviors in young people: results from 21 international studies from the ENIGMA Suicidal Thoughts and Behaviours consortium. Mol Psychiatry. 2022:1–11.

7. Zanghì E, Corallo F, Lo Buono V. Diffusion tensor imaging studies on subjects with suicidal thoughts and behaviors: A descriptive literature review. Brain Behav. 2022;12:e2711.

8. Podwalski P, Szczygieł K, Tyburski E, Sagan L, Misiak B, Samochowiec J. Magnetic resonance diffusion tensor imaging in psychiatry: a narrative review of its potential role in diagnosis. Pharmacol Rep. 2021;73:43–56.

9. Jones DK, Knösche TR, Turner R. White matter integrity, fiber count, and other fallacies: the do’s and don’ts of diffusion MRI. Neuroimage. 2013;73:239–254.

10. Olvet DM, Peruzzo D, Thapa-Chhetry B, Sublette ME, Sullivan GM, Oquendo MA, et al. A diffusion tensor imaging study of suicide attempters. J Psychiatr Res. 2014;51:60–67.

11. Lippard ETC, Cox Lippard ET, Johnston JAY, Spencer L, Quatrano S, Fan S, et al. Preliminary examination of gray and white matter structure and longitudinal structural changes in frontal systems associated with future suicide attempts in adolescents and young adults with mood disorders. Journal of Affective Disorders. 2019;245:1139–1148.

12. Mahon K, Burdick KE, Wu J, Ardekani BA, Szeszko PR. Relationship between suicidality and impulsivity in bipolar I disorder: a diffusion tensor imaging study. Bipolar Disord. 2012;14:80– 89.

13. Gosnell SN, Molfese DL, Salas R. Brain Morphometry: Suicide. In: Spalletta G, Piras F, Gili T, editors. Brain Morphometry, New York, NY: Springer New York; 2018. p. 403–427.

14. Cyprien F, de Champfleur NM, Deverdun J, Olié E, Le Bars E, Bonafé A, et al. Corpus callosum integrity is affected by mood disorders and also by the suicide attempt history: A diffusion tensor imaging study. J Affect Disord. 2016;206:115–124.

15. Lischke A, Domin M, Freyberger HJ, Grabe HJ, Mentel R, Bernheim D, et al. Structural Alterations in the Corpus Callosum Are Associated with Suicidal Behavior in Women with Borderline Personality Disorder. Front Hum Neurosci. 2017;11:196.

16. Wei S, Womer FY, Edmiston EK, Zhang R, Jiang X, Wu F, et al. Structural alterations associated with suicide attempts in major depressive disorder and bipolar disorder: A diffusion tensor imaging study. Prog Neuropsychopharmacol Biol Psychiatry. 2020;98:109827.

17. Jia Z, Huang X, Wu Q, Zhang T, Lui S, Zhang J, et al. High-field magnetic resonance imaging of suicidality in patients with major depressive disorder. Am J Psychiatry. 2010;167:1381– 1390.

18. Johnston JAY, Wang F, Liu J, Blond BN, Wallace A, Liu J, et al. Multimodal Neuroimaging of Frontolimbic Structure and Function Associated With Suicide Attempts in Adolescents and Young Adults With Bipolar Disorder. Am J Psychiatry. 2017;174:667–675.

19. Kim B, Oh J, Kim M-K, Lee S, Tae WS, Kim CM, et al. White matter alterations are associated with suicide attempt in patients with panic disorder. J Affect Disord. 2015;175:139–146.

20. Lee S-J, Kim B, Oh D, Kim M-K, Kim K-H, Bang SY, et al. White matter alterations associated with suicide in patients with schizophrenia or schizophreniform disorder. Psychiatry Res Neuroimaging. 2016;248:23–29.

21. Reis JV, Vieira R, Portugal-Nunes C, Coelho A, Magalhães R, Moreira P, et al. Suicidal Ideation Is Associated With Reduced Functional Connectivity and White Matter Integrity in Drug-Naïve Patients With Major Depression. Front Psychiatry. 2022;13:838111.

22. Zhang R, Jiang X, Chang M, Wei S, Tang Y, Wang F. White matter abnormalities of corpus callosum in patients with bipolar disorder and suicidal ideation. Ann Gen Psychiatry. 2019;18:20.

23. Fan S, Lippard ETC, Sankar A, Wallace A, Johnston JAY, Wang F, et al. Gray and white matter differences in adolescents and young adults with prior suicide attempts across bipolar and major depressive disorders. J Affect Disord. 2019;245:1089–1097.

24. Button KS, Ioannidis JPA, Mokrysz C, Nosek BA, Flint J, Robinson ESJ, et al. Power failure: why small sample size undermines the reliability of neuroscience. Nat Rev Neurosci. 2013;14:365–376.

25. Winklewski PJ, Sabisz A, Naumczyk P, Jodzio K, Szurowska E, Szarmach A. Understanding the Physiopathology Behind Axial and Radial Diffusivity Changes—What Do We Know? Front Neurol. 2018;9:92.

26. Tae WS, Ham BJ, Pyun SB, Kang SH, Kim BJ. Current Clinical Applications of Diffusion-Tensor Imaging in Neurological Disorders. J Clin Neurol. 2018;14:129–140.

27. Fortin J-P, Cullen N, Sheline YI, Taylor WD, Aselcioglu I, Cook PA, et al. Harmonization of cortical thickness measurements across scanners and sites. Neuroimage. 2018;167:104–120.

28. Radua J, Vieta E, Shinohara R, Kochunov P, Quidé Y, Green MJ, et al. Increased power by harmonizing structural MRI site differences with the ComBat batch adjustment method in ENIGMA. Neuroimage. 2020;218:116956.

29. Kaufman J, Birmaher B, Brent D, Rao U, Flynn C, Moreci P, et al. Schedule for Affective Disorders and Schizophrenia for School-Age Children-Present and Lifetime Version (K-SADS-PL): Initial Reliability and Validity Data. Journal of the American Academy of Child & Adolescent Psychiatry. 1997;36:980–988.

30. 30. First MB. Structured clinical interview for DSM-IV Axis I disorders SCID-I: Clinician version, scoresheet. American Psychiatric Press; 1997.

31. Posner K, Brent D, Lucas C, Gould M, Stanley B, Brown G, et al. Columbia-suicide severity rating scale (C-SSRS). New York, NY: Columbia University Medical Center. 2008. 2008.

32. Nock MK, Holmberg EB, Photos VI, Michel BD. Self-Injurious Thoughts and Behaviors Interview: development, reliability, and validity in an adolescent sample. Psychol Assess. 2007;19:309–317.

33. Hamilton M. A rating scale for depression. J Neurol Neurosurg Psychiatry. 1960;23:56–62.

34. Beck AT, Ward C, Mendelson M, Mock J, Erbaugh J. Beck depression inventory (BDI). Arch Gen Psychiatry. 1961;4:561–571.

35. Beck AT, Steer RA, Brown GK, Others. Beck depression inventory-II. San Antonio. 1996;78:490–498.

36. Beck AT, Kovacs M, Weissman A. Assessment of suicidal intention: the Scale for Suicide Ideation. J Consult Clin Psychol. 1979;47:343–352.

37. Beck AT, Steer RA, Ranieri WF. Scale for suicide ideation: Psychometric properties of a self-report version. J Clin Psychol. 1988;44:499–505.

38. 38. R core team. R: A language and environment for statistical computing. R Foundation for Statistical Computing, Vienna, Austria. Http://www.R-Project.Org/. 2013. 2013.

39. van Velzen LS, Kelly S, Isaev D, Aleman A, Aftanas LI, Bauer J, et al. White matter disturbances in major depressive disorder: a coordinated analysis across 20 international cohorts in the ENIGMA MDD working group. Mol Psychiatry. 2020;25:1511–1525.

40. Posner K, Brown GK, Stanley B, Brent DA, Yershova KV, Oquendo MA, et al. The Columbia-Suicide Severity Rating Scale: initial validity and internal consistency findings from three multisite studies with adolescents and adults. Am J Psychiatry. 2011;168:1266–1277.

41. Jenkins LM, Barba A, Campbell M, Lamar M, Shankman SA, Leow AD, et al. Shared white matter alterations across emotional disorders: A voxel-based meta-analysis of fractional anisotropy. Neuroimage Clin. 2016;12:1022–1034.

42. Chen Z, Cui L, Li M, Jiang L, Deng W, Ma X, et al. Voxel based morphometric and diffusion tensor imaging analysis in male bipolar patients with first-episode mania. Prog Neuropsychopharmacol Biol Psychiatry. 2012;36:231–238.

43. Leunissen I, Coxon JP, Caeyenberghs K, Michiels K, Sunaert S, Swinnen SP. Task switching in traumatic brain injury relates to cortico-subcortical integrity. Hum Brain Mapp. 2014;35:2459–2469.

44. Wallace EJ, Mathias JL, Ward L. The relationship between diffusion tensor imaging findings and cognitive outcomes following adult traumatic brain injury: A meta-analysis. Neurosci Biobehav Rev. 2018;92:93–103.

45. Jiang X, Guo Y, Jia L, Zhu Y, Sun Q, Kong L, et al. Altered Levels of Plasma Inflammatory Cytokines and White Matter Integrity in Bipolar Disorder Patients With Suicide Attempts. Front Psychiatry. 2022;13:861881.

46. Colic L, Villa LM, Dauvermann MR, van Velzen LS, Sankar A, Goldman DA, et al. Brain grey and white matter structural associations with future suicidal ideation and behaviors in adolescent and young adult females with mood disorders. JCPP Adv. 2022;2.

47. Chaddock-Heyman L, Erickson KI, Voss MW, Powers JP, Knecht AM, Pontifex MB, et al. White matter microstructure is associated with cognitive control in children. Biol Psychol. 2013;94:109–115.

48. Hu R, Stavish C, Leibenluft E, Linke JO. White Matter Microstructure in Individuals With and At Risk for Bipolar Disorder: Evidence for an Endophenotype From a Voxel-Based Meta-analysis. Biol Psychiatry Cogn Neurosci Neuroimaging. 2020;5:1104–1113.

49. Dome P, Rihmer Z, Gonda X. Suicide Risk in Bipolar Disorder: A Brief Review. Medicina. 2019;55.

50. Blaauw J, Meiners LC. The splenium of the corpus callosum: embryology, anatomy, function and imaging with pathophysiological hypothesis. Neuroradiology. 2020;62:563–585.

51. Clauss J. Extending the neurocircuitry of behavioural inhibition: a role for the bed nucleus of the stria terminalis in risk for anxiety disorders. Gen Psychiatr. 2019;32:e100137.

52. Harnett NG, Stevens JS, van Rooij SJH, Ely TD, Michopoulos V, Hudak L, et al. Multimodal structural neuroimaging markers of risk and recovery from posttrauma anhedonia: A prospective investigation. Depress Anxiety. 2021;38:79–88.

53. Bonanni L, Gualtieri F, Lester D, Falcone G, Nardella A, Fiorillo A, et al. Can Anhedonia Be Considered a Suicide Risk Factor? A Review of the Literature. Medicina. 2019;55:458.

54. Janelle F, Iorio-Morin C, D’amour S, Fortin D. Superior Longitudinal Fasciculus: A Review of the Anatomical Descriptions With Functional Correlates. Front Neurol. 2022;13:794618.

55. Davey DK, Jurick SM, Crocker LD, Hoffman SN, Sanderson-Cimino M, Tate DF, et al. White matter integrity, suicidal ideation, and cognitive dysfunction in combat-exposed Iraq and Afghanistan Veterans. Psychiatry Res Neuroimaging. 2021;317:111389.

56. Pisner DA, Shumake J, Beevers CG, Schnyer DM. The superior longitudinal fasciculus and its functional triple-network mechanisms in brooding. Neuroimage Clin. 2019;24:101935.

57. Horwitz AG, Czyz EK, Berona J, King CA. Rumination, Brooding, and Reflection: Prospective Associations with Suicide Ideation and Suicide Attempts. Suicide Life Threat Behav. 2019;49:1085–1093.

58. Favre P, Pauling M, Stout J, Hozer F, Sarrazin S, Abé C, et al. Widespread white matter microstructural abnormalities in bipolar disorder: evidence from mega- and meta-analyses across 3033 individuals. Neuropsychopharmacology. 2019;44:2285–2293.

59. Kelly S, Jahanshad N, Zalesky A, Kochunov P, Agartz I, Alloza C, et al. Widespread white matter microstructural differences in schizophrenia across 4322 individuals: results from the ENIGMA Schizophrenia DTI Working Group. Mol Psychiatry. 2018;23:1261–1269.

60. Kochunov P, Jahanshad N, Marcus D, Winkler A, Sprooten E, Nichols TE, et al. Heritability of fractional anisotropy in human white matter: a comparison of Human Connectome Project and ENIGMA-DTI data. Neuroimage. 2015;111:300–311.

61. DiPoce J, Guelfguat M, DiPoce J. Radiologic findings in cases of attempted suicide and other self-injurious behavior. Radiographics. 2012;32:2005–2024.

62. Correia MM, Carpenter TA, Williams GB. Looking for the optimal DTI acquisition scheme given a maximum scan time: are more b-values a waste of time? Magn Reson Imaging. 2009;27:163–175.

