## Supplement for "Transdiagnostic alterations in white matter microstructure associated with suicidal thoughts and behaviours in the ENIGMA Suicidal Thoughts and Behaviours consortium"

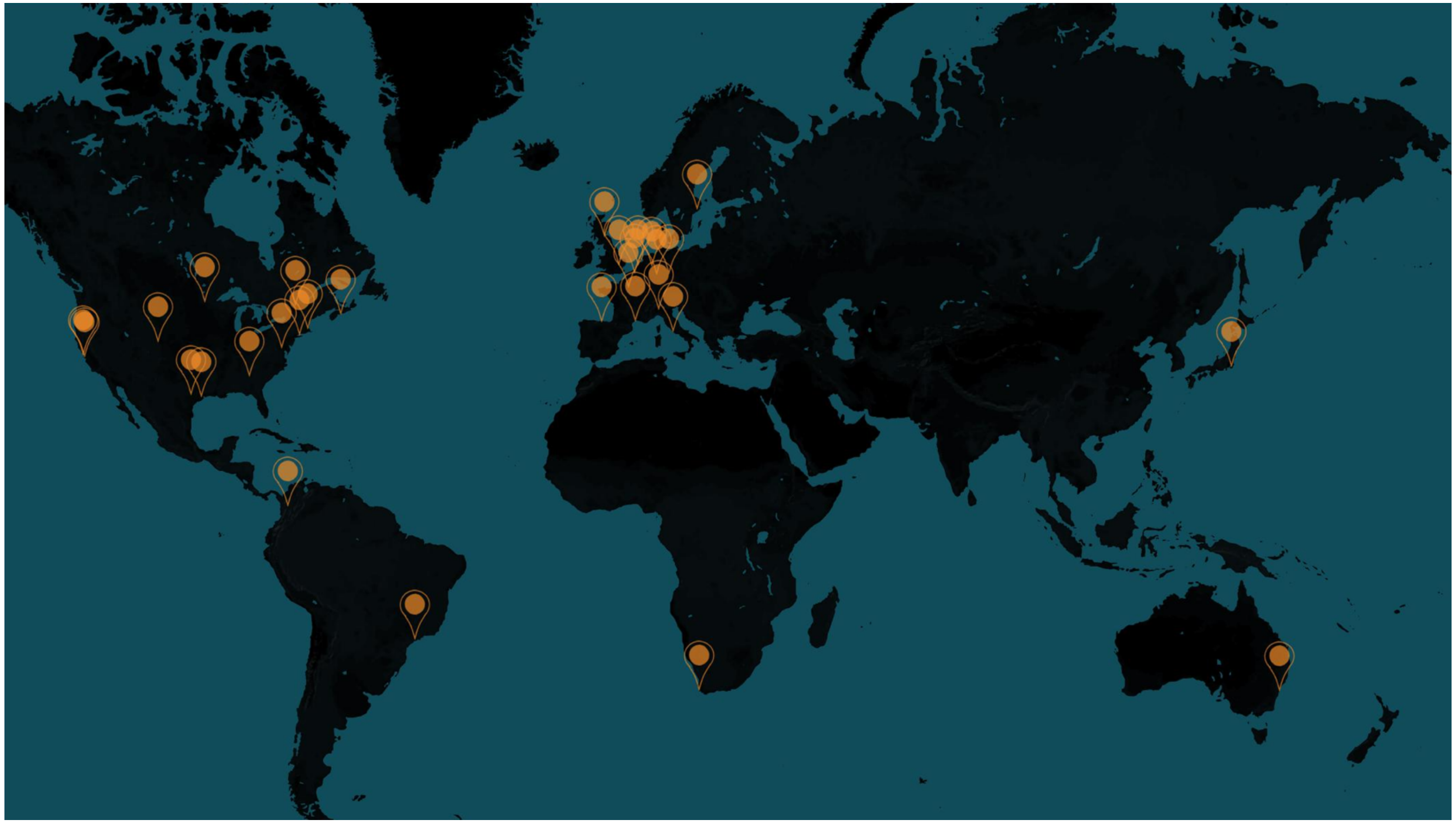

Supplemental Figure 1. Overview of ENIGMA Suicidal Thoughts and Behaviours consortium cohorts included in the analysis.

Table S1. Main diagnosis of participants and study exclusion criteria in the different sites

| Site | Main diagnosis participants | Inclusion criteria | Exclusion criteria |
|---|---|---|---|
| BiDirect | MDD | For patients: The recruitment of outpatients was limited to those who had been hospitalised due to depression at least once during the 12 months period prior to inclusion into the study. Inclusion criteria were (i) age (≥35 and < 66 years) and (ii) current in- or outpatient treatment due to acute depression. For healthy controls: The participants (≥35 and < 66 years) had been randomly sampled from the population register of the city of Münster. | Exclusion criteria for both patients and controls were (i) compulsory ad- mission, (ii) comorbid dementia, and (iii) comorbid drug abuse (including alcohol). |
| Brazilian High Risk Cohort Porto Alegre City | Various | Children and adolescents 6 to 14 years of age at baseline were attending one of the 57 public schools in two Brazilian cities (São Paulo and Porto Alegre). From the total cohort (N = 2511), 1004 children were invited for MRI scanning, and the images from 741 were obtained at cohort baseline. | Contraindications to MRI assessments (metal, braces, etc.) |
| Chiba University | OCD | For patients: OCD patients who are willing to receive CBT | For patients: Suicide attempt, Antisocial Personality Disorder; For healthy controls: History of a psychiatric disorder |
| CHU Montpellier BICS | MDD | For patients : right-handed, Caucasian adults aged between 18 and 70 years. current depressive episode according to the DSM-5 criteria. For healthy controls: right-handed, Caucasian adults aged between 18 and 70 years. No history of psychiatric disorders according to the DSM-5 criteria . No lifetime history of suicide attempt | Current neurologic or inflammatory disease; CRP >10mg/L; current antibiotic or anti-inflammatory treatment; history of head trauma with loss of consciousness; current substance misuse, excluding tobacco; lifetime history of manic or hypomanic episode, schizophrenia, or schizoaffective disorder according to the DSM-5; pregnancy and breastfeeding; contraindication to MRI. |
| CHU Montpellier IMPACT | MDD | Aged between 18 and 65 years, having a history of suicide attempt within the last 12 months | Lifetime history of schizophrenia, a current alcohol/ illicit drug use disorder, a current manic or hypomanic episode, a lifetime history of severe brain injury or neurologic disease and pregnancy;contraindication to MRI. |
| CHU Montpellier UF8555 | MDD/BD | Healthy controls : Caucasian right-handed females,>18 yo, no lifetime history of psychiatric disorder | Lifetime history of severe head trauma, CNS disorder, schizophrenia, and a history of alcohol or drug abuse or |

| | | | |
|---|---|---|---|
| | | Patients : Caucasian right-handed females, euthymic ( Hamilton Depression Rating Scale score <7 and Young Mania Rating Scale score < 7, history of major past depressive episode ; >18 yo | dependence within the past 12 months, pregnancy and breastfeeding ; contraindication to MRI. |
| DCHS | MDD/PTSD | For patients: enrolled in DCHS study; mothers between ages of 18 and 50; could have PTSD, MDD, or both;<br>For healthy controls: enrolled in DCHS study; mothers between ages of 18 and 50; no psychopathology | For patients: no psychopathology; brain injury or loss of consciousness for longer than 30 minutes; MRI exclusion criteria;<br>For healthy controls: brain injury or loss of consciousness for longer than 30 minutes; MRI exclusion criteria |
| MOODS team DEP-ARREST-CLIN | MDD | For patients: 18-65 years, current MDD diagnosis, minimum HDRS scores of 18, antidepressant-free at least one month prior to study inclusion<br>For healthy controls: absence of current or past psychiatric disorders or somatic conditions (including nasal polyposis and acute sinusitis or rhinitis) | For patients: diagnosis of bipolar disorder, psychotic disorder, eating disorder or addiction according to the DSM-V<br>For all participants: pregnancy or breastfeeding, nasal polyposis, acute or chronic sinusitis or rhinitis. |
| EPISCA (Leiden adolescents) | MDD, PTSD | For patients: aged between 12 and 19, estimated full scale IQ>80, right handed, normal or corrected to normal vision, sufficient understanding of the Dutch language, no history of neurological impairments and no contraindications for MRI.Having a depressive or anxiety disorder as classified by DSM-IV criteria (including PTSD), no current or prior use of antidepressants.<br>For healthy controls:<br>aged between 12 and 19, estimated full scale IQ>80, right handed, normal or corrected to normal vision, sufficient understanding of the Dutch language, no history of neurological impairments and no contraindications for MRI. No current or passed DSM-IV classifications, no clinical scores on validated mood and anxiety questionnaires, no history of traumatic experiences, and no current psychotherapeutic and/or psychopharmacological intervention of any kind | For both patients and healthy controls: primary DSM-IV clinical diagnosis of ODD, CD, pervasive developmental disorder, Tourette's syndrome, OCD, bipolar disorder, and psychotic disorder, current substance use, history of neurological disorders or severe head injury, and pregnancy |
| ETPB-STB | MDD | For patients: Eligible participants included men and women, ages 18–65 years. Subjects with MDD had been diagnosed with recurrent MDD without psychotic features using the Structured Clinical Interview for Axis I DSM-IV Disorders (SCID)-Patient Version [40]. Subjects were required to have a score ≥20 on the Montgomery-Åsberg Depression Rating | For patients: psychotic features;<br>For healthy controls: family history of Axis I disorders in first degree relatives |

| | | | |
|---|---|---|---|
| | | Scale (MADRS). Subjects had to have not responded to at least one adequate antidepressant trial during their current episode, as assessed using the Anti- depressant Treatment History Form, and the current episode had to have lasted at least four weeks. Subjects were free from psychotropic medications in the two weeks before randomization (5 weeks for fluoxetine, 3 weeks for aripiprazole);<br>For healthy controls: Healthy control subjects consisted of males and females, 18–65 years old with no Axis I disorder as determined by SCID-NP, and no family history of Axis I disorders in first degree relatives. Healthy control subjects were free of medications affecting neuronal function or cerebral blood flow or metabolism. Subjects in both groups were in good physical health as determined by medical history, physical exam, blood labs, electrocardiogram, chest x-ray, urinalysis, and toxicology. | |
| FSL_NPL- OCD sample | OCD | For patients: DSM-V diagnosis<br>For all participants: age between 18-65 years | Exclusion criteria for all participants: history of alcohol or drug abuse within past 2 years, lifetime drug dependence, traumatic head injury with loss of consciousness, MRI contraindications.<br>For patients: past or present major medical illness or neurological disorder, mental retardation, brain abnormalities, vascular lesions.<br>For controls: current or lifetime DSM-V psychiatric or personality disorder according to SCID, first-degree relative with psychosis, mental retardation, minor or major DSM-V major neurocognitive disorder, any brain abnormalities or vascular lesions |
| FSL_NPL- SZ sample | Psychosis | For patients: DSM-V diagnosis<br>For all participants: age between 18-65 years | Exclusion criteria for all participants: history of alcohol or drug abuse within past 2 years, lifetime drug dependence, traumatic head injury with loss of consciousness, MRI contraindications.<br>For patients: past or present major medical illness or neurological disorder, mental retardation, brain abnormalities, vascular lesions.<br>For controls: current or lifetime DSM-V psychiatric or personality disorder according to SCID, first-degree relative with psychosis, |

| | | | |
|---|---|---|---|
| | | | mental retardation, minor or major DSM-V major neurocognitive disorder, any brain abnormalities or vascular lesions |
| FOR2107 Marburg | MDD | For patients: age 18-65 years; patients were diagnosed by SCID-Interview with major depressive disorder (currently depressed or remitted) or with bipolar disorder (currently depressed, (hypo)manic or remitted) For healthy controls: age 18-65 years | Exclusion criteria all: any MRI contraindications; any neurological abnormalities. Exclusion criteria controls: any current or former psychiatric disorder; Exclusion criteria patients: substance dependence or current benzodiazepine treatment (wash out of at least three half-lives before study participation)" |
| FOR2107 Muenster | MDD | For patients: age 18-65 years; patients were diagnosed by SCID-Interview with major depressive disorder (currently depressed or remitted) or with bipolar disorder (currently depressed, (hypo)manic or remitted) For healthy controls: age 18-65 year | Exclusion criteria all: any MRI contraindications; any neurological abnormalities. Exclusion criteria healthy controls: any current or former psychiatric disorder; Exclusion criteria patients: substance dependence or current benzodiazepine treatment (wash out of at least three half-lives before study participation) |
| Ghent | | For patients: euthymic outpatients with a history of suicide attempts (SAs) and euthymic outpatients without a history of suicide attempt (NA) were included. All patients had a history of depression. They were recruited in different centres in the Flemish part of Belgium; For healthy controls: Healthy controls (HC), without any history of depression or suicidal behavior, were included. | For patients and controls:The MINI was used to rule out lifetime bipolar disorder, lifetime psychotic disorder and alcohol abuse in the past 12 months, as these were exclusion criteria. Possible neurological disorders and current medication use were questioned in detail by the primary researcher. Subjects with a lifetime history of neurological disorders or using psychotropic medication at time of participation, except for selective serotonin reuptake inhibitors and serotonin–norepinephrine reuptake inhibitors, were excluded. |
| GIPSI BD | BD | Diagnosis of Bipolar Disorder I or II based on DSM-IV-TR criteria using the Diagnostic Interview for Genetics Studies (DIGS), age between 18 and 60 years, education level between 5 and 16 years | Personal history of neurological disorders, mental disability, autism, electroconvulsive therapy and/or traumatic brain injury |
| GIPSI SZ | Psychosis | Diagnosis of Schizophrenia based on DSM-IV-TR criteria using the Diagnostic Interview for Genetics Studies (DIGS), age between 18 and 60 years, education level between 5 and 16 years | Personal history of neurological disorders, intellectual disability, autism, electroconvulsive therapy and/or traumatic brain injury |
| Grady Trauma Project Emory University | PTSD | All: 18-65 years of age, English-speaking, endorsed at least 1 criterion A trauma. For PTSD: meet DSM-IV criteria for PTSD | All: Current psychotic symptoms or bipolar disorder, current substance or alcohol dependence, history of head trauma, taking |

| | | | |
|---|---|---|---|
| | | | any psychoactive medication, current illegal drug use (verified with urine drug screen within 24 hours of scan) |
| Halifax | BD | Inclusion criteria. The BD patients (both Li and non-Li groups) had to have: (i) a diagnosis of bipolar I or II disorder made by a psychiatrist using the SCID; (ii) at least 10 years of illness; (iii) a history of at least five episodes of illness (including manic, depressive, or mixed episodes); (iv) current Hamilton Depression Rating Scale, 17-item version (HAM-D-17) score < 7; (v) current Young Mania Rating Scale (YMRS) score < 5; (vi) current Clinical Global Impressions Scale–Bipolar (CGI-BP) score < 3; and (vii) a period of euthymia for at least four months prior to scanning, as aside from state- related factors, patients in acute episodes may present with additional difficult to control confounding variables, including recent medication change or substance abuse. The non-Li group had to have less than three months of lifetime Li exposure, more than 24 months prior to the scanning. The Li group had to have a current Li treatment lasting a minimum of 24 months. (Halifax) Diabetes Study: The subjects with BD were required to 1) have the diagnosis of bipolar I or II disorder made by a psychiatrist; and 2) be at least 18 years of age. Patients were excluded if they had 1) the diagnosis of organic mood disorder; 2) mood disorder not otherwise specified; or 3) more than one lifetime course of electroconvulsive therapy or electroconvulsive therapy within the last 6 months.Halifax High Risk Study: Families were identified through adult probands with BD, who had participated in 1) previous genetic and high-risk studies for the Halifax sample. Only the offspring from these families, not the probands, were a part of the MRI study. The offspring from BD parents were divided into two subgroups: 1) the Unaffected HR group, which consisted of 50 offspring with no lifetime history of psychiatric disorders. These individuals were at an increased risk for BD because they had one parent affected with a primary mood disorder. 2) The Affected Familial group, which consisted of 36 offspring who met criteria for a lifetime Axis I diagnosis of mood disorders (i.e., a personal history of at least one episode of depression, hypomania, or mania meeting full DSM-IV criteria). When available, we recruited more than one offspring per family. From this study, we provided data only from patients who had a personal history of bipolar disorder. | Exclusion criteria. Individuals from any of the three groups were excluded if they met any of the magnetic resonance imaging (MRI) exclusion criteria or had any serious medical illness (e.g., brain injury, Cushings disease, or conditions treated with corticosteroids). Individuals with BD were excluded if they had: (i) more than one lifetime course of electroconvulsive therapy (ECT) or ECT in the previous 12 months; (ii) comorbid psychiatric disorders, and ⁄ or personality disorder; (iii) active substance abuse in the previous 12 months; (iv) significant change in their medication in the previous three months; or (v) current psychotic features or acute suicidality. Individuals from the non-Li group were excluded if they had: (i) Li exposure < 2 years before the scanning; or (ii) lifetime Li exposure of more than three months. The neuropsychiatrically healthy individuals were excluded if they had a personal history of psychiatric disorders. The neuropsychiatrically healthy, euglycemic subjects were excluded if they had 1) a personal history of psychiatric disorders; or 2) T2DM. Subjects from any group were excluded if they 1) met any magnetic resonance imaging (MRI) exclusion criteria; 2) suffered from substance abuse in the last 12 months; had a history of 3) neurodegenerative disorders; or 4) cerebrovascular disease/stroke, as we were interested in the more subtle T2DM-related neuronal changes. |

| | | | |
|---|---|---|---|
| Houston BD | BD | Patients: Bipolar I or II disorder diagnosed by SCID-I, according to DSM-IV-R diagnostic criteria; Any current mood state; Any comorbid psychiatric disorder, except for current substance abuse or dependence (see exclusion criteria); Age 18-65 years old.<br>Controls: No lifetime diagnosis of a major psychiatric (mood, anxiety, psychotic, addictive) and neurological disorders according to DSM-IV-R; No family history of any axis I psychiatric disorder and/or hereditary neurological disorders in first-degree relatives; 18-65 years old. | Healthy controls reporting current or past Axis I disorders, suicidal history and a first-degree relative with any Axis I disorder were excluded. Participants with any endocrinological disease, head trauma, neurological disease, and family history of any hereditary neurological disorder or medical conditions such as hypertension, diabetes, active liver disease and kidney problems were excluded from this study. |
| Jena-SB | MDD | Patients and healthy controls aged between 18–63, right-handed. Patients must meet the criteria of a current major depressive episode according to SCID-I. SA was defined according to Silverman et al. 2007 as a “self-inflicted, potentially injurious behavior with a nonfatal outcome for which there is evidence (either explicit or implicit) of intent to die." Healthy control subjects must have no current or previous mental disorders, suicidal ideation, or suicide attempts. | Comprised a lifetime history of schizophrenia or bipolar disorder, a history of alcohol/substance abuse or dependence spanning the previous 6 months, a major general medical condition requiring ongoing pharmacological treatment, a lifetime history of severe head trauma or central nervous system disorder, and contraindication to MRI. |
| KASP | Psychosis | Patients who seek health care for psychotic symptoms for the first time were recruited at psychiatric emergency wards and in- or outpatient facilities at the psychiatric clinics located in Stockholm, Sweden. A baseline diagnosis is established based on a structured clinical interview of the Diagnostic and Statistical Manual of Mental Disorders IV, DSM-IV (Structured Clinical Interview for DSMIV-Axis I Disorders, SCID-I) at the time of inclusion. Healthy controls were recruited by advertisement and matched on age and sex. | Exclusion criteria for patients were an ongoing or previous prescription of an antipsychotic for more than 30 days, severe somatic and neurological diseases, current substance abuse (except nicotine use), or autism spectrum disorder. The Mini International Neuropsychiatric Interview (MINI), performed by either a resident or a specialist in psychiatry, was used to exclude previous or current psychiatric illness among the healthy controls. Further exclusion criteria were former or current use of illegal drugs, first-degree relatives with psychotic illness or bipolar disorder, as well as neurologic disease and/or severe somatic disease. |
| LTS Colorado | HC | Ongoing participation in the Colorado Lontitudinal Twin Study (LTS). | Contraindications to MRI scanning or health conditions resulting in an inability to provide informed consent or complete tasks. |
| McGill University | MDD | Patients: Aged between 25 to 55, right-handed. Current major depressive episode as part of MDD according to SCID-1 and HDRS>19.<br>CLC: no personal or first-degree family history of suicidal behavior.<br>HC: no personal or second-degree family history of | CLC:<br>Lifetime history of schizophrenia, bipolar disorder, and history of alcohol/drug abuse or dependence over the last 6 months; history of major general medical condition that requires current pharmacological treatment; no use of an accepted method of contraception if nonmenopausal woman; pregnancy; a lifetime |

| | | | |
|---|---|---|---|
| | | depressive disorders or suicidal behavior and no current psychotropic treatment | history of severe head trauma, or central nervous system disorder; counter-indication to MRI (ferromagnetic body implants, piercings, claustrophobia, inability to lie still during one hour, etc.). |
| MR-IMPACT | MDD | Patients: age 11-17, current DSM-IV unipolar MDD diagnosis determined using child and parent interviews using the KSADS-PL, score of 27 or higher on the MFQ | Exclusion criteria for all participants: alcohol dependence, drug dependence, pervasive developmental disorder or generalised learning problem, pregnancy or breastfeeding, current medication use that could adversely interaction with SSRIs, MRI contraindications, brain abnormalities, intolerance to the MRI environment |
| Muenster Neuroimaging Cohort | MDD | "Inclusion criteria: age 17-65 years;patients were diagnosed with major depressive disorder by SCID-Interview, currently depressed or remitted. | For patients: presence of bipolar disorder, schizoaffective disorders and schizophrenia; substance-related disorders or current benzodiazepine treatment (wash out of at least three half-lives before study participation), and former electroconvulsive therapy. For controls: any current or former psychiatric disorder. For patients & controls: any neurological abnormalities, MRI contra-indications |
| PAFIP | Psychosis | For patients: 15-50 years, DSM-IV diagnosis of schizophrenia, schizophreniform disorder, schizoaffective disorder, delusional disorder, brief reactive psychosis or psychosis not otherwise specified<br>For healthy controls: no history of mental illness treatment and no axis-I disorder | For patients: history of neurological disease, head injury, mental retardation or drug dependence.<br>For healthy controls: current or past psychiatric diagnosis, mental retardation, neurological or general medical illnesses. Psychosis in first-degree relatives |
| San Raffaele Hospital | BD/MDD | For patients: Age 18-65, a baseline Hamilton Depression Rating Scale (HDRS) score of 17 or higher;<br>For healthy controls: Age 18-65 | For patients: other diagnoses on Axis I; mental retardation on Axis II; pregnancy, history of epilepsy, major medical and neurological disorders; treatment with long-acting neuroleptic drugs in the last three months before admission; history of drug or alcohol dependency or abuse within the last six months and for part of the sample inflammation-related symptoms, including fever and infectious or inflammatory disease; uncontrolled systemic disease; uncontrolled metabolic disease or other significant uncontrolled somatic disorder known to affect mood; somatic medications known to affect mood or the immune system, such as corticosteroids, non-steroid anti-inflammatory drugs and statins<br>For healthy controls: mental retardation, pregnancy, major medical/neurological or psychiatric disorders. |

| | | | |
|---|---|---|---|
| Stanford University FAA | MDD | For patients: women aged 18–55 years currently experiencing a diagnosable depressive episode (established using DSM-IV-TR criteria assessed with the Structured Clinical Interview for DSM Axis I)<br>For healthy controls: women aged 18–55 years not meeting criteria for any past or current Axis I disorder (established using DSM-IV-TR criteria assessed with the Structured Clinical Interview for DSM Axis I) | For patients: Cardiovascular or neurological disorders, history of head injury, hyper- or hypothyroidism, lifetime diagnosis of bipolar disorder or psychosis; substance abuse/dependence in the past six months<br>For healthy controls: Cardiovascular or neurological disorders, history of head injury, hyper- or hypothyroidism, history of any psychiatric disorder or substance abuse/dependence |
| STRADL | MDD | For patients: MDD diagnosis<br>For healthy controls: No psychological disorders | For patients: neurological disorders<br>For healthy controls: psychological or neurological disorders |
| Sydney Bipolar Kids and Siblings | BD | For patients: Any DSM-IV diagnosis | For healthy controls: parent or sibling with bipolar I or II, recurrent MDD, schizoaffective disorder, schizophrenia, recurrent substance<br>abuse, psychiatric hospitalisation. Parent with a first degree relative who had a past mood disorder hospitalisation or history of psychosis |
| Sydney Brain and Mind Centre | MDD + OCD + BD + Psychosis + GAD + SAD | For patients: Age 12 - 25 and presented with an affective, psychotic or developmental/behavioural syndrome. | Exclusion criteria for all subjects included medical instability (as determined by a psychiatrist), history of neurological disease (e.g. tumour, head trauma, epilepsy), medical illness known to impact cognitive<br>and brain function (e.g. cancer), intellectual and/or developmental disability and insufficient English for neuropsychological assessment. All subjects were asked to abstain from drug or alcohol use for 48 hours prior to testing and informed about a drug screen protocol. |
| TIPS-Jena | MDD | For individuals with MDD: presence of a DSM-IV diagnosis using the German version of the Structured Clinical Interview for DSM-IV Axis I Disorders (SKID-I) that were confirmed by two psychiatrists.<br>For healthy controls: participants did not have current or history of a DSM-IV axis I psychiatric disorder, a first-degree relative with a known psychiatric disorder or use of psychotropic medication. | For both individuals with MDDand healthy controls: exclusion criteria were history of major neurological and untreated major general medical conditions, and IQ lower than 80. |
| UCSF Adolescent MDD | MDD | For patients: presence of a primary diagnosis of MDD as determined via Schedule for Affective Disorders and Schizophrenia for School-Age Children-Present and Lifetime Version (Kaufman et al., 1997). For healthy control | Exclusion criteria for all participants included: 1) use of pharmacotherapeutics for treating psychiatric conditions within the past 6 months, 2) misuse of drugs within two months prior to |

| | | | |
|---|---|---|---|
| | | participants absence of any psychiatric diagnosis as determined by the Diagnostic Interview Schedule for Children (Shaffer et al., 2000) and Diagnostic Predictive Scale (Lucas et al., 2001) instruments. | MRI scanning; 3) two or more alcoholic drinks per week within the previous month (as assessed by the Customary Drinking and Drug Use Record; CDDR) (Brown et al, 1998); 4) a full scale IQ score of less than 75 (as assessed by the Wechsler Abbreviated Scale of Intelligence; WASI) (Wechsler, 1999); 5) contraindications for MRI including ferromagnetic implants and claustrophobia; 6) pregnancy or the possibility of pregnancy; 7) left-handedness; 8) prepubertal status (as assessed as Tanner stages of 1 or 2) (Tanner, 1962); 9) inability to understand and comply with procedures; 10) neurological disorder (including meningitis, migraine, or HIV); 11) head trauma; 12) learning disability; 13) serious health problems; and 14) complicated or premature birth (i.e., birth before 33 weeks of gestation). The MDD group was subject to the additional exclusion criterion of a primary psychiatric diagnosis other than MDD. The healthy control group was subject to the additional exclusion criteria of: 1) history of mood or psychotic disorders in a first- or second-degree relative (as assessed by the Family Interview for Genetics; FIGS) (Maxwell, 1992); and 2) current or lifetime DSM-IV-TR Axis I psychiatric disorder. |
| University of Minnesota | MDD | For patients: primary diagnosis of MDD and had not received any psychotropic medication treatment for the past 2 months. HC participants were eligible if they had no current or past psychiatric diagnoses. | Exclusion criteria for included the presence of a neurologic or other chronic medical condition, mental retardation, pervasive developmental disorder, substance use disorder, bipolar disorder, or schizophrenia |
| University of Texas - Austin - Bipolar Seed Program | BD | For patients: Meeting diagnostic criteria for bipolar disorder type I per the Structured Clinical Interview for DSM-5-Research Version, between 18-25 years of age | For patients: History of major medical illness or significant head trauma resulting in possible neurological or central nervous system damage, Past or current severe alcohol or other substance use disorder, Full scale IQ <85, Positive pregnancy test, Having a medical condition/previous surgery preventing participation in a magnetic resonance imaging scan<br>Not taking medications for greater than or equal to 4 weeks (i.e., participants must be stable on their medications) |

| | | | |
|---|---|---|---|
| | | | For healthy controls: Meeting criteria for psychotic, affective or other anxiety disorders per the Structured Clinical Interview for DSM-5-Research Version, History of major medical illness or significant head trauma resulting in possible neurological or central nervous system damage, Past or current severe alcohol or other substance use disorder, Full scale IQ <85, Positive pregnancy test, Having a medical condition/previous surgery preventing participation in a magnetic resonance imaging scan |
| University of Washington/ Harvard | MDD + PTSD + GAD + Panic disorder + Social Anxiety Disorder | For all participants: 8-20 years of age, English speaking, with and without exposure to trauma. For patients: Endorsement of ideation or behaviour on the SITBI | Exclusion criteria for all participants: Psychiatric medication use (stimulants for ADHD were discontinued for the scan), MRI contraindications, active substance dependence, pervasive developmental disorder. For healthy controls: Endorsement of ideation or behaviour on the SITBI |
| Yale School of Medicine | BD, MDD | For individuals with MDD and BD: ages 13-25 years and meeting criteria for major depressive disorder (MDD) or bipolar disorder (BD) type 1 or type 2 according to the DSM-IV. | Exclusion criteria for all individuals: history of or current medical or neurological condition that could affect the brain (except treated hypothyroidism), and magnetic resonance imaging (MRI) contraindications. |

Table S2. Scanner type and DWI acquisition parameters per site

| Site | Scanner vendor and type | Voxel size and slice thickness | Number of directions & b-factor | Preprocessing software & steps |
|---|---|---|---|---|
| BiDirect | 3T Philips Intera | 0.94 x 0.94 x 3.6 mm (reconstructed) | 20 directions; b-factor=1000 | Eddy correction, DTIfit in FSL |
| Brazilian High Risk Cohort Porto Allegre | GE Signa HD 1.5-T And GE Signa HDx 1.5T | 3 x 3 x 3 mm | 15 directions; b-factor=800 s/mm2 | Eddy current correction and DTIfit in FSL |
| Chiba University | 3T GE Discovery MR750 | 1.875 x 1.875 x 2 mm | 30 directions; b-factor=1000 | Eddy correction, DTIfit in FSL |
| CHU Montpellier BICS | 3T Siemens Magnetom SKYRA | 0.6 x 0.6 x 2 mm | 64 directions; b-factor=1000 | Denoising, distortion correction (SynB0), TOPUP, Eddy, and DTIfit in FSL |
| CHU Montpellier IMPACT | 3T Siemens Magnetom SKYRA | 2.5 x 2.5 x 2.5 mm | 30 directions; b-factor=1000 | Denoising, distortion correction (SynB0), TOPUP, Eddy, and DTIfit in FSL |
| CHU Montpellier UF8555 | 1.5T Siemens Magneton Avanto | 2.5 x 2.5 x 2.5 mm | 64 directions, b-factor=1000 | Denoising, distortion correction (SynB0), TOPUP, Eddy, and DTIfit in FSL |
| DCHS | 3T Siemens Magnetom Allegra Sygno | 2 x 2 x 2 mm | 45 directions; b-factor=1000 | Eddy, TOPUP, DTIfit in FSL |
| MOODS team DEP-ARREST-CLIN | 3T Philips Achieva | 2 x 2 x 2 mm | 60 directions b-factor=1500 | Eddy correction and DTIfit in FSL |
| EPISCA (Leiden adolescents) | 3T Philips Achieva | 2.3 x 2.3 x 2.3 mm | 32 directions; 1 b0 image; b-factor=1000 | Eddy current and motion correction in FSL |
| ETPB-STB | 3T GE HDx | 1.875 x 1.875 x 2.5 mm | 30 directions; b-factor=1100 | TORTOISE |
| FSL_NPL-OCD sample | 3T Siemens Magnetrom Allegra | 1.8 x 1.8 1.8 mm | 30 directions; 2 b0 images, b-factor=1000 | Eddy correction and DTIfit in FSL |
| FSL_NPL-SZ sample | 3T Siemens Magnetrom Allegra | 1.8 x 1.8 1.8 mm | 30 directions; 2 b0 images, b-factor=1000 | Eddy correction and DTIfit in FSL |
| FOR2107 Marburg | 3T Siemens Magnetom Trio | 2.5 x 2.5 x 2.5 mm | 30 directions; 4 b0 images; b-factor=1000 | Eddy correction and DTIfit in FSL |

| Site | Scanner vendor and type | Voxel size and slice thickness | Number of directions & b-factor | Preprocessing software & steps |
|---|---|---|---|---|
| FOR2107 Muenster | 3T Siemens PRISMA | 2.5 x 2.5 x 2.5 mm | 30 directions; 4 b0 images; b-factor=1000 | Eddy correction and DTIfit in FSL |
| Ghent | 3T Siemens Magnetom Trio Tim | 2 x 2 x 2 mm | 12 directions; b-factor=850 | Eddy and DTIfit in FSL |
| GIPSI BD | 3T Philips Ingenia | 1.75 x 1.75 x 2 mm | 16 directions; 1 b0 images, b-factor=1000 | FSL |
| GIPSI SZ | 3T Philips Ingenia | 1.75 x 1.75 x 2 mm | 16 directions; 1 b0 images, b-factor=1000 | FSL |
| Grady Trauma Project Emory University | 3T Siemens TIM-Trio | 2 x 2 x 2 mm | 138 directions; 6 b0 images; b-factor=1000 | Eddy current and motion correction in FSL |
| Halifax | 1.5T GE Signa | 1.875 x 1.875 x 2 mm | 31 directions; 2 b0 images; b-factor=900 | Eddy current correction in FSL |
| Houston BD | 3T Philips Ingenia | 1 x 1 x 1 mm | 42 directions; b-factor=1000 | Eddy currents, BET, DTIfit in FSL |
| Jena-SB | 3T Siemens Tim Trio | 1.25 x 1.25 x 2.5 $mm^3$ | 70 directions; *b* = 1000 $s/mm^2$, six *b* = 0 images | Denoising step, Eddy and DTIfit in FSL |
| KASP | 3T GE | 0.94 x 0.94 x 2.9 mm | 60 directions; 10 b) images;b-factor=1000 | Denoising, Gibbs ringing correction, Eddy currents, motion and DTIfit in FSL |
| LTS Colorado site 1 | 3T Siemens Tim Trio | 2 x 2 x 2 mm | 173 directions; b-factor=3000 | FSL Topup |
| LTS Colorado site 2 | 3T Siemens Prisma | 2 x 2 x 2 mm | 173 directions; b-factor=3000 | FSL Topup |
| McGill University | 3T Siemens Trio | 2 x 2 x 2 mm | 64 directions; b-factor=1000 | Eddy current correction, TOPUP |
| MR-IMPACT | 3T Magnetom Trio Tim | 2 x 2 x 2 mm | 13 direction; 5 b0 images; b-factor=350/1600[62] | Eddy in FSL |
| Muenster Neuroimaging Cohort | 3T Philips Gyroscan Intera | 1.8 x 1.8 x 3.6 mm | 25 directions; 5 b0 images; b-factor=1000 | Eddy current & motion correction in ACID Toolbox |
| PAFIP | 3T Philips Achieva | 2 x 2 x 2 mm | 64 directions; b-factor=1300 | Eddy current correction in FSL |

| Site | Scanner vendor and type | Voxel size and slice thickness | Number of directions & b-factor | Preprocessing software & steps |
|---|---|---|---|---|
| San Raffaele Hospital | 3T Philips Intera / 3T Philips Ingenia | 2 x 2 x 2 mm | 35 directions; 1 b0 image; b-factor=900/1000 | Eddy correction, BET in FSL |
| Stanford University FAA | 3T GE DIscovery MR750 | 2 x 2 x 2 mm | 96 directions; 9b0 images; b-factor=2000 | Eddy current and motion correction in FSL |
| STRADL site Aberdeen | 3T Philips Achieva-TX | 2.3 x 2.3 x 2.3 mm | 64 directions; b-factor=1200 | Eddy in FSL |
| STRADL site Dundee | 3T Siemens Prisma-FIT | 2.3 x 2.3 x 2.3 mm | 64 directions; b-factor=1200 | Eddy in FSL |
| Sydney Bipolar Kids and Siblings | 3T Philips Achieva | 1 x 2.5 x 2.5 mm | 32 directions; b-factor=1000 | Pre-processing using FDT in FSL |
| Sydney Brain and Mind Centre | 3T GE Discovery MR750 | 0.86 x 0.86 x 2 mm | 69 directions; 8 b0 images; b-factor=1159 | Eddy current & motion correction in FSL |
| TIPS-Jena | 3T Siemens Prisma Fit | 1 x 1 x 1 mm | 54 directions; 7 b0 images; b-factor=1200 | FSL Topup based pipeline |
| UCSF Adolescent MDD | 3T GE Discovery MR750 | 1.875 x 1.875 x 2.5 mm | 30 directions; 1 b0 image; b-factor=1500 | Fieldmap correction, eddy current correction, matrix rotation in FSL & AFNI |
| University of Minnesota Adolescent MDD | 3T Siemens Trio | 2 x 2 x 2 mm | 64 directions; b-factor=1000 | Eddy current and distortion correction in FSL |
| University of Texas - Austin Bipolar Seed Program | 3T Siemens Skyra | 2 x 2 x 2 mm | 30 directions; b-factor=1000 | Eddy correction, DTIfit in FSL |
| University of Washington/ Harvard | 3T Philips Achieva | 2 x 2 x 2 mm | 32 directions; b-factor=1200 | Eddy correction in FSL |
| Yale School of Medicine | 3T Siemens Trio | 3 x 3 x 3 mm | 32 directions; b-factor=1000 | Eddy current correction in FSL |

Table S3: List of white matter tracts of interest

| Abbreviation | Full tract name |
|---|---|
| AverageFA | Full skeleton average FA |
| ACR (L+R) | Anterior corona radiata |
| ALIC (L+R) | Anterior limb of internal capsule |
| BCC | Body of corpus callosum |
| CC (BCC+GCC+SCC) | Corpus callosum |
| CGC (L+R) | Cingulum (cingulate gyrus) |
| CGH (L+R) | Cingulum (hippocampal portion) |
| CR (L+R) | Corona radiata |
| CST (L+R) | Corticospinal tract |
| EC (L+R) | External capsule |
| FX | Fornix |

| | |
|---|---|
| FXST (L+R) | Fornix (cres) / Stria terminalis |
| GCC | Genu of corpus callosum |
| IC (L+R) | Internal capsule |
| IFO (L+R) | Inferior fronto-occipital fasciculus |
| PCR (L+R) | Posterior corona radiata |
| PLIC (L+R) | Posterior limb of internal capsule |
| PTR (L+R) | Posterior thalamic radiation |
| RLIC (L+R) | Retrolenticular part of internal capsule |
| SCC | Splenium of corpus callosum |
| SCR (L+R) | Superior corona radiata |
| SFO (L+R) | Superior fronto-occipital fasciculus |
| SLF (L+R) | Superior longitudinal fasciculus |
| SS (L+R) | Sagittal stratum |

| | |
|---|---|
| UNC (L+R) | Uncinate fasciculus |

Table S4. Definitions used for recent suicidal ideation and history of suicide attempt

| Site | Definition<br>recent suicidal ideation | Definition<br>history of suicide attempt |
|---|---|---|
| BiDIRECT | Hamilton Depression Rating Scale suicidal ideation item scored 2 (“Wishes to be dead or thinks about possible death”) or 3 (“Ideas or gestures of suicide) | Clinical interview: number of lifetime suicide attempts > 0 |
| Brazilian High Risk Cohort Porto Allegre | CBCL item 91 (“Talks about killing oneself in past six months”) is scored 1 (“Somewhat or sometimes true”) or 2 (“very true or often true”) | NA |
| Chiba University | Beck Depression Inventory item on suicidal ideation scored 1 (“I think about ending my life but would not do it”), 2 (“I would like to end my life”), 3 (“I would kill myself if I had the opportunity”) | NA |
| CHU Montpellier BICS | Columbia Suicide Severity Rating Scale (C-SSRS) positive answer on one or more of the following items current non-specific active suicidal ideation, current active suicidal ideation with any method (not plan) without intent to act, current active suicidal ideation with some intent without specific plan, current active suicidal ideation with specific plan and intent | Columbia Suicide Severity Rating Scale (C-SSRS) question on lifetime actual suicide attempt answered “yes” |
| CHU Montpellier IMPACT | Inventory of Depressive Symptomatology (Clinician-related; IDS-C) suicidal ideation item scored 2 (thinks of suicide or death multiple times per week for multiple minutes) or 3 (seriously thinks about suicide or death many times per day, or has prepared suicide or has tried to end life) OR Quick Inventory of Depression Symptomatlogy suicidal ideation item scored 2 (“I think about suicide or death multiple times per week for multiple minutes”) or 3 (“I think about suicide or death multiple times per day, or I have prepared suicide or I have tried to end my life”) | Columbia Suicide Severity Rating Scale (C-SSRS) question on lifetime actual suicide attempt answered “yes” |
| CHU Montpellier UF8555 | Scale for Suicidal Ideation (SSI) item: 4 (“Active Suicidal Ideation”) scored 1 (“weak”) OR 2 (“medium to strong) OR SSI item 5 (“passive suicidal thoughts) scored 2 (“would avoid doing what is necessary to stay alive; such as ending diabetes treatment”) OR Beck Depression Inventory item on suicidal ideation scored 1 (“I think about ending my life but would not do it”), 2 (“I would like to end my life”), 3 (“I would kill myself if I had the opportunity”) OR Hamilton Depression Rating Scale suicidal ideation item scored 2 (“Wishes to be dead or thinks about possible death”) or 3 (“Ideas or gestures of suicide”) | Lifetime history of suicide attempt determined using an interview |
| DCHS | NA | Mini International Neuropsychiatric interview (MINI) question BQ14 (“Did you ever make a suicide attempt?”) scored 4 (“yes”) |
| MOODS team DEP-ARREST- | Hamilton Depression Rating Scale suicidal ideation item scored 2 (“Wishes to be dead or thinks about possible death”) or 3 (“Ideas or gestures of suicide”) | Recent OR past suicide attempt determined in an interview |

| | | |
|---|---|---|
| CLIN | | |
| EPISCA (Leiden adolescents) | Childhood Depression Inventory item on suicidal ideation scored 1 (“I think about ending my life but would not do it”) or 2 (“I want to end my life”) | NA |
| ETPB-STB | Beck Depression Inventory item on suicidal ideation scored 1 (“I think about ending my life but would not do it”), 2 (“I would like to end my life”), 3 (“I would kill myself if I had the opportunity”) OR Hamilton Depression Rating Scale suicidal ideation item scored 2 (“Wishes to be dead or thinks about possible death”) or 3 (“Ideas or gestures of suicide”) OR Montgomery-Asberg Depression Rating Scale (MADRS) item on suicidal ideation scored between 2 (“Weary of life, only fleeting suicidal thoughts”) and 6 (“Explicit plans for suicide when there is an opportunity. Active preparations for suicide”) OR Scale for Suicidal Ideation item 4 (“desire to end life”) scored “weak” or “moderate to severe” | Lifetime history of suicide attempt determined using an interview |
| FSL_NPL- OCD sample | Hamilton Depression Rating Scale suicidal ideation item scored 2 (“Wishes to be dead or thinks about possible death”) or 3 (“Ideas or gestures of suicide”) | Clinical interview: number of lifetime suicide attempts > 0 |
| FSL_NPL - SZ sample | NA | Lifetime history of suicide attempt determined using an interview |
| FOR2107 Marburg | Beck Depression Inventory item on suicidal ideation scored 1 (“I think about ending my life but would not do it”), 2 (“I would like to end my life”), 3 (“I would kill myself if I had the opportunity”) OR Hamilton Depression Rating Scale (HDRS) suicidal ideation item scored 2 “Wishes to be dead or thinks about possible death” or 3 (“Ideas or gestures of suicide”) | NA |
| FOR2107 Muenster | Beck Depression Inventory item on suicidal ideation scored 1 (“I think about ending my life but would not do it”), 2 (“I would like to end my life”), 3 (“I would kill myself if I had the opportunity”) OR Hamilton Depression Rating Scale (HDRS-21) suicidal ideation item scored 2 “Wishes to be dead or thinks about possible death” or 3 “Ideas or gestures of suicide” | NA |
| Ghent | NA | Lifetime history of suicide attempt determined using an interview |
| GIPSI BD | NA | Lifetime history of suicide attempt determined using an interview |
| GIPSI SZ | NA | Lifetime history of suicide attempt determined using an interview |

| | | |
|---|---|---|
| Grady Trauma Project Emory University | Beck Depression Inventory item on suicidal ideation scored 1 (“I think about ending my life but would not do it”), 2 (“I would like to end my life”), 3 (“I would kill myself if I had the opportunity”) | SSH item 1 (Have you ever tried to kill yourself or commit suicide?) scored “yes” |
| Halifax | NA | Lifetime history of suicide attempt determined using an interview |
| Houston BD | NA | Clinical interview: number of lifetime suicide attempts > 0 |
| Jena-SB | Beck Depression Inventory item on suicidal ideation scored 1 (“I think about ending my life but would not do it”), 2 (“I would like to end my life”), 3 (“I would kill myself if I had the opportunity”) OR Hamilton Depression Rating Scale suicidal ideation item scored 2 (“Wishes to be dead or thinks about possible death”) or 3 (Ideas or gestures of suicide) | NA |
| KASP | Calgary Depression Scale for Schizophrenia item 8 (“In the past two weeks, have you felt that life wasn’t worth living? Did you feel like ending it all? What did you think you might do?”) scored 1(“Frequent thoughts or being better off dead or occasional thoughts or suicide”) or higher | NA |
| LTS Colorado | NA | Lifetime history of suicide attempt determined using an interview |
| McGill University | Columbia Suicide Severity Rating Scale (C-SSRS) positive answer on one or more of the following items: current non-specific active suicidal ideation, current active suicidal ideation with any method (not plan) without intent to act, current active suicidal ideation with some intent without specific plan, current active suicidal ideation with specific plan and intent | Columbia Suicide Severity Rating Scale (C-SSRS) question on lifetime actual suicide attempt answered “yes” |
| MR-IMPACT | Columbia Suicide Severity Rating Scale (C-SSRS) positive answer on one or more of the following items (in the past month): current non-specific active suicidal ideation, current active suicidal ideation with any method (not plan) without intent to act, current active suicidal ideation with some intent without specific plan, current active suicidal ideation with specific plan and intent | Columbia Suicide Severity Rating Scale (C-SSRS) question on lifetime actual suicide attempt answered “yes” |
| Muenster Neuroimaging Cohort | Beck Depression Inventory-2 item on suicidal ideation scored 1 (“I thave thoughts of killing myself, but I would not carry them out”), 2 (“I would like to kill myself”), 3 (“I would kill myself if I had the chance”) OR Montgomery-Asberg Depression Rating Scale (MADRS) item on suicidal ideation scored between 2 (“Weary of life, only fleeting suicidal thoughts”) and 6 (“Explicit plans for suicide when there is an opportunity. Active preparations for suicide”) | NA |

| | | |
|---|---|---|
| PAFIP | NA | Lifetime history of suicide attempt determined using an interview |
| San Raffaele Hospital | Beck Depression Inventory item on suicidal ideation scored 1 (“I think about ending my life but would not do it”), 2 (“I would like to end my life”), 3 (“I would kill myself if I had the opportunity”) OR Hamilton Depression Rating Scale suicidal ideation item scored 2 (“Wishes to be dead or thinks about possible death”) or 3 (“Ideas or gestures of suicide”) | Clinical interview: number of lifetime suicide attempts > 0 |
| Stanford University FAA | Beck Depression Inventory item on suicidal ideation scored 1 (“I think about ending my life but would not do it”), 2 (“I would like to end my life”), 3 (“I would kill myself if I had the opportunity”) OR Hamilton Depression Rating Scale suicidal ideation item scored 2 (“Wishes to be dead or thinks about possible death”) or 3 (“Ideas or gestures of suicide”) | NA |
| STRADL | SCID item A21 (“Suicidal ideation during current major depressive episode”) scored 1 (“yes”) OR Quick Inventory of Depressive Symptomatology item 12 (“Thoughts of suicide or death”) is scored 2 (“I think of suicide or death several times a week for several minutes”) or higher | NA |
| Sydney Bipolar Kids and Siblings | NA | Diagnostic Interview for Genetic Studies (DIGS) question on lifetime suicide attempt answered “yes” OR Kiddie Schedule for Affective Disorders and Schzophrenia (KSADS) item on lifetime suicide attempt coded as “yes” |
| Sydney Brain and Mind Centre | Hamilton Depression Rating Scale suicidal ideation item scored 2 “Wishes to be dead or thinks about possible death” or 3 “Ideas or gestures of suicide” | Clinical interview: lifetime suicide attempt coded “yes” |
| TIPS-Jena | Beck Depression Inventory item on suicidal ideation scored 1 (“I think about ending my life but would not do it”), 2 (“I would like to end my life”), 3 (“I would kill myself if I had the opportunity”) OR Hamilton Depression Rating Scale suicidal ideation item scored 2 (“Wishes to be dead or thinks about possible death”) or 3 (“Ideas or gestures of suicide”) OR SCID item on current suicidal ideation scored (“yes”) | NA |
| UCSF Adolescent MDD | Columbia Suicide Severity Rating Scale (C-SSRS) positive answer on one or more of the following items (in the past week): current non-specific active suicidal ideation, current active suicidal ideation with any method (not plan) without intent to act, current active suicidal ideation with some intent without specific plan, current active suicidal ideation with specific plan and intent | Columbia Suicide Severity Rating Scale (C-SSRS) question on lifetime non-interrupted attempt answered “yes” |

| | | |
|---|---|---|
| University of Minnesota Adolescent MDD | Beck Depression Inventory-2 item on suicidal ideation scored 1 (“I have thoughts of killing myself, but I would not carry them out”), 2 (“I would like to kill myself”), 3 (“I would kill myself if I had the chance”) OR Childrens Depression Rating Scale 3 or higher | Kiddie Schedule for Affective Disorders and Schizophrenia for School Aged Children- Lifetime version (K-SADS-PL_2009) item on suicide attempt answer 1 |
| University of Texas - Austin - Mood Addiction Interaction Neuroscience (MAIN) Program | Structured Clinical Interview DSM-5 (SCID-RV/NP for DSM-5) Suicidality module: answer “yes” to question on ideation in the past week (“Have you in the past week wished you were dead or wished you could go to sleep and not wake up?”) | Structured Clinical Interview DSM-5 (SCID-RV/NP for DSM-5) Suicidality module: answer “yes” to question “Have you ever tried to kill yourself?” |
| University of Washington/ Harvard | NA | Self-injurious Thoughts and Behaviors Interview (SITBI) measure on lifetime suicide attempt answered “yes” |
| Yale School of Medicine | Columbia Suicide Severity Rating Scale (C-SSRS) positive answer on one or more of the following items (in the past month): current non-specific active suicidal ideation, current active suicidal ideation with any method (not plan) without intent to act, current active suicidal ideation with some intent without specific plan, current active suicidal ideation with specific plan and intent | Columbia Suicide Severity Rating Scale (C-SSRS) question on lifetime non-interrupted suicide attempt answered “yes” |

Table S5. Differences in regional AD between individuals with a history of suicide attempt and healthy controls.
Age, sex, age-by-sex, age$^2$, age$^2$-by-sex included as covariates.

| Region | Cohen's d | SE | CI LB | CI UB | P-value | FDR P-value | Attempt | HC |
|---|---|---|---|---|---|---|---|---|
| ACR | -0.070 | 0.060 | -0.188 | 0.048 | 0.245 | 0.557 | 601 | 511 |
| ALIC | -0.080 | 0.060 | -0.198 | 0.038 | 0.184 | 0.557 | 600 | 511 |
| BCC | 0.053 | 0.060 | -0.065 | 0.172 | 0.377 | 0.674 | 599 | 507 |
| CC | 0.016 | 0.060 | -0.102 | 0.134 | 0.788 | 0.946 | 600 | 508 |
| CGC | -0.101 | 0.060 | -0.219 | 0.016 | 0.092 | 0.557 | 604 | 512 |
| CGH | 0.008 | 0.060 | -0.110 | 0.126 | 0.899 | 0.962 | 600 | 512 |
| CR | -0.061 | 0.060 | -0.179 | 0.057 | 0.311 | 0.649 | 602 | 511 |
| CST | 0.031 | 0.060 | -0.087 | 0.149 | 0.606 | 0.891 | 602 | 510 |
| EC | -0.055 | 0.060 | -0.173 | 0.063 | 0.364 | 0.674 | 601 | 512 |
| FX | 0.033 | 0.060 | -0.085 | 0.151 | 0.582 | 0.891 | 603 | 512 |
| FXST | 0.001 | 0.060 | -0.117 | 0.119 | 0.986 | 0.986 | 604 | 512 |
| GCC | -0.016 | 0.060 | -0.134 | 0.102 | 0.795 | 0.946 | 602 | 509 |
| IC | -0.077 | 0.060 | -0.195 | 0.041 | 0.205 | 0.557 | 600 | 510 |
| IFO | -0.085 | 0.061 | -0.205 | 0.036 | 0.170 | 0.557 | 598 | 475 |
| PCR | -0.075 | 0.060 | -0.193 | 0.043 | 0.212 | 0.557 | 602 | 509 |
| PLIC | -0.043 | 0.060 | -0.161 | 0.074 | 0.471 | 0.785 | 602 | 511 |
| PTR | 0.006 | 0.060 | -0.112 | 0.124 | 0.923 | 0.962 | 603 | 511 |
| RLIC | -0.080 | 0.060 | -0.198 | 0.038 | 0.184 | 0.557 | 603 | 512 |
| SCC | 0.024 | 0.060 | -0.094 | 0.142 | 0.691 | 0.946 | 600 | 509 |
| SCR | -0.071 | 0.060 | -0.189 | 0.047 | 0.242 | 0.557 | 601 | 510 |
| SFO | -0.011 | 0.060 | -0.129 | 0.107 | 0.854 | 0.962 | 601 | 511 |
| SLF | -0.098 | 0.060 | -0.216 | 0.020 | 0.105 | 0.557 | 604 | 512 |
| SS | -0.016 | 0.060 | -0.134 | 0.102 | 0.795 | 0.946 | 601 | 511 |
| UNC | 0.154 | 0.063 | 0.030 | 0.277 | 0.015 | 0.375 | 582 | 447 |
| AverageAD | -0.081 | 0.060 | -0.199 | 0.037 | 0.181 | 0.557 | 603 | 511 |

Table S6. Differences in regional FA between individuals with a history of suicide attempt and healthy controls. Age, sex, age-by-sex, $age^2$, $age^2$-by-sex included as covariates.

| Region | Cohen's d | SE | CI LB | CI UB | P-value | FDR P-value | Attempt | HC |
|---|---|---|---|---|---|---|---|---|
| ACR | -0.094 | 0.057 | -0.206 | 0.018 | 0.101 | 0.127 | 636 | 591 |
| ALIC | -0.144 | 0.057 | -0.256 | -0.032 | 0.012 | 0.022 | 635 | 591 |
| BCC | -0.182 | 0.057 | -0.295 | -0.069 | 0.002 | 0.005 | 630 | 588 |
| CC | -0.223 | 0.057 | -0.336 | -0.111 | 0.000 | 0.001 | 631 | 591 |
| CGC | -0.167 | 0.057 | -0.279 | -0.055 | 0.004 | 0.008 | 638 | 589 |
| CGH | -0.040 | 0.057 | -0.152 | 0.072 | 0.488 | 0.530 | 635 | 590 |
| CR | -0.169 | 0.057 | -0.281 | -0.056 | 0.003 | 0.008 | 636 | 590 |
| CST | -0.011 | 0.057 | -0.123 | 0.101 | 0.849 | 0.849 | 635 | 588 |
| EC | -0.168 | 0.057 | -0.280 | -0.056 | 0.003 | 0.008 | 637 | 591 |
| FX | -0.058 | 0.057 | -0.170 | 0.055 | 0.315 | 0.358 | 632 | 589 |
| FXST | -0.201 | 0.057 | -0.313 | -0.089 | 0.000 | 0.002 | 636 | 590 |
| GCC | -0.200 | 0.057 | -0.313 | -0.088 | 0.000 | 0.002 | 635 | 590 |
| IC | -0.116 | 0.057 | -0.228 | -0.003 | 0.044 | 0.058 | 636 | 589 |
| IFO | -0.130 | 0.058 | -0.245 | -0.015 | 0.027 | 0.040 | 633 | 546 |
| PCR | -0.223 | 0.057 | -0.335 | -0.110 | 0.000 | 0.001 | 638 | 591 |
| PLIC | -0.029 | 0.057 | -0.141 | 0.083 | 0.609 | 0.634 | 635 | 590 |
| PTR | -0.171 | 0.057 | -0.283 | -0.058 | 0.003 | 0.008 | 632 | 590 |
| RLIC | -0.125 | 0.057 | -0.238 | -0.013 | 0.029 | 0.040 | 634 | 591 |
| SCC | -0.202 | 0.057 | -0.315 | -0.090 | 0.000 | 0.002 | 632 | 589 |
| SCR | -0.139 | 0.057 | -0.251 | -0.027 | 0.015 | 0.026 | 635 | 591 |
| SFO | -0.086 | 0.057 | -0.199 | 0.026 | 0.133 | 0.158 | 633 | 588 |
| SLF | -0.226 | 0.057 | -0.339 | -0.113 | 0.000 | 0.001 | 633 | 590 |
| SS | -0.127 | 0.057 | -0.239 | -0.015 | 0.027 | 0.040 | 635 | 591 |
| UNC | -0.172 | 0.059 | -0.288 | -0.055 | 0.004 | 0.008 | 616 | 526 |
| AverageFA | -0.195 | 0.057 | -0.307 | -0.083 | 0.001 | 0.003 | 635 | 589 |

Table S7. Differences in regional MD between individuals with a history of suicide attempt and healthy controls.
Age, sex, age-by-sex, age$^2$, age$^2$-by-sex included as covariates.

| Region | Cohen's d | SE | CI LB | CI UB | P-value | FDR P-value | Attempt | HC |
|---|---|---|---|---|---|---|---|---|
| ACR | 0.050 | 0.059 | -0.065 | 0.165 | 0.392 | 0.577 | 614 | 553 |
| ALIC | 0.002 | 0.059 | -0.114 | 0.117 | 0.979 | 0.997 | 611 | 550 |
| BCC | 0.141 | 0.059 | 0.025 | 0.256 | 0.017 | 0.086 | 610 | 549 |
| CC | 0.141 | 0.059 | 0.025 | 0.256 | 0.017 | 0.086 | 614 | 550 |
| CGC | -0.013 | 0.059 | -0.128 | 0.102 | 0.831 | 0.997 | 614 | 552 |
| CGH | -0.057 | 0.059 | -0.173 | 0.058 | 0.330 | 0.516 | 612 | 551 |
| CR | 0.094 | 0.059 | -0.021 | 0.209 | 0.109 | 0.272 | 615 | 551 |
| CST | 0.000 | 0.059 | -0.115 | 0.115 | 0.997 | 0.997 | 612 | 552 |
| EC | 0.060 | 0.059 | -0.055 | 0.176 | 0.305 | 0.509 | 613 | 550 |
| FX | 0.077 | 0.059 | -0.038 | 0.192 | 0.192 | 0.395 | 613 | 547 |
| FXST | 0.143 | 0.059 | 0.028 | 0.258 | 0.015 | 0.086 | 614 | 552 |
| GCC | 0.096 | 0.059 | -0.018 | 0.211 | 0.101 | 0.272 | 617 | 553 |
| IC | 0.005 | 0.059 | -0.110 | 0.120 | 0.934 | 0.997 | 611 | 550 |
| IFO | -0.020 | 0.060 | -0.138 | 0.098 | 0.737 | 0.969 | 608 | 508 |
| PCR | 0.102 | 0.059 | -0.013 | 0.217 | 0.084 | 0.272 | 615 | 550 |
| PLIC | -0.010 | 0.059 | -0.126 | 0.105 | 0.862 | 0.997 | 609 | 549 |
| PTR | 0.099 | 0.059 | -0.016 | 0.214 | 0.094 | 0.272 | 615 | 551 |
| RLIC | 0.034 | 0.059 | -0.081 | 0.149 | 0.559 | 0.777 | 616 | 551 |
| SCC | 0.164 | 0.059 | 0.049 | 0.280 | 0.005 | 0.067 | 614 | 550 |
| SCR | 0.098 | 0.059 | -0.017 | 0.213 | 0.095 | 0.272 | 616 | 551 |
| SFO | 0.007 | 0.059 | -0.108 | 0.122 | 0.905 | 0.997 | 611 | 554 |
| SLF | 0.084 | 0.059 | -0.031 | 0.200 | 0.152 | 0.346 | 612 | 551 |
| SS | 0.075 | 0.059 | -0.040 | 0.190 | 0.205 | 0.395 | 612 | 553 |
| UNC | 0.220 | 0.061 | 0.099 | 0.340 | 0.000 | 0.009 | 591 | 487 |
| AverageMD | 0.062 | 0.059 | -0.053 | 0.177 | 0.290 | 0.509 | 615 | 549 |

Table S8. Differences in regional RD between individuals with a history of suicide attempt and healthy controls. Age, sex, age-by-sex, age$^2$, age$^2$-by-sex included as covariates.

| Region | Cohen's d | SE | CI LB | CI UB | P-value | FDR P-value | Attempt | HC |
|---|---|---|---|---|---|---|---|---|
| ACR | 0.096 | 0.059 | -0.019 | 0.211 | 0.102 | 0.175 | 613 | 553 |
| ALIC | 0.050 | 0.059 | -0.065 | 0.166 | 0.392 | 0.515 | 610 | 551 |
| BCC | 0.153 | 0.059 | 0.037 | 0.268 | 0.010 | 0.036 | 610 | 549 |
| CC | 0.152 | 0.059 | 0.036 | 0.267 | 0.010 | 0.036 | 612 | 552 |
| CGC | 0.048 | 0.059 | -0.067 | 0.163 | 0.412 | 0.515 | 614 | 552 |
| CGH | -0.027 | 0.059 | -0.142 | 0.088 | 0.652 | 0.741 | 613 | 553 |
| CR | 0.148 | 0.059 | 0.033 | 0.263 | 0.012 | 0.037 | 614 | 552 |
| CST | -0.011 | 0.059 | -0.126 | 0.104 | 0.848 | 0.884 | 611 | 551 |
| EC | 0.086 | 0.059 | -0.029 | 0.201 | 0.143 | 0.224 | 614 | 553 |
| FX | 0.104 | 0.059 | -0.011 | 0.220 | 0.077 | 0.175 | 612 | 545 |
| FXST | 0.154 | 0.059 | 0.039 | 0.269 | 0.009 | 0.036 | 616 | 553 |
| GCC | 0.117 | 0.059 | 0.002 | 0.232 | 0.047 | 0.118 | 613 | 553 |
| IC | 0.059 | 0.059 | -0.056 | 0.174 | 0.318 | 0.442 | 613 | 551 |
| IFO | 0.029 | 0.060 | -0.088 | 0.147 | 0.627 | 0.741 | 612 | 508 |
| PCR | 0.168 | 0.059 | 0.053 | 0.283 | 0.004 | 0.036 | 617 | 551 |
| PLIC | -0.008 | 0.059 | -0.124 | 0.107 | 0.886 | 0.886 | 610 | 552 |
| PTR | 0.101 | 0.059 | -0.014 | 0.216 | 0.086 | 0.175 | 614 | 552 |
| RLIC | 0.082 | 0.059 | -0.033 | 0.197 | 0.162 | 0.238 | 615 | 553 |
| SCC | 0.166 | 0.059 | 0.050 | 0.281 | 0.005 | 0.036 | 613 | 550 |
| SCR | 0.137 | 0.059 | 0.021 | 0.252 | 0.020 | 0.057 | 613 | 552 |
| SFO | 0.017 | 0.059 | -0.098 | 0.132 | 0.769 | 0.836 | 612 | 553 |
| SLF | 0.161 | 0.059 | 0.046 | 0.277 | 0.006 | 0.036 | 614 | 552 |
| SS | 0.095 | 0.059 | -0.020 | 0.210 | 0.105 | 0.175 | 613 | 554 |
| UNC | 0.170 | 0.061 | 0.050 | 0.290 | 0.006 | 0.036 | 594 | 487 |
| AverageRD | 0.098 | 0.059 | -0.017 | 0.213 | 0.096 | 0.175 | 614 | 552 |

Table S9. Differences in regional AD between individuals with a history of suicide attempt and clinical controls. Age, sex, age-by-sex, age$^2$, age$^2$-by-sex included as covariates.

| Region | Cohen's d | SE | CI LB | CI UB | P-value | FDR P-value | Attempt | CLC |
|---|---|---|---|---|---|---|---|---|
| ACR | 0.010 | 0.048 | -0.083 | 0.103 | 0.836 | 0.968 | 601 | 1680 |
| ALIC | 0.024 | 0.048 | -0.069 | 0.117 | 0.615 | 0.968 | 600 | 1677 |
| BCC | 0.049 | 0.048 | -0.044 | 0.142 | 0.304 | 0.968 | 599 | 1679 |
| CC | 0.038 | 0.048 | -0.055 | 0.132 | 0.421 | 0.968 | 600 | 1676 |
| CGC | 0.007 | 0.047 | -0.086 | 0.100 | 0.886 | 0.968 | 604 | 1683 |
| CGH | -0.009 | 0.048 | -0.102 | 0.085 | 0.856 | 0.968 | 600 | 1682 |
| CR | 0.029 | 0.048 | -0.064 | 0.123 | 0.537 | 0.968 | 602 | 1676 |
| CST | 0.018 | 0.048 | -0.075 | 0.111 | 0.709 | 0.968 | 602 | 1678 |
| EC | 0.011 | 0.048 | -0.082 | 0.104 | 0.814 | 0.968 | 601 | 1669 |
| FX | -0.011 | 0.047 | -0.104 | 0.082 | 0.811 | 0.968 | 603 | 1685 |
| FXST | -0.016 | 0.047 | -0.109 | 0.077 | 0.733 | 0.968 | 604 | 1683 |
| GCC | 0.024 | 0.048 | -0.069 | 0.118 | 0.608 | 0.968 | 602 | 1680 |
| IC | -0.002 | 0.048 | -0.095 | 0.091 | 0.966 | 0.968 | 600 | 1675 |
| IFO | -0.042 | 0.048 | -0.135 | 0.052 | 0.380 | 0.968 | 598 | 1661 |
| PCR | 0.002 | 0.048 | -0.091 | 0.095 | 0.968 | 0.968 | 602 | 1675 |
| PLIC | 0.014 | 0.048 | -0.079 | 0.107 | 0.768 | 0.968 | 602 | 1678 |
| PTR | 0.021 | 0.047 | -0.072 | 0.114 | 0.665 | 0.968 | 603 | 1677 |
| RLIC | -0.043 | 0.047 | -0.136 | 0.050 | 0.363 | 0.968 | 603 | 1676 |
| SCC | 0.017 | 0.048 | -0.076 | 0.111 | 0.716 | 0.968 | 600 | 1678 |
| SCR | 0.035 | 0.048 | -0.058 | 0.128 | 0.464 | 0.968 | 601 | 1671 |
| SFO | 0.043 | 0.048 | -0.051 | 0.136 | 0.372 | 0.968 | 601 | 1673 |
| SLF | 0.002 | 0.047 | -0.091 | 0.095 | 0.960 | 0.968 | 604 | 1680 |
| SS | -0.013 | 0.048 | -0.106 | 0.080 | 0.786 | 0.968 | 601 | 1676 |
| UNC | 0.101 | 0.048 | 0.006 | 0.196 | 0.037 | 0.922 | 582 | 1615 |
| AverageAD | -0.016 | 0.048 | -0.110 | 0.077 | 0.729 | 0.968 | 603 | 1672 |

Table S10. Differences in regional FA between individuals with a history of suicide attempt and clinical controls. Age, sex, age-by-sex, $age^2$, $age^2$-by-sex included as covariates.

| Region | Cohen's d | SE | CI LB | CI UB | P-value | FDR P-value | Attempt | CLC |
|---|---|---|---|---|---|---|---|---|
| ACR | -0.102 | 0.046 | -0.192 | -0.011 | 0.028 | 0.071 | 636 | 1802 |
| ALIC | -0.094 | 0.046 | -0.185 | -0.004 | 0.041 | 0.080 | 635 | 1794 |
| BCC | -0.066 | 0.046 | -0.157 | 0.024 | 0.153 | 0.201 | 630 | 1795 |
| CC | -0.104 | 0.046 | -0.195 | -0.013 | 0.025 | 0.071 | 631 | 1792 |
| CGC | -0.099 | 0.046 | -0.189 | -0.009 | 0.032 | 0.071 | 638 | 1797 |
| CGH | -0.005 | 0.046 | -0.096 | 0.085 | 0.912 | 0.956 | 635 | 1797 |
| CR | -0.126 | 0.046 | -0.217 | -0.036 | 0.006 | 0.034 | 636 | 1795 |
| CST | -0.002 | 0.046 | -0.092 | 0.089 | 0.969 | 0.969 | 635 | 1797 |
| EC | -0.067 | 0.046 | -0.158 | 0.023 | 0.145 | 0.201 | 637 | 1792 |
| FX | -0.032 | 0.046 | -0.122 | 0.059 | 0.495 | 0.590 | 632 | 1792 |
| FXST | -0.126 | 0.046 | -0.217 | -0.036 | 0.006 | 0.034 | 636 | 1795 |
| GCC | -0.061 | 0.046 | -0.151 | 0.030 | 0.190 | 0.237 | 635 | 1801 |
| IC | -0.088 | 0.046 | -0.178 | 0.003 | 0.057 | 0.096 | 636 | 1794 |
| IFO | 0.005 | 0.046 | -0.086 | 0.096 | 0.918 | 0.956 | 633 | 1770 |
| PCR | -0.138 | 0.046 | -0.228 | -0.047 | 0.003 | 0.034 | 638 | 1797 |
| PLIC | -0.015 | 0.046 | -0.106 | 0.075 | 0.744 | 0.846 | 635 | 1793 |
| PTR | -0.124 | 0.046 | -0.214 | -0.033 | 0.008 | 0.034 | 632 | 1797 |
| RLIC | -0.100 | 0.046 | -0.190 | -0.009 | 0.031 | 0.071 | 634 | 1792 |
| SCC | -0.123 | 0.046 | -0.214 | -0.032 | 0.008 | 0.034 | 632 | 1791 |
| SCR | -0.077 | 0.046 | -0.167 | 0.014 | 0.098 | 0.153 | 635 | 1798 |
| SFO | -0.098 | 0.046 | -0.189 | -0.007 | 0.034 | 0.071 | 633 | 1799 |
| SLF | -0.125 | 0.046 | -0.216 | -0.034 | 0.007 | 0.034 | 633 | 1802 |
| SS | -0.107 | 0.046 | -0.198 | -0.017 | 0.020 | 0.071 | 635 | 1794 |
| UNC | -0.069 | 0.047 | -0.161 | 0.023 | 0.139 | 0.201 | 616 | 1732 |
| AverageFA | -0.093 | 0.046 | -0.183 | -0.002 | 0.045 | 0.080 | 635 | 1793 |

Table S11. Differences in regional MD between individuals with a history of suicide attempt and clinical controls. Age, sex, age-by-sex, age$^2$, age$^2$-by-sex included as covariates.

| Region | Cohen's d | SE | CI LB | CI UB | P-value | FDR P-value | Attempt | CLC |
|---|---|---|---|---|---|---|---|---|
| ACR | 0.070 | 0.047 | -0.021 | 0.162 | 0.133 | 0.278 | 614 | 1764 |
| ALIC | 0.046 | 0.047 | -0.046 | 0.138 | 0.330 | 0.551 | 611 | 1759 |
| BCC | 0.081 | 0.047 | -0.012 | 0.173 | 0.087 | 0.199 | 610 | 1760 |
| CC | 0.080 | 0.047 | -0.012 | 0.172 | 0.088 | 0.199 | 614 | 1764 |
| CGC | 0.036 | 0.047 | -0.056 | 0.128 | 0.442 | 0.631 | 614 | 1764 |
| CGH | -0.032 | 0.047 | -0.124 | 0.060 | 0.492 | 0.638 | 612 | 1764 |
| CR | 0.100 | 0.047 | 0.009 | 0.192 | 0.032 | 0.199 | 615 | 1762 |
| CST | 0.000 | 0.047 | -0.092 | 0.092 | 0.993 | 0.993 | 612 | 1750 |
| EC | 0.030 | 0.047 | -0.062 | 0.122 | 0.521 | 0.638 | 613 | 1760 |
| FX | 0.015 | 0.047 | -0.077 | 0.107 | 0.747 | 0.778 | 613 | 1755 |
| FXST | 0.092 | 0.047 | 0.001 | 0.184 | 0.049 | 0.199 | 614 | 1759 |
| GCC | 0.035 | 0.047 | -0.057 | 0.127 | 0.454 | 0.631 | 617 | 1767 |
| IC | 0.027 | 0.047 | -0.065 | 0.119 | 0.571 | 0.648 | 611 | 1761 |
| IFO | -0.067 | 0.047 | -0.159 | 0.026 | 0.158 | 0.304 | 608 | 1729 |
| PCR | 0.086 | 0.047 | -0.005 | 0.178 | 0.066 | 0.199 | 615 | 1757 |
| PLIC | 0.029 | 0.047 | -0.063 | 0.121 | 0.536 | 0.638 | 609 | 1762 |
| PTR | 0.081 | 0.047 | -0.011 | 0.173 | 0.085 | 0.199 | 615 | 1762 |
| RLIC | 0.025 | 0.047 | -0.067 | 0.117 | 0.596 | 0.648 | 616 | 1764 |
| SCC | 0.088 | 0.047 | -0.004 | 0.180 | 0.062 | 0.199 | 614 | 1754 |
| SCR | 0.113 | 0.047 | 0.021 | 0.204 | 0.016 | 0.199 | 616 | 1760 |
| SFO | 0.102 | 0.047 | 0.010 | 0.194 | 0.030 | 0.199 | 611 | 1755 |
| SLF | 0.084 | 0.047 | -0.008 | 0.176 | 0.075 | 0.199 | 612 | 1758 |
| SS | 0.053 | 0.047 | -0.039 | 0.145 | 0.258 | 0.460 | 612 | 1756 |
| UNC | 0.143 | 0.048 | 0.049 | 0.237 | 0.003 | 0.070 | 591 | 1692 |
| AverageMD | 0.038 | 0.047 | -0.054 | 0.129 | 0.424 | 0.631 | 615 | 1763 |

Table S12. Differences in regional RD between individuals with a history of suicide attempt and clinical controls. Age, sex, age-by-sex, age$^2$, age$^2$-by-sex included as covariates.

| Region | Cohen's d | SE | CI LB | CI UB | P-value | FDR P-value | Attempt | CLC |
|---|---|---|---|---|---|---|---|---|
| ACR | 0.093 | 0.047 | 0.001 | 0.185 | 0.048 | 0.120 | 613 | 1761 |
| ALIC | 0.046 | 0.047 | -0.046 | 0.139 | 0.324 | 0.470 | 610 | 1758 |
| BCC | 0.070 | 0.047 | -0.022 | 0.162 | 0.137 | 0.264 | 610 | 1763 |
| CC | 0.080 | 0.047 | -0.012 | 0.172 | 0.088 | 0.183 | 612 | 1764 |
| CGC | 0.045 | 0.047 | -0.047 | 0.137 | 0.339 | 0.470 | 614 | 1762 |
| CGH | -0.039 | 0.047 | -0.131 | 0.053 | 0.408 | 0.510 | 613 | 1763 |
| CR | 0.120 | 0.047 | 0.028 | 0.212 | 0.010 | 0.079 | 614 | 1762 |
| CST | -0.018 | 0.047 | -0.110 | 0.074 | 0.706 | 0.706 | 611 | 1762 |
| EC | 0.036 | 0.047 | -0.056 | 0.128 | 0.443 | 0.527 | 614 | 1757 |
| FX | 0.019 | 0.047 | -0.073 | 0.111 | 0.680 | 0.706 | 612 | 1755 |
| FXST | 0.107 | 0.047 | 0.015 | 0.198 | 0.023 | 0.082 | 616 | 1760 |
| GCC | 0.042 | 0.047 | -0.050 | 0.134 | 0.374 | 0.492 | 613 | 1762 |
| IC | 0.024 | 0.047 | -0.068 | 0.116 | 0.606 | 0.659 | 613 | 1761 |
| IFO | -0.057 | 0.047 | -0.149 | 0.036 | 0.229 | 0.382 | 612 | 1728 |
| PCR | 0.129 | 0.047 | 0.037 | 0.221 | 0.006 | 0.079 | 617 | 1759 |
| PLIC | -0.026 | 0.047 | -0.119 | 0.066 | 0.575 | 0.653 | 610 | 1757 |
| PTR | 0.107 | 0.047 | 0.015 | 0.199 | 0.023 | 0.082 | 614 | 1761 |
| RLIC | 0.058 | 0.047 | -0.034 | 0.150 | 0.215 | 0.382 | 615 | 1761 |
| SCC | 0.097 | 0.047 | 0.005 | 0.189 | 0.040 | 0.120 | 613 | 1752 |
| SCR | 0.107 | 0.047 | 0.015 | 0.199 | 0.023 | 0.082 | 613 | 1763 |
| SFO | 0.090 | 0.047 | -0.002 | 0.182 | 0.057 | 0.129 | 612 | 1761 |
| SLF | 0.121 | 0.047 | 0.029 | 0.213 | 0.010 | 0.079 | 614 | 1762 |
| SS | 0.093 | 0.047 | 0.001 | 0.185 | 0.047 | 0.120 | 613 | 1757 |
| UNC | 0.119 | 0.048 | 0.026 | 0.213 | 0.013 | 0.079 | 594 | 1692 |
| AverageRD | 0.047 | 0.047 | -0.045 | 0.139 | 0.320 | 0.470 | 614 | 1762 |

**Recent suicidal ideation**

Table S13. Differences in regional AD between individuals with recent suicidal ideation and healthy controls.
Age, sex, age-by-sex, age$^2$, age$^2$-by-sex included as covariates.

| Region | Cohen's d | SE | CI LB | CI UB | P-value | FDR P-value | Ideation | HC |
|---|---|---|---|---|---|---|---|---|
| ACR | -0.065 | 0.048 | -0.159 | 0.029 | 0.178 | 0.581 | 677 | 1217 |
| ALIC | -0.036 | 0.048 | -0.130 | 0.058 | 0.452 | 0.753 | 675 | 1206 |
| BCC | -0.055 | 0.048 | -0.149 | 0.039 | 0.256 | 0.581 | 675 | 1211 |
| CC | -0.085 | 0.048 | -0.179 | 0.009 | 0.075 | 0.520 | 678 | 1214 |
| CGC | -0.061 | 0.048 | -0.155 | 0.033 | 0.206 | 0.581 | 677 | 1214 |
| CGH | 0.056 | 0.048 | -0.038 | 0.150 | 0.247 | 0.581 | 673 | 1216 |
| CR | -0.046 | 0.048 | -0.140 | 0.048 | 0.342 | 0.713 | 676 | 1216 |
| CST | 0.009 | 0.048 | -0.085 | 0.104 | 0.845 | 0.845 | 674 | 1206 |
| EC | -0.012 | 0.048 | -0.106 | 0.082 | 0.806 | 0.839 | 669 | 1217 |
| FX | 0.025 | 0.048 | -0.069 | 0.119 | 0.606 | 0.802 | 676 | 1214 |
| FXST | 0.041 | 0.048 | -0.053 | 0.135 | 0.390 | 0.749 | 677 | 1219 |
| GCC | -0.071 | 0.048 | -0.165 | 0.023 | 0.138 | 0.576 | 677 | 1215 |
| IC | -0.016 | 0.048 | -0.110 | 0.078 | 0.734 | 0.802 | 674 | 1214 |
| IFO | 0.104 | 0.048 | 0.010 | 0.198 | 0.031 | 0.520 | 676 | 1213 |
| PCR | -0.023 | 0.048 | -0.117 | 0.071 | 0.631 | 0.802 | 675 | 1212 |
| PLIC | -0.016 | 0.048 | -0.110 | 0.078 | 0.737 | 0.802 | 673 | 1216 |
| PTR | 0.033 | 0.048 | -0.061 | 0.127 | 0.495 | 0.773 | 674 | 1217 |
| RLIC | 0.020 | 0.048 | -0.074 | 0.114 | 0.681 | 0.802 | 674 | 1220 |
| SCC | -0.094 | 0.048 | -0.188 | 0.000 | 0.050 | 0.520 | 679 | 1214 |
| SCR | -0.029 | 0.048 | -0.123 | 0.065 | 0.549 | 0.802 | 675 | 1215 |
| SFO | -0.018 | 0.048 | -0.113 | 0.076 | 0.702 | 0.802 | 673 | 1211 |
| SLF | -0.078 | 0.048 | -0.172 | 0.016 | 0.104 | 0.520 | 676 | 1217 |
| SS | 0.056 | 0.048 | -0.038 | 0.150 | 0.242 | 0.581 | 677 | 1216 |
| UNC | 0.083 | 0.049 | -0.013 | 0.179 | 0.089 | 0.520 | 661 | 1152 |
| AverageAD | -0.038 | 0.048 | -0.133 | 0.056 | 0.425 | 0.753 | 674 | 1214 |

Table S14. Differences in regional FA between individuals with recent suicidal ideation and healthy controls.
Age, sex, age-by-sex, age$^2$, age$^2$-by-sex included as covariates.

| Region | Cohen's d | SE | CI LB | CI UB | P-value | FDR P-value | Ideation | HC |
|---|---|---|---|---|---|---|---|---|
| ACR | -0.112 | 0.047 | -0.204 | -0.020 | 0.018 | 0.064 | 712 | 1236 |
| ALIC | -0.095 | 0.047 | -0.187 | -0.003 | 0.044 | 0.104 | 712 | 1235 |
| BCC | -0.087 | 0.047 | -0.179 | 0.006 | 0.066 | 0.128 | 709 | 1232 |
| CC | -0.108 | 0.047 | -0.201 | -0.016 | 0.022 | 0.068 | 710 | 1233 |
| CGC | -0.019 | 0.047 | -0.111 | 0.074 | 0.691 | 0.742 | 711 | 1233 |
| CGH | -0.074 | 0.047 | -0.166 | 0.018 | 0.117 | 0.195 | 711 | 1234 |
| CR | -0.133 | 0.047 | -0.225 | -0.041 | 0.005 | 0.031 | 710 | 1233 |
| CST | -0.029 | 0.047 | -0.122 | 0.063 | 0.537 | 0.610 | 710 | 1223 |
| EC | -0.093 | 0.047 | -0.186 | -0.001 | 0.048 | 0.104 | 713 | 1230 |
| FX | -0.015 | 0.047 | -0.108 | 0.077 | 0.746 | 0.746 | 709 | 1230 |
| FXST | -0.070 | 0.047 | -0.162 | 0.023 | 0.141 | 0.207 | 711 | 1234 |
| GCC | -0.148 | 0.047 | -0.241 | -0.056 | 0.002 | 0.017 | 711 | 1233 |
| IC | -0.070 | 0.047 | -0.162 | 0.023 | 0.140 | 0.207 | 710 | 1235 |
| IFO | 0.017 | 0.047 | -0.075 | 0.110 | 0.713 | 0.742 | 710 | 1226 |
| PCR | -0.118 | 0.047 | -0.210 | -0.025 | 0.013 | 0.063 | 710 | 1234 |
| PLIC | -0.036 | 0.047 | -0.129 | 0.056 | 0.442 | 0.526 | 706 | 1234 |
| PTR | -0.079 | 0.047 | -0.172 | 0.013 | 0.093 | 0.166 | 710 | 1235 |
| RLIC | -0.052 | 0.047 | -0.145 | 0.040 | 0.266 | 0.350 | 712 | 1232 |
| SCC | -0.042 | 0.047 | -0.135 | 0.050 | 0.371 | 0.464 | 711 | 1230 |
| SCR | -0.112 | 0.047 | -0.204 | -0.019 | 0.018 | 0.064 | 709 | 1234 |
| SFO | -0.145 | 0.047 | -0.238 | -0.053 | 0.002 | 0.017 | 709 | 1232 |
| SLF | -0.097 | 0.047 | -0.190 | -0.005 | 0.039 | 0.104 | 712 | 1233 |
| SS | -0.093 | 0.047 | -0.185 | 0.000 | 0.050 | 0.104 | 710 | 1230 |
| UNC | -0.061 | 0.048 | -0.154 | 0.033 | 0.207 | 0.287 | 696 | 1169 |
| AverageFA | -0.158 | 0.048 | -0.251 | -0.064 | 0.001 | 0.017 | 685 | 1231 |

Table S15. Differences in regional MD between individuals with recent suicidal ideation and healthy controls. Age, sex, age-by-sex, age$^2$, age$^2$-by-sex included as covariates.

| Region | Cohen's d | SE | CI LB | CI UB | P-value | FDR P-value | Ideation | HC |
|---|---|---|---|---|---|---|---|---|
| ACR | 0.029 | 0.047 | -0.063 | 0.122 | 0.536 | 0.706 | 703 | 1217 |
| ALIC | 0.053 | 0.048 | -0.040 | 0.147 | 0.262 | 0.409 | 699 | 1204 |
| BCC | 0.059 | 0.047 | -0.033 | 0.152 | 0.211 | 0.389 | 702 | 1213 |
| CC | 0.032 | 0.047 | -0.061 | 0.125 | 0.499 | 0.693 | 703 | 1214 |
| CGC | -0.008 | 0.047 | -0.101 | 0.085 | 0.867 | 0.867 | 700 | 1214 |
| CGH | 0.089 | 0.047 | -0.004 | 0.182 | 0.061 | 0.256 | 702 | 1219 |
| CR | 0.070 | 0.047 | -0.023 | 0.163 | 0.143 | 0.324 | 701 | 1216 |
| CST | -0.008 | 0.048 | -0.101 | 0.085 | 0.863 | 0.867 | 698 | 1203 |
| EC | 0.082 | 0.048 | -0.011 | 0.176 | 0.084 | 0.278 | 697 | 1209 |
| FX | -0.017 | 0.047 | -0.110 | 0.076 | 0.720 | 0.805 | 700 | 1209 |
| FXST | 0.103 | 0.047 | 0.010 | 0.196 | 0.030 | 0.189 | 703 | 1215 |
| GCC | 0.017 | 0.047 | -0.076 | 0.110 | 0.722 | 0.805 | 704 | 1216 |
| IC | 0.043 | 0.047 | -0.050 | 0.136 | 0.370 | 0.544 | 701 | 1212 |
| IFO | 0.078 | 0.047 | -0.016 | 0.171 | 0.103 | 0.287 | 700 | 1213 |
| PCR | 0.081 | 0.047 | -0.012 | 0.174 | 0.089 | 0.278 | 700 | 1217 |
| PLIC | -0.016 | 0.047 | -0.109 | 0.077 | 0.740 | 0.805 | 699 | 1215 |
| PTR | 0.094 | 0.047 | 0.001 | 0.187 | 0.048 | 0.241 | 701 | 1215 |
| RLIC | 0.060 | 0.047 | -0.033 | 0.153 | 0.206 | 0.389 | 703 | 1214 |
| SCC | -0.018 | 0.047 | -0.111 | 0.075 | 0.703 | 0.805 | 701 | 1212 |
| SCR | 0.121 | 0.047 | 0.028 | 0.214 | 0.011 | 0.138 | 703 | 1214 |
| SFO | 0.059 | 0.048 | -0.034 | 0.152 | 0.218 | 0.389 | 699 | 1212 |
| SLF | 0.054 | 0.047 | -0.039 | 0.147 | 0.259 | 0.409 | 701 | 1216 |
| SS | 0.071 | 0.047 | -0.022 | 0.164 | 0.136 | 0.324 | 700 | 1219 |
| UNC | 0.135 | 0.048 | 0.040 | 0.230 | 0.005 | 0.132 | 684 | 1145 |
| AverageMD | 0.109 | 0.048 | 0.015 | 0.203 | 0.023 | 0.189 | 675 | 1212 |

Table S16. Differences in regional RD between individuals with recent suicidal ideation and healthy controls.
Age, sex, age-by-sex, $age^2$, $age^2$-by-sex included as covariates.

| Region | Cohen's d | SE | CI LB | CI UB | P-value | FDR P-value | Ideation | HC |
|---|---|---|---|---|---|---|---|---|
| ACR | 0.086 | 0.048 | -0.008 | 0.180 | 0.073 | 0.142 | 674 | 1214 |
| ALIC | 0.080 | 0.048 | -0.014 | 0.174 | 0.097 | 0.158 | 674 | 1206 |
| BCC | 0.079 | 0.048 | -0.015 | 0.173 | 0.101 | 0.158 | 673 | 1211 |
| CC | 0.068 | 0.048 | -0.026 | 0.162 | 0.158 | 0.227 | 674 | 1214 |
| CGC | -0.006 | 0.048 | -0.100 | 0.088 | 0.900 | 0.938 | 673 | 1213 |
| CGH | 0.088 | 0.048 | -0.006 | 0.182 | 0.066 | 0.142 | 675 | 1221 |
| CR | 0.114 | 0.048 | 0.020 | 0.208 | 0.018 | 0.112 | 674 | 1215 |
| CST | 0.035 | 0.048 | -0.059 | 0.130 | 0.464 | 0.504 | 673 | 1202 |
| EC | 0.099 | 0.048 | 0.005 | 0.193 | 0.040 | 0.125 | 674 | 1213 |
| FX | 0.002 | 0.048 | -0.092 | 0.097 | 0.963 | 0.963 | 674 | 1206 |
| FXST | 0.092 | 0.048 | -0.002 | 0.186 | 0.056 | 0.142 | 676 | 1215 |
| GCC | 0.067 | 0.048 | -0.027 | 0.161 | 0.163 | 0.227 | 677 | 1215 |
| IC | 0.084 | 0.048 | -0.010 | 0.179 | 0.080 | 0.142 | 674 | 1213 |
| IFO | 0.045 | 0.048 | -0.049 | 0.139 | 0.352 | 0.400 | 674 | 1211 |
| PCR | 0.101 | 0.048 | 0.007 | 0.195 | 0.036 | 0.125 | 675 | 1213 |
| PLIC | 0.046 | 0.048 | -0.048 | 0.140 | 0.340 | 0.400 | 671 | 1215 |
| PTR | 0.105 | 0.048 | 0.010 | 0.199 | 0.030 | 0.125 | 676 | 1214 |
| RLIC | 0.057 | 0.048 | -0.037 | 0.151 | 0.238 | 0.313 | 676 | 1215 |
| SCC | 0.050 | 0.048 | -0.044 | 0.145 | 0.296 | 0.370 | 675 | 1207 |
| SCR | 0.150 | 0.048 | 0.056 | 0.244 | 0.002 | 0.040 | 677 | 1215 |
| SFO | 0.088 | 0.048 | -0.006 | 0.182 | 0.068 | 0.142 | 673 | 1209 |
| SLF | 0.100 | 0.048 | 0.006 | 0.194 | 0.038 | 0.125 | 678 | 1214 |
| SS | 0.085 | 0.048 | -0.009 | 0.179 | 0.078 | 0.142 | 676 | 1219 |
| UNC | 0.122 | 0.049 | 0.026 | 0.218 | 0.013 | 0.107 | 659 | 1143 |
| AverageRD | 0.142 | 0.048 | 0.048 | 0.236 | 0.003 | 0.040 | 677 | 1214 |

Table S17. Differences in regional AD between individuals with recent suicidal ideation and clinical controls. Age, sex, age-by-sex, age$^2$, age$^2$-by-sex included as covariates.

| Region | Cohen's d | SE | CI LB | CI UB | P-value | FDR P-value | Ideation | CLC |
|---|---|---|---|---|---|---|---|---|
| ACR | 0.069 | 0.049 | -0.026 | 0.164 | 0.156 | 0.862 | 677 | 1136 |
| ALIC | 0.007 | 0.049 | -0.088 | 0.102 | 0.885 | 0.976 | 675 | 1139 |
| BCC | -0.016 | 0.049 | -0.111 | 0.079 | 0.746 | 0.938 | 675 | 1138 |
| CC | -0.001 | 0.049 | -0.097 | 0.094 | 0.976 | 0.976 | 678 | 1137 |
| CGC | 0.126 | 0.049 | 0.031 | 0.221 | 0.010 | 0.135 | 677 | 1137 |
| CGH | 0.006 | 0.049 | -0.089 | 0.102 | 0.898 | 0.976 | 673 | 1138 |
| CR | 0.040 | 0.049 | -0.055 | 0.135 | 0.414 | 0.938 | 676 | 1136 |
| CST | -0.003 | 0.049 | -0.098 | 0.093 | 0.959 | 0.976 | 674 | 1136 |
| EC | 0.063 | 0.049 | -0.033 | 0.158 | 0.199 | 0.862 | 669 | 1134 |
| FX | -0.025 | 0.049 | -0.121 | 0.070 | 0.601 | 0.938 | 676 | 1130 |
| FXST | 0.031 | 0.049 | -0.064 | 0.126 | 0.529 | 0.938 | 677 | 1140 |
| GCC | 0.039 | 0.049 | -0.057 | 0.134 | 0.427 | 0.938 | 677 | 1136 |
| IC | 0.018 | 0.049 | -0.077 | 0.113 | 0.709 | 0.938 | 674 | 1139 |
| IFO | 0.088 | 0.049 | -0.008 | 0.183 | 0.072 | 0.599 | 676 | 1139 |
| PCR | 0.019 | 0.049 | -0.076 | 0.114 | 0.699 | 0.938 | 675 | 1134 |
| PLIC | 0.015 | 0.049 | -0.081 | 0.110 | 0.762 | 0.938 | 673 | 1139 |
| PTR | 0.054 | 0.049 | -0.041 | 0.150 | 0.265 | 0.938 | 674 | 1137 |
| RLIC | 0.061 | 0.049 | -0.034 | 0.157 | 0.207 | 0.862 | 674 | 1137 |
| SCC | -0.017 | 0.049 | -0.112 | 0.078 | 0.723 | 0.938 | 679 | 1137 |
| SCR | 0.013 | 0.049 | -0.082 | 0.108 | 0.788 | 0.938 | 675 | 1136 |
| SFO | -0.020 | 0.049 | -0.115 | 0.075 | 0.683 | 0.938 | 673 | 1132 |
| SLF | 0.040 | 0.049 | -0.055 | 0.135 | 0.411 | 0.938 | 676 | 1134 |
| SS | 0.019 | 0.049 | -0.076 | 0.115 | 0.689 | 0.938 | 677 | 1134 |
| UNC | 0.027 | 0.049 | -0.069 | 0.124 | 0.579 | 0.938 | 661 | 1088 |
| AverageAD | 0.124 | 0.049 | 0.029 | 0.220 | 0.011 | 0.135 | 674 | 1134 |

Table S18. Differences in regional FA between individuals with recent suicidal ideation and clinical controls.
Age, sex, age-by-sex, $age^2$, $age^2$-by-sex included as covariates.

| Region | Cohen's d | SE | CI LB | CI UB | P-value | FDR P-value | Ideation | CLC |
|---|---|---|---|---|---|---|---|---|
| ACR | -0.048 | 0.047 | -0.141 | 0.045 | 0.312 | 0.576 | 712 | 1182 |
| ALIC | -0.026 | 0.047 | -0.119 | 0.067 | 0.582 | 0.766 | 712 | 1177 |
| BCC | 0.013 | 0.048 | -0.080 | 0.106 | 0.786 | 0.854 | 709 | 1174 |
| CC | 0.009 | 0.048 | -0.084 | 0.102 | 0.847 | 0.883 | 710 | 1175 |
| CGC | 0.038 | 0.048 | -0.055 | 0.131 | 0.422 | 0.660 | 711 | 1177 |
| CGH | -0.065 | 0.048 | -0.159 | 0.028 | 0.169 | 0.576 | 711 | 1176 |
| CR | -0.065 | 0.048 | -0.158 | 0.028 | 0.173 | 0.576 | 710 | 1179 |
| CST | -0.005 | 0.048 | -0.099 | 0.088 | 0.910 | 0.910 | 710 | 1176 |
| EC | -0.034 | 0.047 | -0.127 | 0.059 | 0.477 | 0.663 | 713 | 1175 |
| FX | 0.048 | 0.048 | -0.045 | 0.141 | 0.316 | 0.576 | 709 | 1177 |
| FXST | -0.064 | 0.048 | -0.157 | 0.029 | 0.181 | 0.576 | 711 | 1176 |
| GCC | -0.014 | 0.047 | -0.107 | 0.080 | 0.775 | 0.854 | 711 | 1177 |
| IC | -0.053 | 0.048 | -0.146 | 0.040 | 0.268 | 0.576 | 710 | 1178 |
| IFO | 0.059 | 0.047 | -0.034 | 0.152 | 0.215 | 0.576 | 710 | 1182 |
| PCR | -0.058 | 0.048 | -0.151 | 0.035 | 0.223 | 0.576 | 710 | 1179 |
| PLIC | -0.015 | 0.048 | -0.109 | 0.078 | 0.749 | 0.854 | 706 | 1176 |
| PTR | -0.065 | 0.048 | -0.158 | 0.028 | 0.172 | 0.576 | 710 | 1179 |
| RLIC | -0.077 | 0.047 | -0.170 | 0.016 | 0.104 | 0.576 | 712 | 1178 |
| SCC | 0.047 | 0.048 | -0.047 | 0.140 | 0.328 | 0.576 | 711 | 1176 |
| SCR | -0.054 | 0.048 | -0.147 | 0.039 | 0.255 | 0.576 | 709 | 1180 |
| SFO | -0.101 | 0.048 | -0.195 | -0.008 | 0.033 | 0.576 | 709 | 1180 |
| SLF | -0.023 | 0.047 | -0.116 | 0.070 | 0.632 | 0.790 | 712 | 1183 |
| SS | -0.087 | 0.048 | -0.180 | 0.006 | 0.067 | 0.576 | 710 | 1180 |
| UNC | -0.035 | 0.048 | -0.130 | 0.059 | 0.467 | 0.663 | 696 | 1131 |
| AverageFA | -0.046 | 0.048 | -0.140 | 0.049 | 0.345 | 0.576 | 685 | 1140 |

Table S19. Differences in regional MD between individuals with recent suicidal ideation and clinical controls. Age, sex, age-by-sex, $age^2$, $age^2$-by-sex included as covariates.

| Region | Cohen's d | SE | CI LB | CI UB | P-value | FDR P-value | Ideation | CLC |
|---|---|---|---|---|---|---|---|---|
| ACR | 0.056 | 0.048 | -0.037 | 0.150 | 0.240 | 0.659 | 703 | 1175 |
| ALIC | 0.047 | 0.048 | -0.046 | 0.141 | 0.324 | 0.659 | 699 | 1173 |
| BCC | -0.012 | 0.048 | -0.105 | 0.082 | 0.805 | 0.839 | 702 | 1174 |
| CC | -0.022 | 0.048 | -0.116 | 0.071 | 0.643 | 0.765 | 703 | 1178 |
| CGC | 0.092 | 0.048 | -0.002 | 0.186 | 0.054 | 0.659 | 700 | 1177 |
| CGH | 0.028 | 0.048 | -0.066 | 0.121 | 0.560 | 0.719 | 702 | 1176 |
| CR | 0.057 | 0.048 | -0.036 | 0.151 | 0.231 | 0.659 | 701 | 1176 |
| CST | -0.015 | 0.048 | -0.109 | 0.079 | 0.752 | 0.818 | 698 | 1170 |
| EC | 0.062 | 0.048 | -0.032 | 0.156 | 0.197 | 0.659 | 697 | 1174 |
| FX | -0.036 | 0.048 | -0.129 | 0.058 | 0.459 | 0.719 | 700 | 1162 |
| FXST | 0.053 | 0.048 | -0.040 | 0.147 | 0.264 | 0.659 | 703 | 1174 |
| GCC | 0.017 | 0.048 | -0.077 | 0.110 | 0.725 | 0.818 | 704 | 1177 |
| IC | 0.048 | 0.048 | -0.046 | 0.142 | 0.315 | 0.659 | 701 | 1173 |
| IFO | 0.006 | 0.048 | -0.087 | 0.100 | 0.896 | 0.896 | 700 | 1175 |
| PCR | 0.033 | 0.048 | -0.060 | 0.127 | 0.485 | 0.719 | 700 | 1175 |
| PLIC | 0.030 | 0.048 | -0.064 | 0.124 | 0.530 | 0.719 | 699 | 1175 |
| PTR | 0.045 | 0.048 | -0.048 | 0.139 | 0.343 | 0.659 | 701 | 1176 |
| RLIC | 0.069 | 0.048 | -0.025 | 0.162 | 0.149 | 0.659 | 703 | 1177 |
| SCC | -0.050 | 0.048 | -0.143 | 0.044 | 0.300 | 0.659 | 701 | 1174 |
| SCR | 0.076 | 0.048 | -0.018 | 0.169 | 0.113 | 0.659 | 703 | 1173 |
| SFO | 0.034 | 0.048 | -0.059 | 0.128 | 0.473 | 0.719 | 699 | 1171 |
| SLF | 0.059 | 0.048 | -0.035 | 0.152 | 0.221 | 0.659 | 701 | 1174 |
| SS | 0.032 | 0.048 | -0.062 | 0.125 | 0.509 | 0.719 | 700 | 1171 |
| UNC | 0.027 | 0.048 | -0.068 | 0.122 | 0.575 | 0.719 | 684 | 1127 |
| AverageMD | 0.090 | 0.049 | -0.006 | 0.185 | 0.066 | 0.659 | 675 | 1136 |

Table S20. Differences in regional RD between individuals with recent suicidal ideation and clinical controls. Age, sex, age-by-sex, age$^2$, age$^2$-by-sex included as covariates.

| Region | Cohen's d | SE | CI LB | CI UB | P-value | FDR P-value | Ideation | CLC |
|---|---|---|---|---|---|---|---|---|
| ACR | 0.080 | 0.049 | -0.015 | 0.176 | 0.099 | 0.248 | 674 | 1135 |
| ALIC | 0.064 | 0.049 | -0.031 | 0.160 | 0.187 | 0.334 | 674 | 1133 |
| BCC | -0.012 | 0.049 | -0.108 | 0.083 | 0.801 | 0.870 | 673 | 1137 |
| CC | -0.006 | 0.049 | -0.101 | 0.089 | 0.901 | 0.939 | 674 | 1135 |
| CGC | 0.034 | 0.049 | -0.061 | 0.130 | 0.479 | 0.631 | 673 | 1136 |
| CGH | 0.047 | 0.049 | -0.048 | 0.143 | 0.330 | 0.485 | 675 | 1135 |
| CR | 0.081 | 0.049 | -0.015 | 0.176 | 0.098 | 0.248 | 674 | 1135 |
| CST | -0.003 | 0.049 | -0.099 | 0.092 | 0.946 | 0.946 | 673 | 1137 |
| EC | 0.074 | 0.049 | -0.021 | 0.170 | 0.128 | 0.267 | 674 | 1132 |
| FX | -0.038 | 0.049 | -0.133 | 0.058 | 0.438 | 0.608 | 674 | 1122 |
| FXST | 0.089 | 0.049 | -0.006 | 0.184 | 0.067 | 0.248 | 676 | 1136 |
| GCC | 0.025 | 0.049 | -0.070 | 0.120 | 0.604 | 0.755 | 677 | 1134 |
| IC | 0.096 | 0.049 | 0.001 | 0.191 | 0.049 | 0.248 | 674 | 1136 |
| IFO | -0.018 | 0.049 | -0.113 | 0.077 | 0.710 | 0.807 | 674 | 1133 |
| PCR | 0.061 | 0.049 | -0.034 | 0.156 | 0.210 | 0.338 | 675 | 1136 |
| PLIC | 0.077 | 0.049 | -0.019 | 0.172 | 0.115 | 0.261 | 671 | 1135 |
| PTR | 0.066 | 0.049 | -0.030 | 0.161 | 0.178 | 0.334 | 676 | 1136 |
| RLIC | 0.085 | 0.049 | -0.010 | 0.180 | 0.081 | 0.248 | 676 | 1137 |
| SCC | -0.022 | 0.049 | -0.118 | 0.073 | 0.647 | 0.771 | 675 | 1133 |
| SCR | 0.095 | 0.049 | 0.000 | 0.190 | 0.051 | 0.248 | 677 | 1134 |
| SFO | 0.082 | 0.049 | -0.013 | 0.178 | 0.092 | 0.248 | 673 | 1135 |
| SLF | 0.081 | 0.049 | -0.014 | 0.176 | 0.095 | 0.248 | 678 | 1136 |
| SS | 0.099 | 0.049 | 0.004 | 0.194 | 0.042 | 0.248 | 676 | 1132 |
| UNC | 0.061 | 0.049 | -0.036 | 0.158 | 0.216 | 0.338 | 659 | 1087 |
| AverageRD | 0.083 | 0.049 | -0.012 | 0.178 | 0.088 | 0.248 | 677 | 1136 |

**CSSRS analyses**

Table S21. Association between lifetime intensity of ideation (scored 0-5) in the Columbia Suicide Severity Rating Scale and regional AD (healthy controls were excluded). Beta: standardized beta, p: p-value, FDR-p: FDR corrected p-value.

| Region | Beta | Lower CI | Upper CI | T-value | P | FDR-p | N |
|---|---|---|---|---|---|---|---|
| ACR | -0.070 | -0.186 | 0.047 | -1.174 | 0.241 | 0.367 | 299 |
| ALIC | -0.080 | -0.197 | 0.037 | -1.338 | 0.182 | 0.325 | 299 |
| BCC | -0.122 | -0.240 | -0.005 | -2.050 | 0.041 | 0.206 | 299 |
| CC | -0.109 | -0.227 | 0.009 | -1.825 | 0.069 | 0.288 | 299 |
| CGC | 0.076 | -0.040 | 0.193 | 1.287 | 0.199 | 0.332 | 299 |
| CGH | -0.063 | -0.177 | 0.052 | -1.076 | 0.283 | 0.393 | 299 |
| CR | -0.080 | -0.197 | 0.038 | -1.338 | 0.182 | 0.325 | 299 |
| CST | -0.102 | -0.219 | 0.015 | -1.723 | 0.086 | 0.298 | 297 |
| EC | -0.041 | -0.159 | 0.077 | -0.690 | 0.491 | 0.548 | 297 |
| FX | -0.117 | -0.228 | -0.006 | -2.081 | 0.038 | 0.206 | 298 |
| FXST | -0.058 | -0.170 | 0.055 | -1.002 | 0.317 | 0.396 | 299 |
| GCC | -0.093 | -0.211 | 0.024 | -1.563 | 0.119 | 0.298 | 297 |
| IC | -0.118 | -0.229 | -0.006 | -2.077 | 0.039 | 0.206 | 297 |
| IFO | -0.093 | -0.209 | 0.024 | -1.565 | 0.119 | 0.298 | 298 |
| PCR | -0.040 | -0.158 | 0.078 | -0.666 | 0.506 | 0.548 | 299 |
| PLIC | -0.126 | -0.235 | -0.017 | -2.285 | 0.023 | 0.206 | 298 |
| PTR | -0.052 | -0.170 | 0.066 | -0.860 | 0.390 | 0.465 | 299 |
| RLIC | -0.092 | -0.205 | 0.020 | -1.612 | 0.108 | 0.298 | 299 |
| SCC | -0.062 | -0.179 | 0.056 | -1.031 | 0.303 | 0.396 | 298 |
| SCR | -0.082 | -0.199 | 0.036 | -1.371 | 0.171 | 0.325 | 299 |
| SFO | -0.089 | -0.207 | 0.029 | -1.489 | 0.137 | 0.312 | 298 |
| SLF | -0.069 | -0.187 | 0.049 | -1.154 | 0.250 | 0.367 | 299 |
| SS | -0.036 | -0.154 | 0.081 | -0.608 | 0.544 | 0.548 | 298 |
| UNC | -0.035 | -0.151 | 0.081 | -0.601 | 0.548 | 0.548 | 299 |
| AverageAD | -0.127 | -0.245 | -0.010 | -2.137 | 0.033 | 0.206 | 298 |

Table S22. Association between lifetime intensity of ideation (scored 0-5) in the Columbia Suicide Severity Rating Scale and regional FA (healthy controls were excluded). Beta: standardized beta, p: p-value, FDR-p: FDR corrected p-value.

| Region | Beta | Lower CI | Upper CI | T-value | P | FDR-p | N |
|---|---|---|---|---|---|---|---|
| ACR | -0.047 | -0.151 | 0.057 | -0.894 | 0.372 | 0.990 | 298 |
| ALIC | -0.028 | -0.144 | 0.088 | -0.474 | 0.636 | 0.990 | 298 |
| BCC | -0.010 | -0.126 | 0.107 | -0.160 | 0.873 | 0.990 | 298 |
| CC | -0.038 | -0.153 | 0.076 | -0.662 | 0.508 | 0.990 | 298 |
| CGC | 0.002 | -0.113 | 0.117 | 0.040 | 0.968 | 0.990 | 299 |
| CGH | -0.004 | -0.122 | 0.113 | -0.071 | 0.943 | 0.990 | 299 |
| CR | -0.005 | -0.111 | 0.100 | -0.102 | 0.919 | 0.990 | 298 |
| CST | 0.008 | -0.110 | 0.126 | 0.137 | 0.891 | 0.990 | 298 |
| EC | -0.028 | -0.138 | 0.082 | -0.496 | 0.620 | 0.990 | 298 |
| FX | 0.056 | -0.052 | 0.163 | 1.022 | 0.308 | 0.990 | 296 |
| FXST | -0.079 | -0.191 | 0.032 | -1.402 | 0.162 | 0.990 | 297 |
| GCC | -0.052 | -0.162 | 0.058 | -0.931 | 0.353 | 0.990 | 298 |
| IC | -0.027 | -0.141 | 0.087 | -0.470 | 0.638 | 0.990 | 298 |
| IFO | -0.104 | -0.219 | 0.011 | -1.779 | 0.076 | 0.953 | 298 |
| PCR | 0.014 | -0.098 | 0.126 | 0.249 | 0.803 | 0.990 | 299 |
| PLIC | 0.013 | -0.106 | 0.131 | 0.209 | 0.835 | 0.990 | 296 |
| PTR | -0.053 | -0.158 | 0.052 | -0.995 | 0.321 | 0.990 | 297 |
| RLIC | -0.028 | -0.138 | 0.081 | -0.514 | 0.608 | 0.990 | 298 |
| SCC | -0.056 | -0.173 | 0.061 | -0.946 | 0.345 | 0.990 | 295 |
| SCR | 0.032 | -0.080 | 0.143 | 0.561 | 0.575 | 0.990 | 299 |
| SFO | -0.135 | -0.251 | -0.020 | -2.311 | 0.022 | 0.538 | 297 |
| SLF | -0.009 | -0.121 | 0.103 | -0.160 | 0.873 | 0.990 | 299 |
| SS | 0.027 | -0.081 | 0.135 | 0.490 | 0.625 | 0.990 | 297 |
| UNC | -0.001 | -0.115 | 0.114 | -0.012 | 0.990 | 0.990 | 299 |
| AverageFA | -0.014 | -0.125 | 0.096 | -0.258 | 0.797 | 0.990 | 298 |

Table S23. Association between lifetime intensity of ideation (scored 0-5) in the Columbia Suicide Severity Rating Scale and regional MD (healthy controls were excluded). Beta: standardized beta, p: p-value, FDR-p: FDR corrected p-value.

| Region | Beta | Lower CI | Upper CI | T-value | P | FDR-p | N |
|---|---|---|---|---|---|---|---|
| ACR | -0.013 | -0.126 | 0.101 | -0.219 | 0.827 | 0.862 | 299 |
| ALIC | -0.005 | -0.123 | 0.113 | -0.085 | 0.933 | 0.933 | 298 |
| BCC | -0.073 | -0.190 | 0.044 | -1.234 | 0.218 | 0.683 | 298 |
| CC | -0.058 | -0.175 | 0.059 | -0.982 | 0.327 | 0.683 | 299 |
| CGC | 0.023 | -0.094 | 0.141 | 0.391 | 0.696 | 0.857 | 299 |
| CGH | -0.064 | -0.180 | 0.051 | -1.096 | 0.274 | 0.683 | 299 |
| CR | -0.040 | -0.155 | 0.076 | -0.678 | 0.498 | 0.779 | 299 |
| CST | -0.085 | -0.201 | 0.032 | -1.430 | 0.154 | 0.683 | 297 |
| EC | -0.021 | -0.135 | 0.092 | -0.367 | 0.714 | 0.857 | 297 |
| FX | -0.139 | -0.244 | -0.035 | -2.620 | 0.009 | 0.232 | 294 |
| FXST | 0.015 | -0.104 | 0.133 | 0.246 | 0.806 | 0.862 | 298 |
| GCC | -0.029 | -0.144 | 0.087 | -0.488 | 0.626 | 0.857 | 299 |
| IC | -0.067 | -0.185 | 0.050 | -1.126 | 0.261 | 0.683 | 297 |
| IFO | 0.014 | -0.103 | 0.131 | 0.231 | 0.818 | 0.862 | 298 |
| PCR | -0.040 | -0.158 | 0.077 | -0.678 | 0.499 | 0.779 | 299 |
| PLIC | -0.089 | -0.201 | 0.024 | -1.555 | 0.121 | 0.683 | 297 |
| PTR | -0.058 | -0.175 | 0.059 | -0.980 | 0.328 | 0.683 | 296 |
| RLIC | -0.080 | -0.199 | 0.038 | -1.337 | 0.182 | 0.683 | 298 |
| SCC | -0.022 | -0.140 | 0.097 | -0.361 | 0.719 | 0.857 | 298 |
| SCR | -0.070 | -0.187 | 0.047 | -1.177 | 0.240 | 0.683 | 299 |
| SFO | 0.022 | -0.096 | 0.139 | 0.359 | 0.720 | 0.857 | 298 |
| SLF | -0.079 | -0.196 | 0.038 | -1.337 | 0.182 | 0.683 | 298 |
| SS | -0.041 | -0.157 | 0.076 | -0.686 | 0.493 | 0.779 | 298 |
| UNC | -0.053 | -0.169 | 0.064 | -0.893 | 0.373 | 0.717 | 298 |
| AverageMD | -0.073 | -0.190 | 0.045 | -1.217 | 0.225 | 0.683 | 297 |

Table S24. Association between lifetime intensity of ideation (scored 0-5) in the Columbia Suicide Severity Rating Scale and regional RD (healthy controls were excluded). Beta: standardized beta, p: p-value, FDR-p: FDR corrected p-value.

| Region | Beta | Lower CI | Upper CI | T-value | P | FDR-p | N |
|---|---|---|---|---|---|---|---|
| ACR | 0.008 | -0.100 | 0.116 | 0.143 | 0.887 | 0.964 | 298 |
| ALIC | 0.000 | -0.117 | 0.116 | -0.006 | 0.995 | 0.995 | 297 |
| BCC | -0.033 | -0.151 | 0.084 | -0.554 | 0.580 | 0.926 | 298 |
| CC | -0.002 | -0.119 | 0.115 | -0.038 | 0.970 | 0.995 | 298 |
| CGC | -0.039 | -0.155 | 0.076 | -0.673 | 0.502 | 0.926 | 298 |
| CGH | -0.042 | -0.159 | 0.074 | -0.717 | 0.474 | 0.926 | 299 |
| CR | -0.016 | -0.126 | 0.095 | -0.282 | 0.778 | 0.926 | 298 |
| CST | -0.071 | -0.188 | 0.046 | -1.190 | 0.235 | 0.926 | 297 |
| EC | -0.024 | -0.135 | 0.087 | -0.431 | 0.667 | 0.926 | 297 |
| FX | -0.152 | -0.256 | -0.048 | -2.877 | 0.004 | 0.108 | 293 |
| FXST | 0.042 | -0.075 | 0.158 | 0.704 | 0.482 | 0.926 | 299 |
| GCC | 0.024 | -0.091 | 0.139 | 0.413 | 0.680 | 0.926 | 298 |
| IC | -0.017 | -0.134 | 0.100 | -0.284 | 0.777 | 0.926 | 298 |
| IFO | 0.067 | -0.049 | 0.182 | 1.136 | 0.257 | 0.926 | 298 |
| PCR | -0.023 | -0.138 | 0.093 | -0.385 | 0.701 | 0.926 | 299 |
| PLIC | -0.055 | -0.173 | 0.062 | -0.923 | 0.357 | 0.926 | 296 |
| PTR | -0.024 | -0.138 | 0.089 | -0.426 | 0.671 | 0.926 | 296 |
| RLIC | -0.018 | -0.134 | 0.097 | -0.315 | 0.753 | 0.926 | 298 |
| SCC | 0.014 | -0.105 | 0.133 | 0.230 | 0.819 | 0.930 | 297 |
| SCR | -0.040 | -0.153 | 0.073 | -0.701 | 0.484 | 0.926 | 298 |
| SFO | 0.061 | -0.056 | 0.178 | 1.027 | 0.305 | 0.926 | 299 |
| SLF | -0.034 | -0.148 | 0.080 | -0.589 | 0.556 | 0.926 | 299 |
| SS | -0.046 | -0.158 | 0.067 | -0.801 | 0.424 | 0.926 | 297 |
| UNC | -0.046 | -0.161 | 0.069 | -0.792 | 0.429 | 0.926 | 299 |
| AverageRD | -0.052 | -0.167 | 0.063 | -0.894 | 0.372 | 0.926 | 298 |

Table S25. Association between recent intensity of ideation (scored 0-5) in the Columbia Suicide Severity Rating Scale and regional AD (healthy controls were excluded). Beta: standardized beta, p: p-value, FDR-p: FDR corrected p-value.

| Region | Beta | Lower CI | Upper CI | T-value | P | FDR-p | N |
|---|---|---|---|---|---|---|---|
| ACR | -0.077 | -0.188 | 0.034 | -1.359 | 0.175 | 0.273 | 338 |
| ALIC | -0.078 | -0.189 | 0.033 | -1.376 | 0.170 | 0.273 | 338 |
| BCC | -0.101 | -0.213 | 0.011 | -1.780 | 0.076 | 0.190 | 337 |
| CC | -0.086 | -0.198 | 0.026 | -1.507 | 0.133 | 0.255 | 337 |
| CGC | -0.067 | -0.179 | 0.045 | -1.182 | 0.238 | 0.313 | 338 |
| CGH | 0.014 | -0.097 | 0.124 | 0.241 | 0.810 | 0.843 | 338 |
| CR | -0.119 | -0.231 | -0.008 | -2.110 | 0.036 | 0.149 | 338 |
| CST | -0.036 | -0.149 | 0.076 | -0.637 | 0.525 | 0.596 | 336 |
| EC | -0.089 | -0.201 | 0.023 | -1.561 | 0.120 | 0.255 | 335 |
| FX | -0.052 | -0.161 | 0.056 | -0.946 | 0.345 | 0.410 | 337 |
| FXST | -0.082 | -0.192 | 0.029 | -1.454 | 0.147 | 0.263 | 337 |
| GCC | -0.022 | -0.134 | 0.091 | -0.379 | 0.705 | 0.766 | 336 |
| IC | -0.147 | -0.256 | -0.039 | -2.665 | 0.008 | 0.083 | 336 |
| IFO | -0.105 | -0.216 | 0.006 | -1.865 | 0.063 | 0.175 | 338 |
| PCR | -0.087 | -0.199 | 0.025 | -1.531 | 0.127 | 0.255 | 338 |
| PLIC | -0.144 | -0.249 | -0.038 | -2.683 | 0.008 | 0.083 | 337 |
| PTR | -0.068 | -0.181 | 0.045 | -1.185 | 0.237 | 0.313 | 337 |
| RLIC | -0.145 | -0.254 | -0.035 | -2.585 | 0.010 | 0.083 | 337 |
| SCC | -0.074 | -0.186 | 0.038 | -1.304 | 0.193 | 0.284 | 336 |
| SCR | -0.141 | -0.252 | -0.030 | -2.488 | 0.013 | 0.083 | 338 |
| SFO | -0.117 | -0.229 | -0.005 | -2.056 | 0.041 | 0.149 | 337 |
| SLF | -0.112 | -0.224 | -0.001 | -1.981 | 0.048 | 0.151 | 338 |
| SS | -0.057 | -0.169 | 0.056 | -0.986 | 0.325 | 0.406 | 336 |
| UNC | 0.008 | -0.104 | 0.120 | 0.139 | 0.890 | 0.890 | 338 |
| AverageAD | -0.116 | -0.227 | -0.004 | -2.043 | 0.042 | 0.149 | 338 |

Table S26. Association between recent intensity of ideation (scored 0-5) in the Columbia Suicide Severity Rating Scale and regional FA (healthy controls were excluded). Beta: standardized beta, p: p-value, FDR-p: FDR corrected p-value.

| Region | Beta | Lower CI | Upper CI | T-value | P | FDR-p | N |
|---|---|---|---|---|---|---|---|
| ACR | 0.026 | -0.078 | 0.130 | 0.490 | 0.625 | 0.855 | 336 |
| ALIC | 0.047 | -0.063 | 0.157 | 0.842 | 0.401 | 0.855 | 335 |
| BCC | -0.021 | -0.133 | 0.091 | -0.372 | 0.710 | 0.855 | 334 |
| CC | -0.040 | -0.149 | 0.070 | -0.711 | 0.477 | 0.855 | 335 |
| CGC | -0.017 | -0.127 | 0.093 | -0.306 | 0.759 | 0.855 | 338 |
| CGH | 0.013 | -0.099 | 0.126 | 0.233 | 0.816 | 0.855 | 338 |
| CR | 0.019 | -0.087 | 0.124 | 0.347 | 0.729 | 0.855 | 336 |
| CST | 0.052 | -0.060 | 0.164 | 0.914 | 0.361 | 0.855 | 337 |
| EC | -0.052 | -0.160 | 0.057 | -0.936 | 0.350 | 0.855 | 337 |
| FX | -0.080 | -0.189 | 0.029 | -1.450 | 0.148 | 0.855 | 335 |
| FXST | -0.013 | -0.124 | 0.098 | -0.233 | 0.816 | 0.855 | 334 |
| GCC | -0.040 | -0.144 | 0.065 | -0.749 | 0.455 | 0.855 | 335 |
| IC | -0.013 | -0.122 | 0.097 | -0.227 | 0.821 | 0.855 | 336 |
| IFO | -0.163 | -0.272 | -0.054 | -2.933 | 0.004 | 0.090 | 337 |
| PCR | 0.022 | -0.088 | 0.132 | 0.399 | 0.690 | 0.855 | 338 |
| PLIC | 0.014 | -0.098 | 0.126 | 0.242 | 0.809 | 0.855 | 336 |
| PTR | -0.020 | -0.127 | 0.087 | -0.366 | 0.715 | 0.855 | 336 |
| RLIC | -0.048 | -0.156 | 0.061 | -0.867 | 0.386 | 0.855 | 336 |
| SCC | -0.047 | -0.159 | 0.065 | -0.825 | 0.410 | 0.855 | 333 |
| SCR | -0.008 | -0.117 | 0.101 | -0.143 | 0.886 | 0.886 | 338 |
| SFO | -0.050 | -0.162 | 0.062 | -0.878 | 0.381 | 0.855 | 336 |
| SLF | 0.036 | -0.073 | 0.145 | 0.645 | 0.519 | 0.855 | 338 |
| SS | 0.038 | -0.070 | 0.146 | 0.691 | 0.490 | 0.855 | 335 |
| UNC | 0.056 | -0.052 | 0.165 | 1.018 | 0.309 | 0.855 | 338 |
| AverageFA | 0.035 | -0.073 | 0.144 | 0.640 | 0.522 | 0.855 | 336 |

Table S27. Association between recent intensity of ideation (scored 0-5) in the Columbia Suicide Severity Rating Scale and regional MD (healthy controls were excluded). Beta: standardized beta, p: p-value, FDR-p: FDR corrected p-value.

| Region | Beta | Lower CI | Upper CI | T-value | P | FDR-p | N |
|---|---|---|---|---|---|---|---|
| ACR | -0.068 | -0.177 | 0.040 | -1.242 | 0.215 | 0.358 | 337 |
| ALIC | -0.088 | -0.200 | 0.023 | -1.562 | 0.119 | 0.271 | 337 |
| BCC | -0.055 | -0.166 | 0.057 | -0.959 | 0.338 | 0.448 | 336 |
| CC | -0.060 | -0.171 | 0.051 | -1.058 | 0.291 | 0.428 | 338 |
| CGC | -0.097 | -0.208 | 0.014 | -1.715 | 0.087 | 0.260 | 337 |
| CGH | -0.003 | -0.114 | 0.109 | -0.052 | 0.959 | 0.959 | 338 |
| CR | -0.104 | -0.214 | 0.005 | -1.872 | 0.062 | 0.222 | 337 |
| CST | -0.052 | -0.164 | 0.060 | -0.916 | 0.360 | 0.451 | 334 |
| EC | -0.053 | -0.162 | 0.056 | -0.954 | 0.341 | 0.448 | 337 |
| FX | -0.020 | -0.126 | 0.086 | -0.371 | 0.711 | 0.808 | 334 |
| FXST | -0.094 | -0.207 | 0.018 | -1.650 | 0.100 | 0.260 | 337 |
| GCC | -0.009 | -0.117 | 0.100 | -0.156 | 0.876 | 0.913 | 338 |
| IC | -0.164 | -0.275 | -0.053 | -2.897 | 0.004 | 0.087 | 336 |
| IFO | 0.017 | -0.095 | 0.128 | 0.294 | 0.769 | 0.836 | 337 |
| PCR | -0.093 | -0.205 | 0.019 | -1.630 | 0.104 | 0.260 | 337 |
| PLIC | -0.149 | -0.257 | -0.041 | -2.714 | 0.007 | 0.087 | 336 |
| PTR | -0.074 | -0.186 | 0.038 | -1.307 | 0.192 | 0.343 | 334 |
| RLIC | -0.140 | -0.251 | -0.029 | -2.482 | 0.014 | 0.101 | 336 |
| SCC | -0.063 | -0.175 | 0.050 | -1.093 | 0.275 | 0.428 | 336 |
| SCR | -0.135 | -0.246 | -0.024 | -2.395 | 0.017 | 0.101 | 337 |
| SFO | -0.076 | -0.188 | 0.037 | -1.323 | 0.187 | 0.343 | 337 |
| SLF | -0.132 | -0.243 | -0.021 | -2.333 | 0.020 | 0.101 | 336 |
| SS | -0.080 | -0.191 | 0.031 | -1.415 | 0.158 | 0.329 | 336 |
| UNC | -0.035 | -0.147 | 0.077 | -0.612 | 0.541 | 0.644 | 338 |
| AverageMD | -0.111 | -0.222 | 0.000 | -1.967 | 0.050 | 0.208 | 337 |

Table S28. Association between recent intensity of ideation (scored 0-5) in the Columbia Suicide Severity Rating Scale and regional RD (healthy controls were excluded). Beta: standardized beta, p: p-value, FDR-p: FDR corrected p-value.

| Region | Beta | Lower CI | Upper CI | T-value | P | FDR-p | N |
|---|---|---|---|---|---|---|---|
| ACR | -0.060 | -0.166 | 0.045 | -1.125 | 0.261 | 0.670 | 334 |
| ALIC | -0.068 | -0.179 | 0.043 | -1.201 | 0.231 | 0.670 | 336 |
| BCC | -0.009 | -0.121 | 0.103 | -0.157 | 0.875 | 0.916 | 336 |
| CC | 0.009 | -0.103 | 0.120 | 0.152 | 0.879 | 0.916 | 335 |
| CGC | -0.049 | -0.159 | 0.060 | -0.885 | 0.377 | 0.685 | 336 |
| CGH | -0.006 | -0.118 | 0.107 | -0.100 | 0.920 | 0.920 | 338 |
| CR | -0.062 | -0.169 | 0.045 | -1.143 | 0.254 | 0.670 | 334 |
| CST | -0.014 | -0.126 | 0.098 | -0.248 | 0.804 | 0.916 | 334 |
| EC | -0.018 | -0.125 | 0.090 | -0.327 | 0.744 | 0.916 | 336 |
| FX | -0.009 | -0.113 | 0.096 | -0.160 | 0.873 | 0.916 | 334 |
| FXST | -0.080 | -0.192 | 0.031 | -1.417 | 0.157 | 0.670 | 338 |
| GCC | 0.030 | -0.077 | 0.136 | 0.548 | 0.584 | 0.913 | 335 |
| IC | -0.058 | -0.169 | 0.053 | -1.033 | 0.302 | 0.670 | 336 |
| IFO | 0.115 | 0.005 | 0.225 | 2.051 | 0.041 | 0.670 | 337 |
| PCR | -0.060 | -0.172 | 0.051 | -1.062 | 0.289 | 0.670 | 336 |
| PLIC | -0.056 | -0.168 | 0.055 | -0.993 | 0.322 | 0.670 | 335 |
| PTR | -0.015 | -0.126 | 0.095 | -0.268 | 0.789 | 0.916 | 333 |
| RLIC | -0.042 | -0.153 | 0.069 | -0.740 | 0.460 | 0.766 | 336 |
| SCC | 0.022 | -0.091 | 0.134 | 0.376 | 0.707 | 0.916 | 335 |
| SCR | -0.073 | -0.182 | 0.036 | -1.314 | 0.190 | 0.670 | 336 |
| SFO | -0.025 | -0.138 | 0.088 | -0.435 | 0.664 | 0.916 | 338 |
| SLF | -0.092 | -0.202 | 0.017 | -1.659 | 0.098 | 0.670 | 337 |
| SS | -0.071 | -0.180 | 0.039 | -1.271 | 0.205 | 0.670 | 334 |
| UNC | -0.049 | -0.159 | 0.061 | -0.873 | 0.383 | 0.685 | 338 |
| AverageRD | -0.080 | -0.189 | 0.030 | -1.435 | 0.152 | 0.670 | 337 |

Table S29. Differences in regional AD between people with a lifetime history of an actual suicide attempt and those without a history of any suicide attempt determined using the Columbia Suicide Severity Rating Scale (healthy controls were excluded).
D: Cohen's d effect size, SE: standard error; p: p-value, FDR-p: FDR corrected p-value, CI: confidence interval, HC: healthy controls, CC: clinical controls.

| Region | D | SE | P | FDR-p | Lower CI | Upper CI | N actual attempt | N no attempt |
|---|---|---|---|---|---|---|---|---|
| ACR | -0.100 | 0.108 | 0.357 | 0.733 | -0.311 | 0.111 | 134 | 242 |
| ALIC | -0.102 | 0.108 | 0.347 | 0.733 | -0.314 | 0.109 | 134 | 241 |
| BCC | -0.244 | 0.108 | 0.026 | 0.177 | -0.456 | -0.032 | 134 | 240 |
| CC | -0.256 | 0.108 | 0.019 | 0.177 | -0.468 | -0.044 | 134 | 239 |
| CGC | 0.025 | 0.108 | 0.821 | 0.933 | -0.186 | 0.235 | 134 | 243 |
| CGH | 0.056 | 0.108 | 0.605 | 0.756 | -0.155 | 0.267 | 134 | 244 |
| CR | -0.081 | 0.108 | 0.459 | 0.733 | -0.292 | 0.131 | 134 | 240 |
| CST | 0.007 | 0.108 | 0.951 | 0.988 | -0.205 | 0.218 | 133 | 242 |
| EC | -0.117 | 0.108 | 0.285 | 0.713 | -0.329 | 0.095 | 133 | 240 |
| FX | 0.032 | 0.108 | 0.769 | 0.915 | -0.179 | 0.243 | 134 | 244 |
| FXST | -0.068 | 0.108 | 0.529 | 0.735 | -0.280 | 0.143 | 134 | 242 |
| GCC | -0.236 | 0.108 | 0.031 | 0.177 | -0.448 | -0.024 | 133 | 244 |
| IC | -0.137 | 0.108 | 0.211 | 0.658 | -0.349 | 0.075 | 133 | 239 |
| IFO | -0.085 | 0.108 | 0.433 | 0.733 | -0.296 | 0.126 | 134 | 242 |
| PCR | -0.008 | 0.108 | 0.942 | 0.988 | -0.219 | 0.203 | 134 | 242 |
| PLIC | -0.078 | 0.108 | 0.474 | 0.733 | -0.290 | 0.133 | 134 | 240 |
| PTR | -0.161 | 0.108 | 0.141 | 0.504 | -0.373 | 0.051 | 134 | 239 |
| RLIC | -0.230 | 0.108 | 0.035 | 0.177 | -0.443 | -0.018 | 134 | 238 |
| SCC | -0.120 | 0.108 | 0.271 | 0.713 | -0.332 | 0.091 | 134 | 239 |
| SCR | -0.074 | 0.108 | 0.499 | 0.733 | -0.286 | 0.138 | 134 | 238 |
| SFO | -0.002 | 0.108 | 0.988 | 0.988 | -0.213 | 0.209 | 134 | 242 |
| SLF | -0.260 | 0.108 | 0.017 | 0.177 | -0.473 | -0.048 | 134 | 240 |
| SS | -0.060 | 0.108 | 0.582 | 0.756 | -0.271 | 0.151 | 134 | 242 |
| UNC | 0.078 | 0.108 | 0.472 | 0.733 | -0.133 | 0.289 | 134 | 242 |
| AverageAD | -0.210 | 0.108 | 0.055 | 0.228 | -0.422 | 0.002 | 134 | 239 |

Table S30. Differences in regional FA between people with a lifetime history of an actual suicide attempt and those without a history of any suicide attempt determined using the Columbia Suicide Severity Rating Scale (healthy controls were excluded).
D: Cohen's d effect size, SE: standard error; p: p-value, FDR-p: FDR corrected p-value, CI: confidence interval, HC: healthy controls, CC: clinical controls.

| Region | D | SE | P | FDR-p | Lower CI | Upper CI | N actual attempt | N no attempt |
|---|---|---|---|---|---|---|---|---|
| ACR | 0.045 | 0.108 | 0.678 | 0.983 | -0.167 | 0.257 | 132 | 243 |
| ALIC | -0.022 | 0.108 | 0.838 | 0.983 | -0.234 | 0.189 | 133 | 241 |
| BCC | -0.002 | 0.108 | 0.983 | 0.983 | -0.215 | 0.210 | 131 | 244 |
| CC | -0.034 | 0.108 | 0.757 | 0.983 | -0.246 | 0.178 | 132 | 242 |
| CGC | -0.129 | 0.108 | 0.235 | 0.983 | -0.340 | 0.082 | 134 | 244 |
| CGH | -0.065 | 0.108 | 0.549 | 0.983 | -0.276 | 0.146 | 134 | 242 |
| CR | 0.045 | 0.108 | 0.679 | 0.983 | -0.167 | 0.257 | 132 | 242 |
| CST | 0.078 | 0.108 | 0.471 | 0.983 | -0.133 | 0.290 | 134 | 242 |
| EC | 0.005 | 0.108 | 0.964 | 0.983 | -0.207 | 0.216 | 133 | 243 |
| FX | -0.009 | 0.108 | 0.933 | 0.983 | -0.221 | 0.203 | 132 | 242 |
| FXST | -0.132 | 0.108 | 0.227 | 0.983 | -0.345 | 0.080 | 132 | 241 |
| GCC | 0.039 | 0.108 | 0.719 | 0.983 | -0.173 | 0.251 | 132 | 243 |
| IC | -0.006 | 0.108 | 0.957 | 0.983 | -0.218 | 0.206 | 133 | 241 |
| IFO | 0.012 | 0.108 | 0.911 | 0.983 | -0.199 | 0.223 | 134 | 243 |
| PCR | -0.049 | 0.108 | 0.653 | 0.983 | -0.260 | 0.162 | 134 | 244 |
| PLIC | 0.103 | 0.108 | 0.344 | 0.983 | -0.108 | 0.315 | 133 | 242 |
| PTR | -0.127 | 0.108 | 0.246 | 0.983 | -0.339 | 0.085 | 132 | 242 |
| RLIC | -0.062 | 0.108 | 0.571 | 0.983 | -0.274 | 0.150 | 133 | 241 |
| SCC | -0.093 | 0.109 | 0.397 | 0.983 | -0.307 | 0.121 | 130 | 237 |
| SCR | 0.065 | 0.108 | 0.547 | 0.983 | -0.145 | 0.276 | 134 | 244 |
| SFO | -0.113 | 0.108 | 0.301 | 0.983 | -0.325 | 0.099 | 133 | 242 |
| SLF | -0.122 | 0.108 | 0.262 | 0.983 | -0.333 | 0.089 | 134 | 244 |
| SS | 0.051 | 0.108 | 0.643 | 0.983 | -0.162 | 0.263 | 132 | 239 |
| UNC | -0.096 | 0.108 | 0.376 | 0.983 | -0.307 | 0.115 | 134 | 243 |
| AverageFA | -0.022 | 0.108 | 0.837 | 0.983 | -0.235 | 0.190 | 132 | 241 |

Table S31. Differences in regional MD between people with a lifetime history of an actual suicide attempt and those without a history of any suicide attempt determined using the Columbia Suicide Severity Rating Scale (healthy controls were excluded).
D: Cohen's d effect size, SE: standard error; p: p-value, FDR-p: FDR corrected p-value, CI: confidence interval, HC: healthy controls, CC: clinical controls.

| Region | D | SE | P | FDR-p | Lower CI | Upper CI | N actual attempt | N no attempt |
|---|---|---|---|---|---|---|---|---|
| ACR | -0.033 | 0.108 | 0.760 | 0.966 | -0.244 | 0.178 | 134 | 242 |
| ALIC | 0.015 | 0.108 | 0.888 | 0.966 | -0.196 | 0.227 | 133 | 244 |
| BCC | -0.063 | 0.108 | 0.563 | 0.966 | -0.275 | 0.149 | 132 | 244 |
| CC | -0.092 | 0.108 | 0.399 | 0.966 | -0.302 | 0.119 | 134 | 244 |
| CGC | -0.015 | 0.108 | 0.889 | 0.966 | -0.226 | 0.196 | 134 | 243 |
| CGH | 0.024 | 0.108 | 0.827 | 0.966 | -0.187 | 0.235 | 134 | 243 |
| CR | 0.006 | 0.108 | 0.957 | 0.997 | -0.205 | 0.217 | 134 | 243 |
| CST | -0.040 | 0.108 | 0.712 | 0.966 | -0.252 | 0.172 | 134 | 237 |
| EC | -0.090 | 0.108 | 0.409 | 0.966 | -0.302 | 0.121 | 133 | 243 |
| FX | -0.027 | 0.108 | 0.806 | 0.966 | -0.239 | 0.185 | 132 | 243 |
| FXST | 0.051 | 0.108 | 0.643 | 0.966 | -0.161 | 0.262 | 133 | 244 |
| GCC | -0.203 | 0.108 | 0.063 | 0.966 | -0.414 | 0.009 | 134 | 244 |
| IC | -0.070 | 0.108 | 0.522 | 0.966 | -0.281 | 0.142 | 133 | 243 |
| IFO | -0.109 | 0.108 | 0.316 | 0.966 | -0.321 | 0.102 | 133 | 244 |
| PCR | 0.038 | 0.108 | 0.725 | 0.966 | -0.173 | 0.249 | 134 | 243 |
| PLIC | -0.065 | 0.108 | 0.548 | 0.966 | -0.277 | 0.146 | 133 | 243 |
| PTR | -0.085 | 0.108 | 0.437 | 0.966 | -0.296 | 0.127 | 133 | 242 |
| RLIC | -0.150 | 0.108 | 0.171 | 0.966 | -0.361 | 0.062 | 133 | 242 |
| SCC | -0.020 | 0.108 | 0.857 | 0.966 | -0.231 | 0.192 | 133 | 242 |
| SCR | 0.000 | 0.108 | 0.997 | 0.997 | -0.212 | 0.211 | 134 | 242 |
| SFO | 0.090 | 0.108 | 0.408 | 0.966 | -0.121 | 0.301 | 134 | 243 |
| SLF | -0.077 | 0.108 | 0.480 | 0.966 | -0.288 | 0.134 | 134 | 242 |
| SS | -0.141 | 0.108 | 0.195 | 0.966 | -0.353 | 0.070 | 133 | 243 |
| UNC | 0.126 | 0.108 | 0.250 | 0.966 | -0.086 | 0.338 | 132 | 244 |
| AverageMD | -0.086 | 0.108 | 0.432 | 0.966 | -0.297 | 0.126 | 133 | 243 |

Table S32. Differences in regional RD between people with a lifetime history of an actual suicide attempt and those without a history of any suicide attempt determined using the Columbia Suicide Severity Rating Scale (healthy controls were excluded).
D: Cohen's d effect size, SE: standard error; p: p-value, FDR-p: FDR corrected p-value, CI: confidence interval, HC: healthy controls, CC: clinical controls.

| Region | D | SE | P | FDR-p | Lower CI | Upper CI | N actual attempt | N no attempt |
|---|---|---|---|---|---|---|---|---|
| ACR | -0.066 | 0.108 | 0.548 | 0.833 | -0.278 | 0.147 | 132 | 241 |
| ALIC | -0.084 | 0.108 | 0.443 | 0.833 | -0.296 | 0.128 | 132 | 244 |
| BCC | -0.028 | 0.108 | 0.794 | 0.870 | -0.240 | 0.183 | 132 | 244 |
| CC | -0.047 | 0.108 | 0.665 | 0.833 | -0.259 | 0.165 | 132 | 244 |
| CGC | -0.078 | 0.108 | 0.474 | 0.833 | -0.289 | 0.133 | 133 | 244 |
| CGH | -0.001 | 0.108 | 0.991 | 0.991 | -0.212 | 0.210 | 134 | 243 |
| CR | -0.011 | 0.108 | 0.919 | 0.957 | -0.223 | 0.201 | 133 | 241 |
| CST | -0.058 | 0.108 | 0.593 | 0.833 | -0.270 | 0.153 | 133 | 240 |
| EC | -0.127 | 0.108 | 0.244 | 0.833 | -0.339 | 0.084 | 133 | 243 |
| FX | -0.063 | 0.108 | 0.563 | 0.833 | -0.276 | 0.149 | 131 | 243 |
| FXST | 0.059 | 0.108 | 0.588 | 0.833 | -0.152 | 0.270 | 134 | 244 |
| GCC | -0.104 | 0.108 | 0.340 | 0.833 | -0.316 | 0.108 | 132 | 243 |
| IC | -0.102 | 0.108 | 0.348 | 0.833 | -0.314 | 0.109 | 133 | 243 |
| IFO | -0.109 | 0.108 | 0.316 | 0.833 | -0.320 | 0.102 | 134 | 244 |
| PCR | 0.047 | 0.108 | 0.667 | 0.833 | -0.164 | 0.258 | 134 | 242 |
| PLIC | -0.125 | 0.108 | 0.252 | 0.833 | -0.337 | 0.087 | 133 | 241 |
| PTR | -0.049 | 0.108 | 0.651 | 0.833 | -0.262 | 0.163 | 132 | 241 |
| RLIC | -0.071 | 0.108 | 0.513 | 0.833 | -0.283 | 0.140 | 133 | 242 |
| SCC | 0.028 | 0.108 | 0.798 | 0.870 | -0.184 | 0.240 | 132 | 242 |
| SCR | -0.058 | 0.108 | 0.597 | 0.833 | -0.269 | 0.154 | 133 | 243 |
| SFO | 0.089 | 0.108 | 0.412 | 0.833 | -0.122 | 0.300 | 134 | 244 |
| SLF | 0.028 | 0.108 | 0.800 | 0.870 | -0.183 | 0.238 | 134 | 243 |
| SS | -0.186 | 0.108 | 0.090 | 0.833 | -0.398 | 0.027 | 132 | 242 |
| UNC | 0.074 | 0.108 | 0.494 | 0.833 | -0.136 | 0.285 | 134 | 244 |
| AverageRD | -0.099 | 0.108 | 0.362 | 0.833 | -0.311 | 0.112 | 133 | 244 |

Table S33. Differences in regional AD between people with a lifetime history of an actual suicide and those without a history of an actual suicide attempt but with lifetime suicidal ideation determined with the Columbia Suicide Severity Rating Scale (healthy controls were excluded). D: Cohen's d effect size, SE: standard error; p: p-value, FDR-p: FDR corrected p-value, CI: confidence interval.

| Region | D | SE | P | FDR-p | Lower CI | Upper CI | N ideation | N attempt |
|---|---|---|---|---|---|---|---|---|
| ACR | -0.254 | 0.126 | 0.047 | 0.343 | -0.500 | -0.007 | 122 | 134 |
| ALIC | -0.098 | 0.125 | 0.439 | 0.610 | -0.344 | 0.147 | 122 | 134 |
| BCC | -0.171 | 0.125 | 0.179 | 0.448 | -0.417 | 0.075 | 122 | 134 |
| CC | -0.178 | 0.125 | 0.161 | 0.448 | -0.424 | 0.067 | 122 | 134 |
| CGC | -0.100 | 0.125 | 0.430 | 0.610 | -0.346 | 0.145 | 122 | 134 |
| CGH | 0.034 | 0.125 | 0.792 | 0.825 | -0.212 | 0.279 | 122 | 134 |
| CR | -0.175 | 0.125 | 0.168 | 0.448 | -0.421 | 0.070 | 122 | 134 |
| CST | 0.120 | 0.126 | 0.345 | 0.576 | -0.126 | 0.367 | 121 | 133 |
| EC | -0.126 | 0.126 | 0.324 | 0.576 | -0.372 | 0.121 | 121 | 133 |
| FX | 0.175 | 0.126 | 0.169 | 0.448 | -0.071 | 0.422 | 121 | 134 |
| FXST | -0.173 | 0.125 | 0.175 | 0.448 | -0.418 | 0.073 | 122 | 134 |
| GCC | -0.273 | 0.126 | 0.033 | 0.343 | -0.521 | -0.026 | 121 | 133 |
| IC | -0.037 | 0.126 | 0.773 | 0.825 | -0.283 | 0.209 | 121 | 133 |
| IFO | -0.123 | 0.126 | 0.334 | 0.576 | -0.369 | 0.123 | 121 | 134 |
| PCR | -0.152 | 0.125 | 0.232 | 0.511 | -0.398 | 0.094 | 122 | 134 |
| PLIC | -0.005 | 0.125 | 0.971 | 0.971 | -0.250 | 0.241 | 121 | 134 |
| PTR | -0.232 | 0.126 | 0.069 | 0.343 | -0.478 | 0.014 | 122 | 134 |
| RLIC | -0.106 | 0.125 | 0.404 | 0.610 | -0.352 | 0.139 | 122 | 134 |
| SCC | -0.041 | 0.125 | 0.747 | 0.825 | -0.287 | 0.205 | 121 | 134 |
| SCR | -0.045 | 0.125 | 0.725 | 0.825 | -0.290 | 0.201 | 122 | 134 |
| SFO | -0.053 | 0.125 | 0.678 | 0.825 | -0.299 | 0.193 | 121 | 134 |
| SLF | -0.346 | 0.126 | 0.007 | 0.173 | -0.593 | -0.098 | 122 | 134 |
| SS | -0.148 | 0.126 | 0.245 | 0.511 | -0.394 | 0.098 | 121 | 134 |
| UNC | 0.081 | 0.125 | 0.522 | 0.686 | -0.164 | 0.327 | 122 | 134 |
| AverageAD | -0.244 | 0.126 | 0.056 | 0.343 | -0.491 | 0.002 | 121 | 134 |

Table S34. Differences in regional FA between people with a lifetime history of an actual suicide and those without a history of an actual suicide attempt but with lifetime suicidal ideation determined with the Columbia Suicide Severity Rating Scale (healthy controls were excluded). D: Cohen's d effect size, SE: standard error; p: p-value, FDR-p: FDR corrected p-value, CI: confidence interval.

| Region | D | SE | P | FDR-p | Lower CI | Upper CI | N ideation | N attempt |
|---|---|---|---|---|---|---|---|---|
| ACR | 0.113 | 0.126 | 0.378 | 0.865 | -0.134 | 0.359 | 122 | 132 |
| ALIC | 0.075 | 0.125 | 0.558 | 0.865 | -0.171 | 0.320 | 122 | 133 |
| BCC | 0.045 | 0.126 | 0.727 | 0.865 | -0.202 | 0.291 | 122 | 131 |
| CC | 0.037 | 0.126 | 0.772 | 0.877 | -0.209 | 0.283 | 122 | 132 |
| CGC | -0.098 | 0.125 | 0.440 | 0.865 | -0.344 | 0.147 | 122 | 134 |
| CGH | -0.095 | 0.125 | 0.453 | 0.865 | -0.341 | 0.150 | 122 | 134 |
| CR | 0.110 | 0.126 | 0.389 | 0.865 | -0.136 | 0.356 | 122 | 132 |
| CST | 0.117 | 0.126 | 0.359 | 0.865 | -0.129 | 0.363 | 121 | 134 |
| EC | 0.046 | 0.125 | 0.715 | 0.865 | -0.199 | 0.292 | 122 | 133 |
| FX | -0.078 | 0.126 | 0.541 | 0.865 | -0.324 | 0.168 | 122 | 132 |
| FXST | -0.123 | 0.126 | 0.334 | 0.865 | -0.370 | 0.123 | 122 | 132 |
| GCC | 0.051 | 0.126 | 0.689 | 0.865 | -0.195 | 0.297 | 122 | 132 |
| IC | 0.099 | 0.125 | 0.436 | 0.865 | -0.147 | 0.345 | 122 | 133 |
| IFO | -0.070 | 0.125 | 0.580 | 0.865 | -0.316 | 0.175 | 121 | 134 |
| PCR | -0.022 | 0.125 | 0.865 | 0.901 | -0.267 | 0.224 | 122 | 134 |
| PLIC | 0.137 | 0.126 | 0.285 | 0.865 | -0.110 | 0.383 | 121 | 133 |
| PTR | -0.182 | 0.126 | 0.155 | 0.865 | -0.428 | 0.065 | 122 | 132 |
| RLIC | 0.008 | 0.125 | 0.947 | 0.947 | -0.237 | 0.254 | 122 | 133 |
| SCC | 0.045 | 0.126 | 0.725 | 0.865 | -0.202 | 0.292 | 122 | 130 |
| SCR | 0.102 | 0.125 | 0.423 | 0.865 | -0.144 | 0.347 | 122 | 134 |
| SFO | 0.022 | 0.126 | 0.862 | 0.901 | -0.224 | 0.268 | 121 | 133 |
| SLF | -0.125 | 0.125 | 0.326 | 0.865 | -0.370 | 0.121 | 122 | 134 |
| SS | 0.139 | 0.126 | 0.275 | 0.865 | -0.107 | 0.386 | 122 | 132 |
| UNC | -0.068 | 0.125 | 0.594 | 0.865 | -0.313 | 0.178 | 122 | 134 |
| AverageFA | 0.062 | 0.126 | 0.628 | 0.865 | -0.184 | 0.308 | 122 | 132 |

Table S35. Differences in regional MD between people with a lifetime history of an actual suicide and those without a history of an actual suicide attempt but with lifetime suicidal ideation determined with the Columbia Suicide Severity Rating Scale (healthy controls were excluded). D: Cohen's d effect size, SE: standard error; p: p-value, FDR-p: FDR corrected p-value, CI: confidence interval.

| Region | D | SE | P | FDR-p | Lower CI | Upper CI | N ideation | N attempt |
|---|---|---|---|---|---|---|---|---|
| ACR | -0.218 | 0.126 | 0.088 | 0.476 | -0.464 | 0.028 | 122 | 134 |
| ALIC | -0.081 | 0.125 | 0.526 | 0.722 | -0.327 | 0.165 | 122 | 133 |
| BCC | -0.097 | 0.126 | 0.447 | 0.684 | -0.343 | 0.149 | 122 | 132 |
| CC | -0.118 | 0.125 | 0.353 | 0.684 | -0.364 | 0.127 | 122 | 134 |
| CGC | -0.168 | 0.125 | 0.186 | 0.476 | -0.414 | 0.077 | 122 | 134 |
| CGH | 0.066 | 0.125 | 0.603 | 0.722 | -0.179 | 0.311 | 122 | 134 |
| CR | -0.169 | 0.125 | 0.185 | 0.476 | -0.414 | 0.077 | 122 | 134 |
| CST | -0.011 | 0.126 | 0.931 | 0.948 | -0.257 | 0.235 | 120 | 134 |
| EC | -0.198 | 0.126 | 0.122 | 0.476 | -0.444 | 0.049 | 121 | 133 |
| FX | 0.154 | 0.126 | 0.231 | 0.524 | -0.094 | 0.401 | 120 | 132 |
| FXST | -0.039 | 0.125 | 0.758 | 0.824 | -0.285 | 0.207 | 122 | 133 |
| GCC | -0.183 | 0.125 | 0.151 | 0.476 | -0.428 | 0.063 | 122 | 134 |
| IC | -0.102 | 0.126 | 0.426 | 0.684 | -0.348 | 0.145 | 121 | 133 |
| IFO | -0.074 | 0.125 | 0.562 | 0.722 | -0.320 | 0.172 | 122 | 133 |
| PCR | -0.110 | 0.125 | 0.387 | 0.684 | -0.355 | 0.136 | 122 | 134 |
| PLIC | -0.065 | 0.126 | 0.609 | 0.722 | -0.312 | 0.181 | 121 | 133 |
| PTR | -0.174 | 0.126 | 0.174 | 0.476 | -0.420 | 0.073 | 121 | 133 |
| RLIC | -0.181 | 0.126 | 0.155 | 0.476 | -0.427 | 0.065 | 122 | 133 |
| SCC | -0.060 | 0.125 | 0.636 | 0.722 | -0.306 | 0.185 | 122 | 133 |
| SCR | -0.097 | 0.125 | 0.444 | 0.684 | -0.343 | 0.148 | 122 | 134 |
| SFO | -0.008 | 0.125 | 0.948 | 0.948 | -0.253 | 0.237 | 122 | 134 |
| SLF | -0.175 | 0.125 | 0.170 | 0.476 | -0.421 | 0.071 | 122 | 134 |
| SS | -0.218 | 0.126 | 0.088 | 0.476 | -0.464 | 0.029 | 122 | 133 |
| UNC | 0.093 | 0.126 | 0.465 | 0.684 | -0.153 | 0.339 | 122 | 132 |
| AverageMD | -0.167 | 0.126 | 0.191 | 0.476 | -0.414 | 0.079 | 121 | 133 |

Table S36. Differences in regional RD between people with a lifetime history of an actual suicide and those without a history of an actual suicide attempt but with lifetime suicidal ideation determined with the Columbia Suicide Severity Rating Scale (healthy controls were excluded). D: Cohen's d effect size, SE: standard error; p: p-value, FDR-p: FDR corrected p-value, CI: confidence interval.

| Region | D | SE | P | FDR-p | Lower CI | Upper CI | N ideation | N attempt |
|---|---|---|---|---|---|---|---|---|
| ACR | -0.204 | 0.126 | 0.111 | 0.684 | -0.450 | 0.043 | 122 | 132 |
| ALIC | -0.137 | 0.126 | 0.284 | 0.684 | -0.383 | 0.110 | 122 | 132 |
| BCC | -0.067 | 0.126 | 0.600 | 0.770 | -0.313 | 0.179 | 122 | 132 |
| CC | -0.089 | 0.126 | 0.487 | 0.770 | -0.335 | 0.158 | 122 | 132 |
| CGC | -0.127 | 0.125 | 0.319 | 0.684 | -0.373 | 0.119 | 122 | 133 |
| CGH | 0.052 | 0.125 | 0.684 | 0.770 | -0.194 | 0.297 | 122 | 134 |
| CR | -0.145 | 0.126 | 0.255 | 0.684 | -0.391 | 0.101 | 122 | 133 |
| CST | -0.072 | 0.126 | 0.574 | 0.770 | -0.318 | 0.175 | 121 | 133 |
| EC | -0.163 | 0.126 | 0.200 | 0.684 | -0.409 | 0.083 | 122 | 133 |
| FX | 0.125 | 0.126 | 0.331 | 0.684 | -0.123 | 0.373 | 120 | 131 |
| FXST | 0.044 | 0.125 | 0.730 | 0.770 | -0.202 | 0.289 | 122 | 134 |
| GCC | -0.121 | 0.126 | 0.341 | 0.684 | -0.368 | 0.125 | 122 | 132 |
| IC | -0.166 | 0.126 | 0.192 | 0.684 | -0.413 | 0.080 | 122 | 133 |
| IFO | -0.049 | 0.125 | 0.703 | 0.770 | -0.294 | 0.197 | 121 | 134 |
| PCR | -0.052 | 0.125 | 0.682 | 0.770 | -0.297 | 0.193 | 122 | 134 |
| PLIC | -0.158 | 0.126 | 0.217 | 0.684 | -0.404 | 0.089 | 121 | 133 |
| PTR | -0.079 | 0.126 | 0.536 | 0.770 | -0.326 | 0.168 | 121 | 132 |
| RLIC | -0.118 | 0.125 | 0.356 | 0.684 | -0.364 | 0.128 | 122 | 133 |
| SCC | -0.050 | 0.126 | 0.694 | 0.770 | -0.296 | 0.196 | 122 | 132 |
| SCR | -0.138 | 0.126 | 0.278 | 0.684 | -0.384 | 0.108 | 122 | 133 |
| SFO | 0.042 | 0.125 | 0.739 | 0.770 | -0.203 | 0.288 | 122 | 134 |
| SLF | -0.032 | 0.125 | 0.804 | 0.804 | -0.277 | 0.214 | 122 | 134 |
| SS | -0.248 | 0.126 | 0.052 | 0.684 | -0.495 | -0.001 | 122 | 132 |
| UNC | 0.083 | 0.125 | 0.511 | 0.770 | -0.162 | 0.329 | 122 | 134 |
| AverageRD | -0.133 | 0.125 | 0.298 | 0.684 | -0.379 | 0.113 | 122 | 133 |

Table S37. Differences in regional AD between people with a lifetime history an interrupted, aborted or actual attempt and those without a history of an interrupted, aborted or actual suicide attempt determined with the Columbia Suicide Severity Rating Scale (healthy controls were excluded). D: Cohen's d effect size, SE: standard error; p: p-value, FDR-p: FDR corrected p-value, CI: confidence interval.

| Region | D | SE | P | FDR-p | N attempt | N no attempt |
|---|---|---|---|---|---|---|
| ACR | -0.074 | 0.099 | 0.457 | 0.722 | 177 | 242 |
| ALIC | -0.132 | 0.099 | 0.187 | 0.451 | 177 | 241 |
| BCC | -0.245 | 0.099 | 0.015 | 0.147 | 177 | 240 |
| CC | -0.235 | 0.100 | 0.019 | 0.147 | 177 | 239 |
| CGC | 0.007 | 0.099 | 0.942 | 0.942 | 177 | 243 |
| CGH | 0.056 | 0.099 | 0.578 | 0.753 | 177 | 244 |
| CR | -0.088 | 0.099 | 0.377 | 0.674 | 177 | 240 |
| CST | -0.064 | 0.099 | 0.520 | 0.722 | 176 | 242 |
| EC | -0.144 | 0.100 | 0.151 | 0.420 | 175 | 240 |
| FX | -0.044 | 0.099 | 0.663 | 0.753 | 176 | 244 |
| FXST | -0.047 | 0.099 | 0.640 | 0.753 | 177 | 242 |
| GCC | -0.198 | 0.099 | 0.048 | 0.200 | 175 | 244 |
| IC | -0.176 | 0.100 | 0.080 | 0.278 | 175 | 239 |
| IFO | -0.069 | 0.099 | 0.488 | 0.722 | 177 | 242 |
| PCR | -0.012 | 0.099 | 0.905 | 0.942 | 177 | 242 |
| PLIC | -0.109 | 0.099 | 0.275 | 0.528 | 176 | 240 |
| PTR | -0.170 | 0.099 | 0.089 | 0.278 | 177 | 239 |
| RLIC | -0.219 | 0.100 | 0.029 | 0.147 | 177 | 238 |
| SCC | -0.124 | 0.099 | 0.216 | 0.451 | 176 | 239 |
| SCR | -0.128 | 0.099 | 0.201 | 0.451 | 177 | 238 |
| SFO | -0.065 | 0.099 | 0.513 | 0.722 | 177 | 242 |
| SLF | -0.266 | 0.100 | 0.008 | 0.147 | 177 | 240 |
| SS | -0.051 | 0.099 | 0.612 | 0.753 | 176 | 242 |
| UNC | 0.015 | 0.099 | 0.882 | 0.942 | 177 | 242 |
| AverageAD | -0.219 | 0.099 | 0.029 | 0.147 | 177 | 239 |

Table S38. Differences in regional FA between people with a lifetime history an interrupted, aborted or actual attempt and those without a history of an interrupted, aborted or actual suicide attempt determined with the Columbia Suicide Severity Rating Scale (healthy controls were excluded). D: Cohen's d effect size, SE: standard error; p: p-value, FDR-p: FDR corrected p-value, CI: confidence interval.

| Region | D | SE | P | FDR-p | N attempt | N no attempt |
|---|---|---|---|---|---|---|
| ACR | 0.018 | 0.099 | 0.858 | 0.968 | 175 | 243 |
| ALIC | -0.020 | 0.099 | 0.838 | 0.968 | 176 | 241 |
| BCC | -0.009 | 0.099 | 0.930 | 0.968 | 174 | 244 |
| CC | -0.051 | 0.099 | 0.608 | 0.968 | 175 | 242 |
| CGC | -0.123 | 0.099 | 0.218 | 0.968 | 177 | 244 |
| CGH | -0.056 | 0.099 | 0.573 | 0.968 | 177 | 242 |
| CR | 0.030 | 0.099 | 0.768 | 0.968 | 175 | 242 |
| CST | -0.012 | 0.099 | 0.901 | 0.968 | 177 | 242 |
| EC | -0.010 | 0.099 | 0.923 | 0.968 | 176 | 243 |
| FX | 0.000 | 0.099 | 0.999 | 0.999 | 175 | 242 |
| FXST | -0.149 | 0.099 | 0.138 | 0.968 | 175 | 241 |
| GCC | 0.012 | 0.099 | 0.902 | 0.968 | 175 | 243 |
| IC | -0.013 | 0.099 | 0.898 | 0.968 | 176 | 241 |
| IFO | 0.042 | 0.099 | 0.674 | 0.968 | 176 | 243 |
| PCR | -0.020 | 0.099 | 0.843 | 0.968 | 177 | 244 |
| PLIC | 0.077 | 0.099 | 0.441 | 0.968 | 176 | 242 |
| PTR | -0.063 | 0.099 | 0.528 | 0.968 | 175 | 242 |
| RLIC | -0.052 | 0.099 | 0.606 | 0.968 | 176 | 241 |
| SCC | -0.129 | 0.100 | 0.203 | 0.968 | 173 | 237 |
| SCR | 0.059 | 0.099 | 0.551 | 0.968 | 177 | 244 |
| SFO | -0.143 | 0.099 | 0.155 | 0.968 | 175 | 242 |
| SLF | -0.084 | 0.099 | 0.398 | 0.968 | 177 | 244 |
| SS | 0.049 | 0.100 | 0.629 | 0.968 | 175 | 239 |
| UNC | -0.121 | 0.099 | 0.225 | 0.968 | 177 | 243 |
| AverageFA | -0.032 | 0.099 | 0.746 | 0.968 | 175 | 241 |

Table S39. Differences in regional MD between people with a lifetime history an interrupted, aborted or actual attempt and those without a history of an interrupted, aborted or actual suicide attempt determined with the Columbia Suicide Severity Rating Scale (healthy controls were excluded). D: Cohen's d effect size, SE: standard error; p: p-value, FDR-p: FDR corrected p-value, CI: confidence interval.

| Region | D | SE | P | FDR-p | N attempt | N no attempt |
|---|---|---|---|---|---|---|
| ACR | -0.009 | 0.099 | 0.930 | 0.999 | 177 | 242 |
| ALIC | -0.030 | 0.099 | 0.767 | 0.965 | 176 | 244 |
| BCC | -0.085 | 0.099 | 0.396 | 0.761 | 175 | 244 |
| CC | -0.088 | 0.099 | 0.377 | 0.761 | 177 | 244 |
| CGC | -0.017 | 0.099 | 0.868 | 0.999 | 177 | 243 |
| CGH | 0.031 | 0.099 | 0.758 | 0.965 | 177 | 243 |
| CR | -0.005 | 0.099 | 0.962 | 0.999 | 177 | 243 |
| CST | -0.048 | 0.099 | 0.634 | 0.950 | 177 | 237 |
| EC | -0.092 | 0.099 | 0.358 | 0.761 | 176 | 243 |
| FX | -0.095 | 0.099 | 0.346 | 0.761 | 174 | 243 |
| FXST | 0.061 | 0.099 | 0.541 | 0.902 | 176 | 244 |
| GCC | -0.173 | 0.099 | 0.082 | 0.761 | 177 | 244 |
| IC | -0.105 | 0.099 | 0.292 | 0.761 | 175 | 243 |
| IFO | -0.129 | 0.099 | 0.196 | 0.761 | 176 | 244 |
| PCR | 0.012 | 0.099 | 0.906 | 0.999 | 177 | 243 |
| PLIC | -0.093 | 0.099 | 0.352 | 0.761 | 175 | 243 |
| PTR | -0.079 | 0.099 | 0.428 | 0.765 | 175 | 242 |
| RLIC | -0.126 | 0.099 | 0.207 | 0.761 | 176 | 242 |
| SCC | 0.000 | 0.099 | 0.999 | 0.999 | 176 | 242 |
| SCR | -0.046 | 0.099 | 0.646 | 0.950 | 177 | 242 |
| SFO | 0.029 | 0.099 | 0.772 | 0.965 | 177 | 243 |
| SLF | -0.109 | 0.099 | 0.275 | 0.761 | 177 | 242 |
| SS | -0.114 | 0.099 | 0.253 | 0.761 | 176 | 243 |
| UNC | 0.089 | 0.099 | 0.374 | 0.761 | 175 | 244 |
| AverageMD | -0.095 | 0.099 | 0.342 | 0.761 | 176 | 243 |

Table S40. Differences in regional RD between people with a lifetime history an interrupted, aborted or actual attempt and those without a history of an interrupted, aborted or actual suicide attempt determined with the Columbia Suicide Severity Rating Scale (healthy controls were excluded). D: Cohen's d effect size, SE: standard error; p: p-value, FDR-p: FDR corrected p-value, CI: confidence interval.

| Region | D | SE | P | FDR-p | N attempt | N no attempt |
|---|---|---|---|---|---|---|
| ACR | -0.029 | 0.099 | 0.772 | 0.938 | 175 | 241 |
| ALIC | -0.093 | 0.099 | 0.350 | 0.919 | 175 | 244 |
| BCC | -0.033 | 0.099 | 0.740 | 0.938 | 175 | 244 |
| CC | -0.029 | 0.099 | 0.771 | 0.938 | 175 | 244 |
| CGC | -0.078 | 0.099 | 0.437 | 0.919 | 175 | 244 |
| CGH | 0.009 | 0.099 | 0.930 | 0.938 | 177 | 243 |
| CR | -0.008 | 0.099 | 0.938 | 0.938 | 176 | 241 |
| CST | 0.011 | 0.099 | 0.912 | 0.938 | 176 | 240 |
| EC | -0.118 | 0.099 | 0.236 | 0.919 | 176 | 243 |
| FX | -0.121 | 0.100 | 0.228 | 0.919 | 173 | 243 |
| FXST | 0.070 | 0.099 | 0.484 | 0.919 | 177 | 244 |
| GCC | -0.073 | 0.099 | 0.464 | 0.919 | 175 | 243 |
| IC | -0.095 | 0.099 | 0.339 | 0.919 | 176 | 243 |
| IFO | -0.117 | 0.099 | 0.243 | 0.919 | 176 | 244 |
| PCR | 0.013 | 0.099 | 0.897 | 0.938 | 177 | 242 |
| PLIC | -0.103 | 0.099 | 0.303 | 0.919 | 176 | 241 |
| PTR | -0.047 | 0.099 | 0.638 | 0.938 | 174 | 241 |
| RLIC | -0.054 | 0.099 | 0.588 | 0.919 | 176 | 242 |
| SCC | 0.059 | 0.099 | 0.553 | 0.919 | 175 | 242 |
| SCR | -0.071 | 0.099 | 0.477 | 0.919 | 176 | 243 |
| SFO | 0.056 | 0.099 | 0.573 | 0.919 | 177 | 244 |
| SLF | -0.012 | 0.099 | 0.906 | 0.938 | 177 | 243 |
| SS | -0.151 | 0.099 | 0.133 | 0.919 | 175 | 242 |
| UNC | 0.083 | 0.099 | 0.403 | 0.919 | 177 | 244 |
| AverageRD | -0.094 | 0.099 | 0.344 | 0.919 | 176 | 244 |

**Supplemental Note 1.**

While the main analyses of this manuscript focused on comparing healthy controls and clinical controls to participants with recent suicidal ideation or a lifetime suicide attempt, we also performed supplementary analyses in which we compare WM microstructure between healthy controls and clinical controls. These analyses may not reveal new information related to the neural correlates of suicidality, but may help identify alterations related to having a psychiatric diagnosis and contribute to the interpretation of our main findings. These supplementary analyses were performed similar to the main analyses (with age, sex, age-by-sex, age$^2$ and age$^2$-by-sex included as covariates). In order to compare to our main findings, we compared healthy controls and clinical controls in samples included in the analyses on lifetime suicide attempt (*N* CLC= 1871, *N* HC= 642) and in samples included in the analyses on recent suicidal ideation (*N* CLC= 1184, *N* HC= 1240). Results are presented in supplemental tables S41-48. Results showed no significant differences between in global or regional AD, FA, MD or RD between healthy controls and clinical controls in samples included in the analyses on suicide attempt (supplemental tables S41-44). In samples included in the ideation analyses, we observed lower global AD and regional AD in the ACR, CGC, GCC and SLF in the CLC group compared to HC (Cohen's d range: 0.112-0.184; supplemental table S45). In addition, we observed lower global FA and regional FA in the GCC in the CLC group compared to HC (Cohen's D=0.123 and 0.127 respectively; supplemental table S46).

Table S41 Differences in regional AD between clinical controls and healthy controls in samples included in the analysis on suicide attempt
Age, sex, age-by-sex, $age^2$, $age^2$-by-sex included as covariates.

| Region | Cohen's d | SE | CI LB | CI UB | P-value | FDR P-value | HC | CLC |
|---|---|---|---|---|---|---|---|---|
| ACR | -0.056 | 0.051 | -0.155 | 0.043 | 0.269 | 0.770 | 511 | 1680 |
| ALIC | -0.085 | 0.051 | -0.184 | 0.014 | 0.095 | 0.770 | 511 | 1677 |
| BCC | 0.037 | 0.051 | -0.062 | 0.137 | 0.461 | 0.770 | 507 | 1679 |
| CC | 0.009 | 0.051 | -0.091 | 0.108 | 0.863 | 0.926 | 508 | 1676 |
| CGC | -0.084 | 0.050 | -0.183 | 0.015 | 0.096 | 0.770 | 512 | 1683 |
| CGH | 0.036 | 0.050 | -0.063 | 0.135 | 0.478 | 0.770 | 512 | 1682 |
| CR | -0.062 | 0.051 | -0.161 | 0.037 | 0.223 | 0.770 | 511 | 1676 |
| CST | 0.032 | 0.051 | -0.067 | 0.131 | 0.524 | 0.770 | 510 | 1678 |
| EC | -0.057 | 0.051 | -0.156 | 0.042 | 0.260 | 0.770 | 512 | 1669 |
| FX | 0.050 | 0.050 | -0.049 | 0.149 | 0.323 | 0.770 | 512 | 1685 |
| FXST | 0.023 | 0.050 | -0.076 | 0.122 | 0.652 | 0.841 | 512 | 1683 |
| GCC | -0.005 | 0.051 | -0.104 | 0.094 | 0.926 | 0.926 | 509 | 1680 |
| IC | -0.044 | 0.051 | -0.143 | 0.055 | 0.387 | 0.770 | 510 | 1675 |
| IFO | -0.028 | 0.052 | -0.130 | 0.074 | 0.595 | 0.827 | 475 | 1661 |
| PCR | -0.055 | 0.051 | -0.154 | 0.045 | 0.281 | 0.770 | 509 | 1675 |
| PLIC | -0.034 | 0.051 | -0.133 | 0.065 | 0.498 | 0.770 | 511 | 1678 |
| PTR | -0.006 | 0.051 | -0.105 | 0.093 | 0.911 | 0.926 | 511 | 1677 |
| RLIC | -0.005 | 0.050 | -0.104 | 0.094 | 0.924 | 0.926 | 512 | 1676 |
| SCC | 0.021 | 0.051 | -0.078 | 0.120 | 0.674 | 0.841 | 509 | 1678 |
| SCR | -0.077 | 0.051 | -0.177 | 0.022 | 0.127 | 0.770 | 510 | 1671 |
| SFO | -0.039 | 0.051 | -0.138 | 0.060 | 0.441 | 0.770 | 511 | 1673 |
| SLF | -0.067 | 0.050 | -0.166 | 0.032 | 0.184 | 0.770 | 512 | 1680 |
| SS | 0.019 | 0.051 | -0.080 | 0.118 | 0.707 | 0.841 | 511 | 1676 |
| UNC | 0.059 | 0.053 | -0.046 | 0.164 | 0.269 | 0.770 | 447 | 1615 |
| AverageAD | -0.037 | 0.051 | -0.136 | 0.062 | 0.461 | 0.770 | 511 | 1672 |

Table S42 Differences in regional FA between clinical controls and healthy controls in samples included in the analysis on suicide attempt
Age, sex, age-by-sex, age$^2$, age$^2$-by-sex included as covariates.

| Region | Cohen's d | SE | CI LB | CI UB | P-value | FDR P-value | HC | CLC |
|---|---|---|---|---|---|---|---|---|
| ACR | 0.007 | 0.047 | -0.086 | 0.100 | 0.877 | 0.913 | 591 | 1802 |
| ALIC | -0.033 | 0.047 | -0.126 | 0.060 | 0.482 | 0.753 | 591 | 1794 |
| BCC | -0.120 | 0.048 | -0.213 | -0.026 | 0.012 | 0.075 | 588 | 1795 |
| CC | -0.126 | 0.047 | -0.219 | -0.033 | 0.008 | 0.068 | 591 | 1792 |
| CGC | -0.070 | 0.047 | -0.163 | 0.024 | 0.144 | 0.299 | 589 | 1797 |
| CGH | -0.024 | 0.047 | -0.117 | 0.069 | 0.607 | 0.779 | 590 | 1797 |
| CR | -0.044 | 0.047 | -0.137 | 0.049 | 0.357 | 0.595 | 590 | 1795 |
| CST | -0.020 | 0.048 | -0.113 | 0.073 | 0.670 | 0.779 | 588 | 1797 |
| EC | -0.091 | 0.047 | -0.184 | 0.002 | 0.055 | 0.165 | 591 | 1792 |
| FX | -0.031 | 0.047 | -0.124 | 0.062 | 0.512 | 0.753 | 589 | 1792 |
| FXST | -0.070 | 0.047 | -0.163 | 0.023 | 0.138 | 0.299 | 590 | 1795 |
| GCC | -0.131 | 0.047 | -0.224 | -0.038 | 0.006 | 0.068 | 590 | 1801 |
| IC | -0.015 | 0.047 | -0.108 | 0.078 | 0.750 | 0.816 | 589 | 1794 |
| IFO | -0.118 | 0.049 | -0.214 | -0.022 | 0.016 | 0.082 | 546 | 1770 |
| PCR | -0.094 | 0.047 | -0.187 | -0.001 | 0.048 | 0.165 | 591 | 1797 |
| PLIC | -0.005 | 0.047 | -0.098 | 0.088 | 0.924 | 0.924 | 590 | 1793 |
| PTR | -0.060 | 0.047 | -0.153 | 0.033 | 0.204 | 0.364 | 590 | 1797 |
| RLIC | -0.020 | 0.047 | -0.113 | 0.073 | 0.676 | 0.779 | 591 | 1792 |
| SCC | -0.075 | 0.048 | -0.168 | 0.018 | 0.113 | 0.283 | 589 | 1791 |
| SCR | -0.064 | 0.047 | -0.157 | 0.029 | 0.180 | 0.347 | 591 | 1798 |
| SFO | 0.019 | 0.048 | -0.074 | 0.112 | 0.685 | 0.779 | 588 | 1799 |
| SLF | -0.090 | 0.047 | -0.183 | 0.003 | 0.059 | 0.165 | 590 | 1802 |
| SS | -0.024 | 0.047 | -0.117 | 0.069 | 0.617 | 0.779 | 591 | 1794 |
| UNC | -0.144 | 0.050 | -0.241 | -0.046 | 0.004 | 0.068 | 526 | 1732 |
| AverageFA | -0.107 | 0.048 | -0.200 | -0.014 | 0.025 | 0.102 | 589 | 1793 |

Table S43 Differences in regional MD between clinical controls and healthy controls in samples included in the analysis on suicide attempt Age, sex, age-by-sex, $age^2$, $age^2$-by-sex included as covariates.

| Region | Cohen's d | SE | CI LB | CI UB | P-value | FDR P-value | HC | CLC |
|---|---|---|---|---|---|---|---|---|
| ACR | -0.003 | 0.049 | -0.098 | 0.093 | 0.954 | 0.954 | 553 | 1764 |
| ALIC | -0.045 | 0.049 | -0.141 | 0.051 | 0.360 | 0.853 | 550 | 1759 |
| BCC | 0.083 | 0.049 | -0.012 | 0.179 | 0.088 | 0.500 | 549 | 1760 |
| CC | 0.078 | 0.049 | -0.018 | 0.174 | 0.111 | 0.500 | 550 | 1764 |
| CGC | -0.027 | 0.049 | -0.123 | 0.068 | 0.577 | 0.853 | 552 | 1764 |
| CGH | -0.011 | 0.049 | -0.107 | 0.085 | 0.823 | 0.935 | 551 | 1764 |
| CR | 0.012 | 0.049 | -0.083 | 0.108 | 0.800 | 0.935 | 551 | 1762 |
| CST | 0.022 | 0.049 | -0.074 | 0.117 | 0.660 | 0.868 | 552 | 1750 |
| EC | 0.029 | 0.049 | -0.067 | 0.125 | 0.554 | 0.853 | 550 | 1760 |
| FX | 0.073 | 0.049 | -0.023 | 0.169 | 0.137 | 0.500 | 547 | 1755 |
| FXST | 0.051 | 0.049 | -0.045 | 0.146 | 0.301 | 0.853 | 552 | 1759 |
| GCC | 0.081 | 0.049 | -0.014 | 0.177 | 0.095 | 0.500 | 553 | 1767 |
| IC | -0.004 | 0.049 | -0.100 | 0.092 | 0.937 | 0.954 | 550 | 1761 |
| IFO | 0.037 | 0.050 | -0.062 | 0.136 | 0.461 | 0.853 | 508 | 1729 |
| PCR | 0.028 | 0.049 | -0.068 | 0.124 | 0.565 | 0.853 | 550 | 1757 |
| PLIC | -0.015 | 0.049 | -0.111 | 0.080 | 0.753 | 0.935 | 549 | 1762 |
| PTR | 0.035 | 0.049 | -0.060 | 0.131 | 0.472 | 0.853 | 551 | 1762 |
| RLIC | 0.025 | 0.049 | -0.071 | 0.120 | 0.614 | 0.853 | 551 | 1764 |
| SCC | 0.078 | 0.049 | -0.018 | 0.174 | 0.111 | 0.500 | 550 | 1754 |
| SCR | 0.008 | 0.049 | -0.088 | 0.104 | 0.872 | 0.948 | 551 | 1760 |
| SFO | -0.072 | 0.049 | -0.168 | 0.023 | 0.140 | 0.500 | 554 | 1755 |
| SLF | 0.025 | 0.049 | -0.071 | 0.121 | 0.612 | 0.853 | 551 | 1758 |
| SS | 0.040 | 0.049 | -0.055 | 0.136 | 0.408 | 0.853 | 553 | 1756 |
| UNC | 0.092 | 0.051 | -0.009 | 0.193 | 0.075 | 0.500 | 487 | 1692 |
| AverageMD | 0.044 | 0.049 | -0.052 | 0.140 | 0.368 | 0.853 | 549 | 1763 |

Table S44 Differences in regional RD between clinical controls and healthy controls in samples included in the analysis on suicide attempt
Age, sex, age-by-sex, $age^2$, $age^2$-by-sex included as covariates.

| Region | Cohen's d | SE | CI LB | CI UB | P-value | FDR P-value | HC | CLC |
|---|---|---|---|---|---|---|---|---|
| ACR | 0.016 | 0.049 | -0.080 | 0.111 | 0.750 | 0.812 | 553 | 1761 |
| ALIC | -0.005 | 0.049 | -0.100 | 0.091 | 0.922 | 0.922 | 551 | 1758 |
| BCC | 0.089 | 0.049 | -0.007 | 0.185 | 0.070 | 0.630 | 549 | 1763 |
| CC | 0.074 | 0.049 | -0.022 | 0.170 | 0.130 | 0.630 | 552 | 1764 |
| CGC | 0.014 | 0.049 | -0.082 | 0.109 | 0.779 | 0.812 | 552 | 1762 |
| CGH | 0.019 | 0.049 | -0.077 | 0.114 | 0.702 | 0.812 | 553 | 1763 |
| CR | 0.040 | 0.049 | -0.055 | 0.136 | 0.409 | 0.723 | 552 | 1762 |
| CST | 0.017 | 0.049 | -0.078 | 0.113 | 0.723 | 0.812 | 551 | 1762 |
| EC | 0.052 | 0.049 | -0.043 | 0.148 | 0.286 | 0.630 | 553 | 1757 |
| FX | 0.091 | 0.049 | -0.005 | 0.187 | 0.064 | 0.630 | 545 | 1755 |
| FXST | 0.049 | 0.049 | -0.046 | 0.145 | 0.312 | 0.630 | 553 | 1760 |
| GCC | 0.074 | 0.049 | -0.021 | 0.170 | 0.129 | 0.630 | 553 | 1762 |
| IC | 0.036 | 0.049 | -0.060 | 0.132 | 0.462 | 0.723 | 551 | 1761 |
| IFO | 0.071 | 0.050 | -0.028 | 0.170 | 0.162 | 0.630 | 508 | 1728 |
| PCR | 0.048 | 0.049 | -0.048 | 0.144 | 0.328 | 0.630 | 551 | 1759 |
| PLIC | 0.019 | 0.049 | -0.077 | 0.115 | 0.699 | 0.812 | 552 | 1757 |
| PTR | 0.020 | 0.049 | -0.076 | 0.116 | 0.683 | 0.812 | 552 | 1761 |
| RLIC | 0.027 | 0.049 | -0.069 | 0.122 | 0.581 | 0.812 | 553 | 1761 |
| SCC | 0.058 | 0.049 | -0.038 | 0.153 | 0.239 | 0.630 | 550 | 1752 |
| SCR | 0.038 | 0.049 | -0.058 | 0.133 | 0.439 | 0.723 | 552 | 1763 |
| SFO | -0.058 | 0.049 | -0.153 | 0.038 | 0.236 | 0.630 | 553 | 1761 |
| SLF | 0.048 | 0.049 | -0.048 | 0.144 | 0.326 | 0.630 | 552 | 1762 |
| SS | 0.019 | 0.049 | -0.077 | 0.114 | 0.699 | 0.812 | 554 | 1757 |
| UNC | 0.080 | 0.051 | -0.021 | 0.181 | 0.121 | 0.630 | 487 | 1692 |
| AverageRD | 0.066 | 0.049 | -0.030 | 0.161 | 0.179 | 0.630 | 552 | 1762 |

Table S45 Differences in regional AD between clinical controls and healthy controls in samples included in the analysis on suicidal ideation
Age, sex, age-by-sex, age$^2$, age$^2$-by-sex included as covariates.

| Region | Cohen's d | SE | CI LB | CI UB | P-value | FDR P-value | HC | CLC |
|---|---|---|---|---|---|---|---|---|
| ACR | -0.132 | 0.041 | -0.213 | -0.051 | 0.001 | 0.012 | 1217 | 1136 |
| ALIC | -0.050 | 0.041 | -0.131 | 0.031 | 0.229 | 0.446 | 1206 | 1139 |
| BCC | -0.032 | 0.041 | -0.113 | 0.049 | 0.442 | 0.614 | 1211 | 1138 |
| CC | -0.080 | 0.041 | -0.161 | 0.001 | 0.054 | 0.209 | 1214 | 1137 |
| CGC | -0.184 | 0.041 | -0.265 | -0.103 | 0.000 | 0.000 | 1214 | 1137 |
| CGH | 0.049 | 0.041 | -0.031 | 0.130 | 0.232 | 0.446 | 1216 | 1138 |
| CR | -0.078 | 0.041 | -0.159 | 0.003 | 0.059 | 0.209 | 1216 | 1136 |
| CST | 0.018 | 0.041 | -0.063 | 0.099 | 0.666 | 0.724 | 1206 | 1136 |
| EC | -0.072 | 0.041 | -0.153 | 0.009 | 0.081 | 0.228 | 1217 | 1134 |
| FX | 0.051 | 0.041 | -0.030 | 0.132 | 0.219 | 0.446 | 1214 | 1130 |
| FXST | 0.008 | 0.041 | -0.073 | 0.089 | 0.842 | 0.877 | 1219 | 1140 |
| GCC | -0.113 | 0.041 | -0.194 | -0.032 | 0.006 | 0.033 | 1215 | 1136 |
| IC | -0.032 | 0.041 | -0.113 | 0.049 | 0.442 | 0.614 | 1214 | 1139 |
| IFO | 0.022 | 0.041 | -0.059 | 0.103 | 0.597 | 0.679 | 1213 | 1139 |
| PCR | -0.039 | 0.041 | -0.120 | 0.042 | 0.342 | 0.562 | 1212 | 1134 |
| PLIC | -0.023 | 0.041 | -0.104 | 0.058 | 0.579 | 0.679 | 1216 | 1139 |
| PTR | -0.023 | 0.041 | -0.104 | 0.057 | 0.572 | 0.679 | 1217 | 1137 |
| RLIC | -0.038 | 0.041 | -0.119 | 0.043 | 0.360 | 0.562 | 1220 | 1137 |
| SCC | -0.072 | 0.041 | -0.153 | 0.009 | 0.082 | 0.228 | 1214 | 1137 |
| SCR | -0.029 | 0.041 | -0.110 | 0.052 | 0.478 | 0.629 | 1215 | 1136 |
| SFO | 0.006 | 0.041 | -0.075 | 0.087 | 0.888 | 0.888 | 1211 | 1132 |
| SLF | -0.112 | 0.041 | -0.193 | -0.031 | 0.007 | 0.033 | 1217 | 1134 |
| SS | 0.041 | 0.041 | -0.040 | 0.122 | 0.322 | 0.562 | 1216 | 1134 |
| UNC | 0.063 | 0.042 | -0.020 | 0.146 | 0.139 | 0.347 | 1152 | 1088 |
| AverageAD | -0.157 | 0.041 | -0.239 | -0.076 | 0.000 | 0.002 | 1214 | 1134 |

Table S46 Differences in regional FA between clinical controls and healthy controls in samples included in the analysis on suicidal ideation Age, sex, age-by-sex, age$^2$, age$^2$-by-sex included as covariates.

| Region | Cohen's d | SE | CI LB | CI UB | P-value | FDR P-value | HC | CLC |
|---|---|---|---|---|---|---|---|---|
| ACR | -0.061 | 0.041 | -0.141 | 0.019 | 0.135 | 0.303 | 1236 | 1182 |
| ALIC | -0.071 | 0.041 | -0.151 | 0.009 | 0.083 | 0.226 | 1235 | 1177 |
| BCC | -0.093 | 0.041 | -0.173 | -0.013 | 0.024 | 0.147 | 1232 | 1174 |
| CC | -0.111 | 0.041 | -0.191 | -0.032 | 0.006 | 0.053 | 1233 | 1175 |
| CGC | -0.059 | 0.041 | -0.139 | 0.020 | 0.145 | 0.303 | 1233 | 1177 |
| CGH | -0.018 | 0.041 | -0.097 | 0.062 | 0.666 | 0.742 | 1234 | 1176 |
| CR | -0.069 | 0.041 | -0.149 | 0.010 | 0.089 | 0.226 | 1233 | 1179 |
| CST | -0.022 | 0.041 | -0.102 | 0.059 | 0.599 | 0.742 | 1223 | 1176 |
| EC | -0.069 | 0.041 | -0.149 | 0.011 | 0.090 | 0.226 | 1230 | 1175 |
| FX | -0.057 | 0.041 | -0.137 | 0.023 | 0.162 | 0.311 | 1230 | 1177 |
| FXST | -0.014 | 0.041 | -0.094 | 0.066 | 0.736 | 0.742 | 1234 | 1176 |
| GCC | -0.127 | 0.041 | -0.207 | -0.047 | 0.002 | 0.036 | 1233 | 1177 |
| IC | -0.019 | 0.041 | -0.099 | 0.061 | 0.640 | 0.742 | 1235 | 1178 |
| IFO | -0.039 | 0.041 | -0.119 | 0.041 | 0.342 | 0.534 | 1226 | 1182 |
| PCR | -0.071 | 0.041 | -0.151 | 0.009 | 0.083 | 0.226 | 1234 | 1179 |
| PLIC | -0.016 | 0.041 | -0.096 | 0.063 | 0.688 | 0.742 | 1234 | 1176 |
| PTR | -0.026 | 0.041 | -0.106 | 0.053 | 0.518 | 0.720 | 1235 | 1179 |
| RLIC | 0.021 | 0.041 | -0.059 | 0.101 | 0.608 | 0.742 | 1232 | 1178 |
| SCC | -0.083 | 0.041 | -0.163 | -0.003 | 0.043 | 0.217 | 1230 | 1176 |
| SCR | -0.052 | 0.041 | -0.132 | 0.028 | 0.203 | 0.363 | 1234 | 1180 |
| SFO | -0.040 | 0.041 | -0.120 | 0.040 | 0.325 | 0.534 | 1232 | 1180 |
| SLF | -0.075 | 0.041 | -0.154 | 0.005 | 0.068 | 0.226 | 1233 | 1183 |
| SS | -0.013 | 0.041 | -0.093 | 0.066 | 0.742 | 0.742 | 1230 | 1180 |
| UNC | -0.035 | 0.042 | -0.117 | 0.047 | 0.402 | 0.592 | 1169 | 1131 |
| AverageFA | -0.123 | 0.041 | -0.203 | -0.042 | 0.003 | 0.036 | 1231 | 1140 |

Table S47 Differences in regional MD between clinical controls and healthy controls in samples included in the analysis on suicidal ideation
Age, sex, age-by-sex, age$^2$, age$^2$-by-sex included as covariates.

| Region | Cohen's d | SE | CI LB | CI UB | P-value | FDR P-value | HC | CLC |
|---|---|---|---|---|---|---|---|---|
| ACR | -0.032 | 0.041 | -0.112 | 0.048 | 0.435 | 0.777 | 1217 | 1175 |
| ALIC | 0.003 | 0.041 | -0.077 | 0.084 | 0.940 | 0.970 | 1204 | 1173 |
| BCC | 0.070 | 0.041 | -0.010 | 0.150 | 0.089 | 0.507 | 1213 | 1174 |
| CC | 0.051 | 0.041 | -0.029 | 0.132 | 0.210 | 0.507 | 1214 | 1178 |
| CGC | -0.094 | 0.041 | -0.174 | -0.014 | 0.022 | 0.276 | 1214 | 1177 |
| CGH | 0.061 | 0.041 | -0.019 | 0.141 | 0.137 | 0.507 | 1219 | 1176 |
| CR | 0.012 | 0.041 | -0.068 | 0.092 | 0.771 | 0.948 | 1216 | 1176 |
| CST | 0.011 | 0.041 | -0.070 | 0.091 | 0.797 | 0.948 | 1203 | 1170 |
| EC | 0.024 | 0.041 | -0.056 | 0.104 | 0.562 | 0.826 | 1209 | 1174 |
| FX | 0.021 | 0.041 | -0.060 | 0.101 | 0.617 | 0.857 | 1209 | 1162 |
| FXST | 0.052 | 0.041 | -0.029 | 0.132 | 0.208 | 0.507 | 1215 | 1174 |
| GCC | -0.007 | 0.041 | -0.087 | 0.073 | 0.863 | 0.948 | 1216 | 1177 |
| IC | -0.007 | 0.041 | -0.087 | 0.074 | 0.871 | 0.948 | 1212 | 1173 |
| IFO | 0.074 | 0.041 | -0.006 | 0.154 | 0.070 | 0.507 | 1213 | 1175 |
| PCR | 0.052 | 0.041 | -0.028 | 0.132 | 0.202 | 0.507 | 1217 | 1175 |
| PLIC | -0.052 | 0.041 | -0.133 | 0.028 | 0.201 | 0.507 | 1215 | 1175 |
| PTR | 0.052 | 0.041 | -0.028 | 0.133 | 0.200 | 0.507 | 1215 | 1176 |
| RLIC | -0.007 | 0.041 | -0.087 | 0.074 | 0.872 | 0.948 | 1214 | 1177 |
| SCC | 0.035 | 0.041 | -0.046 | 0.115 | 0.399 | 0.766 | 1212 | 1174 |
| SCR | 0.050 | 0.041 | -0.030 | 0.130 | 0.223 | 0.507 | 1214 | 1173 |
| SFO | 0.025 | 0.041 | -0.055 | 0.106 | 0.539 | 0.826 | 1212 | 1171 |
| SLF | -0.002 | 0.041 | -0.082 | 0.079 | 0.970 | 0.970 | 1216 | 1174 |
| SS | 0.045 | 0.041 | -0.035 | 0.125 | 0.270 | 0.564 | 1219 | 1171 |
| UNC | 0.115 | 0.042 | 0.033 | 0.197 | 0.006 | 0.155 | 1145 | 1127 |
| AverageMD | 0.028 | 0.041 | -0.053 | 0.109 | 0.505 | 0.826 | 1212 | 1136 |

Table S48 Differences in regional RD between clinical controls and healthy controls in samples included in the analysis on suicidal ideation
Age, sex, age-by-sex, age$^2$, age$^2$-by-sex included as covariates.

| Region | Cohen's d | SE | CI LB | CI UB | P-value | FDR P-value | HC | CLC |
|---|---|---|---|---|---|---|---|---|
| ACR | 0.006 | 0.041 | -0.075 | 0.087 | 0.883 | 0.920 | 1214 | 1135 |
| ALIC | 0.022 | 0.041 | -0.059 | 0.103 | 0.593 | 0.743 | 1206 | 1133 |
| BCC | 0.091 | 0.041 | 0.010 | 0.172 | 0.028 | 0.421 | 1211 | 1137 |
| CC | 0.072 | 0.041 | -0.009 | 0.153 | 0.081 | 0.421 | 1214 | 1135 |
| CGC | -0.033 | 0.041 | -0.114 | 0.048 | 0.425 | 0.626 | 1213 | 1136 |
| CGH | 0.048 | 0.041 | -0.033 | 0.129 | 0.244 | 0.611 | 1221 | 1135 |
| CR | 0.039 | 0.041 | -0.042 | 0.120 | 0.350 | 0.622 | 1215 | 1135 |
| CST | 0.038 | 0.041 | -0.043 | 0.119 | 0.357 | 0.622 | 1202 | 1137 |
| EC | 0.037 | 0.041 | -0.044 | 0.118 | 0.374 | 0.622 | 1213 | 1132 |
| FX | 0.042 | 0.041 | -0.040 | 0.123 | 0.315 | 0.622 | 1206 | 1122 |
| FXST | 0.015 | 0.041 | -0.066 | 0.096 | 0.712 | 0.847 | 1215 | 1136 |
| GCC | 0.040 | 0.041 | -0.041 | 0.121 | 0.329 | 0.622 | 1215 | 1134 |
| IC | -0.008 | 0.041 | -0.089 | 0.073 | 0.846 | 0.920 | 1213 | 1136 |
| IFO | 0.068 | 0.041 | -0.013 | 0.149 | 0.101 | 0.421 | 1211 | 1133 |
| PCR | 0.048 | 0.041 | -0.033 | 0.129 | 0.243 | 0.611 | 1213 | 1136 |
| PLIC | -0.035 | 0.041 | -0.116 | 0.046 | 0.398 | 0.622 | 1215 | 1135 |
| PTR | 0.053 | 0.041 | -0.028 | 0.134 | 0.200 | 0.611 | 1214 | 1136 |
| RLIC | -0.022 | 0.041 | -0.103 | 0.059 | 0.594 | 0.743 | 1215 | 1137 |
| SCC | 0.073 | 0.041 | -0.008 | 0.154 | 0.078 | 0.421 | 1207 | 1133 |
| SCR | 0.058 | 0.041 | -0.023 | 0.139 | 0.158 | 0.563 | 1215 | 1134 |
| SFO | 0.007 | 0.041 | -0.074 | 0.088 | 0.870 | 0.920 | 1209 | 1135 |
| SLF | 0.025 | 0.041 | -0.056 | 0.106 | 0.547 | 0.743 | 1214 | 1136 |
| SS | -0.002 | 0.041 | -0.083 | 0.078 | 0.953 | 0.953 | 1219 | 1132 |
| UNC | 0.075 | 0.042 | -0.009 | 0.158 | 0.079 | 0.421 | 1143 | 1087 |
| AverageRD | 0.071 | 0.041 | -0.010 | 0.152 | 0.084 | 0.421 | 1214 | 1136 |

**Supplemental note 2.**

The ENIGMA-Suicidal Thoughts and Behaviours consortium is transdiagnostic, and includes participants with various types of psychiatric illness, including major depressive disorder, bipolar disorder, anxiety disorders, obsessive-compulsive disorder, posttraumatic stress disorder and psychotic disorders. We were interested to examine if the association between suicidality and WM microstructure differed according to diagnosis type. In supplemental analyses, we aimed to examine interaction effects between group (clinical controls and recent suicidal ideation or clinical controls and lifetime suicide attempt) and type of lifetime psychiatric disorder. Healthy controls were not included in this analysis as they did not have a psychiatric disorder. As information on lifetime type of psychiatric disorder was only available for a subsample of participants (some sites only assessed current disorders), these analyses were performed as supplemental analyses.

Lifetime diagnosis type was categorized into six main lifetime diagnosis types: MDD, BD, anxiety disorders, PTSD, OCD and psychotic disorders. In the suicide attempt-by-diagnosis interaction analyses, we were only able to include MDD (N=1139, of whom 355 attempted suicide), BD (N=553; of whom 141 attempted suicide) and psychotic disorders (N=198; of whom 44 attempted suicide) given the small sample sizes per lifetime diagnosis type. Similarly, in the recent suicidal ideation-by-diagnosis interaction analyses, we were only able to include MDD (N=1128 of whom 424 had recent suicidal ideation) and BD (N=225; of whom 114 had recent suicidal ideation).

Similar to our main analyses, we used multiple linear regression models in R, in which we included the group-by-diagnosis type interaction term, and main effects of group and diagnosis type. Results of these analyses are presented in supplemental tables S49-56. Results show that there are no significant interaction effects between suicide attempt group (clinical control or lifetime suicide attempt) and diagnosis type (MDD, BD or psychotic disorder) on WM microstructure (supplemental tables S49-52). In contrast, we observed many significant interaction effects between suicidal ideation group (clinical control or recent suicidal ideation) and diagnosis type (MDD or BD) on WM microstructure, specifically on regional FA, MD or RD (supplemental tables 53-56). These findings indicate that the association between ideation and these WM microstructure metrics differs in MDD patients and BD patients. To follow-up on this, we performed stratified analyses, looking at the effect of recent suicidal ideation (clinical controls vs. recent ideation) in MDD patients and BD patients separately (with age, sex, age-by-sex, $age^2$ and $age^2$-by-sex included as covariates) for regions where we found significant interaction effects. Results of these stratified analyses are presented in supplemental table S57. For most of these associations, the strength of the association appeared to differ between MDD and BD patients, but the associations were not significant in the MDD only sample or BD only sample. Plotting the results showed that the difference between clinical controls and ideators appeared larger in the BD group, than in the MDD group.

Table S49. Interaction of attempt group-by-type of psychiatric diagnosis on regional AD measures

| Region | P-value | FDR P-value | Attempt | CLC |
|---|---|---|---|---|
| ACR | 0.151 | 0.497 | 503 | 1240 |
| ALIC | 0.407 | 0.848 | 502 | 1238 |
| BCC | 0.490 | 0.942 | 501 | 1238 |
| CC | 0.293 | 0.740 | 502 | 1238 |
| CGC | 0.759 | 0.959 | 505 | 1244 |
| CGH | 0.122 | 0.497 | 502 | 1244 |
| CR | 0.853 | 0.959 | 503 | 1236 |
| CST | 0.148 | 0.497 | 503 | 1239 |
| EC | 0.730 | 0.959 | 503 | 1233 |
| FX | 0.159 | 0.497 | 504 | 1245 |
| FXST | 0.022 | 0.281 | 505 | 1242 |
| GCC | 0.021 | 0.281 | 503 | 1242 |
| IC | 0.882 | 0.959 | 502 | 1236 |
| IFO | 0.075 | 0.497 | 503 | 1243 |
| PCR | 0.328 | 0.745 | 503 | 1236 |
| PLIC | 0.840 | 0.959 | 503 | 1239 |
| PTR | 0.155 | 0.497 | 504 | 1236 |
| RLIC | 0.296 | 0.740 | 504 | 1236 |
| SCC | 0.775 | 0.959 | 502 | 1241 |
| SCR | 0.973 | 0.973 | 503 | 1231 |
| SFO | 0.622 | 0.959 | 503 | 1238 |
| SLF | 0.878 | 0.959 | 505 | 1240 |
| SS | 0.577 | 0.959 | 504 | 1239 |
| UNC | 0.850 | 0.959 | 495 | 1220 |
| AverageAD | 0.949 | 0.973 | 505 | 1232 |

Table S50. Interaction of attempt group-by-type of psychiatric diagnosis on regional FA measures

| Region | P-value | FDR P-value | Attempt | CLC |
|---|---|---|---|---|
| ACR | 0.377 | 0.884 | 525 | 1286 |
| ALIC | 0.442 | 0.884 | 523 | 1280 |
| BCC | 0.441 | 0.884 | 521 | 1283 |
| CC | 0.201 | 0.839 | 521 | 1278 |
| CGC | 0.652 | 0.884 | 526 | 1286 |
| CGH | 0.773 | 0.884 | 524 | 1283 |
| CR | 0.621 | 0.884 | 525 | 1280 |
| CST | 0.720 | 0.884 | 523 | 1283 |
| EC | 0.506 | 0.884 | 525 | 1279 |
| FX | 0.285 | 0.884 | 520 | 1279 |
| FXST | 0.720 | 0.884 | 524 | 1282 |
| GCC | 0.085 | 0.597 | 524 | 1285 |
| IC | 0.486 | 0.884 | 525 | 1279 |
| IFO | 0.038 | 0.475 | 526 | 1283 |
| PCR | 0.778 | 0.884 | 526 | 1282 |
| PLIC | 0.441 | 0.884 | 524 | 1280 |
| PTR | 0.767 | 0.884 | 521 | 1284 |
| RLIC | 0.967 | 0.981 | 523 | 1280 |
| SCC | 0.027 | 0.475 | 522 | 1278 |
| SCR | 0.981 | 0.981 | 524 | 1283 |
| SFO | 0.920 | 0.981 | 521 | 1283 |
| SLF | 0.294 | 0.884 | 522 | 1286 |
| SS | 0.172 | 0.839 | 523 | 1278 |
| UNC | 0.096 | 0.597 | 516 | 1263 |
| AverageFA | 0.748 | 0.884 | 525 | 1278 |

Table S51. Interaction of attempt group-by-type of psychiatric diagnosis on regional MD measures

| Region | P-value | FDR P-value | Attempt | CLC |
|---|---|---|---|---|
| ACR | 0.024 | 0.218 | 511 | 1264 |
| ALIC | 0.799 | 0.868 | 508 | 1262 |
| BCC | 0.942 | 0.942 | 509 | 1263 |
| CC | 0.385 | 0.561 | 511 | 1267 |
| CGC | 0.215 | 0.518 | 510 | 1267 |
| CGH | 0.375 | 0.561 | 509 | 1268 |
| CR | 0.107 | 0.447 | 512 | 1264 |
| CST | 0.315 | 0.561 | 508 | 1258 |
| EC | 0.142 | 0.506 | 510 | 1263 |
| FX | 0.067 | 0.336 | 509 | 1256 |
| FXST | 0.028 | 0.218 | 511 | 1265 |
| GCC | 0.005 | 0.117 | 513 | 1269 |
| IC | 0.375 | 0.561 | 508 | 1262 |
| IFO | 0.849 | 0.884 | 509 | 1261 |
| PCR | 0.182 | 0.506 | 511 | 1260 |
| PLIC | 0.592 | 0.672 | 506 | 1263 |
| PTR | 0.174 | 0.506 | 511 | 1264 |
| RLIC | 0.365 | 0.561 | 512 | 1266 |
| SCC | 0.559 | 0.672 | 511 | 1261 |
| SCR | 0.588 | 0.672 | 513 | 1262 |
| SFO | 0.427 | 0.561 | 509 | 1260 |
| SLF | 0.410 | 0.561 | 510 | 1260 |
| SS | 0.035 | 0.218 | 510 | 1262 |
| UNC | 0.271 | 0.561 | 499 | 1241 |
| AverageMD | 0.228 | 0.518 | 512 | 1264 |

Table S52. Interaction of attempt group-by-type of psychiatric diagnosis on regional RD measures

| Region | P-value | FDR P-value | Attempt | CLC |
|---|---|---|---|---|
| ACR | 0.028 | 0.231 | 511 | 1262 |
| ALIC | 0.608 | 0.799 | 507 | 1263 |
| BCC | 0.341 | 0.609 | 509 | 1265 |
| CC | 0.119 | 0.430 | 510 | 1267 |
| CGC | 0.210 | 0.478 | 510 | 1267 |
| CGH | 0.985 | 0.985 | 510 | 1268 |
| CR | 0.169 | 0.430 | 511 | 1263 |
| CST | 0.474 | 0.698 | 507 | 1266 |
| EC | 0.156 | 0.430 | 511 | 1263 |
| FX | 0.064 | 0.401 | 508 | 1256 |
| FXST | 0.794 | 0.863 | 513 | 1267 |
| GCC | 0.002 | 0.059 | 510 | 1264 |
| IC | 0.922 | 0.960 | 510 | 1264 |
| IFO | 0.373 | 0.622 | 513 | 1259 |
| PCR | 0.294 | 0.565 | 513 | 1263 |
| PLIC | 0.752 | 0.855 | 508 | 1262 |
| PTR | 0.403 | 0.630 | 510 | 1263 |
| RLIC | 0.721 | 0.855 | 511 | 1265 |
| SCC | 0.128 | 0.430 | 510 | 1260 |
| SCR | 0.673 | 0.841 | 511 | 1265 |
| SFO | 0.536 | 0.745 | 509 | 1264 |
| SLF | 0.274 | 0.565 | 511 | 1264 |
| SS | 0.026 | 0.231 | 510 | 1264 |
| UNC | 0.090 | 0.430 | 502 | 1242 |
| AverageRD | 0.172 | 0.430 | 511 | 1264 |

Table S53. Interaction of ideation group-by-type of psychiatric diagnosis on regional AD measures

| Region | P | FDR-P | Ideation | CLC |
|---|---|---|---|---|
| ACR | 0.336 | 0.621 | 502 | 769 |
| ALIC | 0.398 | 0.621 | 499 | 773 |
| BCC | 0.027 | 0.167 | 499 | 769 |
| CC | 0.056 | 0.215 | 502 | 770 |
| CGC | 0.060 | 0.215 | 502 | 772 |
| CGH | 0.365 | 0.621 | 498 | 771 |
| CR | 0.777 | 0.809 | 501 | 769 |
| CST | 0.303 | 0.621 | 498 | 768 |
| EC | 0.033 | 0.167 | 496 | 769 |
| FX | 0.000 | 0.000 | 501 | 766 |
| FXST | 0.019 | 0.161 | 501 | 772 |
| GCC | 0.381 | 0.621 | 501 | 769 |
| IC | 0.723 | 0.786 | 499 | 772 |
| IFO | 0.848 | 0.848 | 502 | 772 |
| PCR | 0.693 | 0.786 | 500 | 768 |
| PLIC | 0.077 | 0.242 | 498 | 772 |
| PTR | 0.434 | 0.638 | 499 | 770 |
| RLIC | 0.712 | 0.786 | 498 | 771 |
| SCC | 0.354 | 0.621 | 503 | 771 |
| SCR | 0.542 | 0.713 | 500 | 769 |
| SFO | 0.158 | 0.438 | 499 | 768 |
| SLF | 0.506 | 0.703 | 503 | 768 |
| SS | 0.681 | 0.786 | 501 | 770 |
| UNC | 0.011 | 0.140 | 497 | 756 |
| AverageAD | 0.392 | 0.621 | 499 | 767 |

Table S54. Interaction of ideation group-by-type of psychiatric diagnosis on regional FA measures

| Region | P | FDR-P | Ideation | CLC |
|---|---|---|---|---|
| ACR | 0.000 | 0.000 | 537 | 813 |
| ALIC | 0.375 | 0.395 | 537 | 810 |
| BCC | 0.001 | 0.004 | 534 | 810 |
| CC | 0.000 | 0.001 | 535 | 808 |
| CGC | 0.290 | 0.362 | 537 | 813 |
| CGH | 0.379 | 0.395 | 536 | 810 |
| CR | 0.000 | 0.001 | 536 | 810 |
| CST | 0.378 | 0.395 | 535 | 809 |
| EC | 0.031 | 0.052 | 537 | 809 |
| FX | 0.001 | 0.004 | 535 | 812 |
| FXST | 0.001 | 0.003 | 535 | 811 |
| GCC | 0.000 | 0.000 | 536 | 809 |
| IC | 0.061 | 0.090 | 534 | 810 |
| IFO | 0.905 | 0.905 | 534 | 814 |
| PCR | 0.007 | 0.015 | 536 | 811 |
| PLIC | 0.152 | 0.200 | 531 | 809 |
| PTR | 0.000 | 0.001 | 535 | 811 |
| RLIC | 0.044 | 0.069 | 536 | 811 |
| SCC | 0.019 | 0.037 | 536 | 809 |
| SCR | 0.013 | 0.027 | 535 | 812 |
| SFO | 0.368 | 0.395 | 534 | 814 |
| SLF | 0.021 | 0.037 | 537 | 814 |
| SS | 0.001 | 0.002 | 534 | 811 |
| UNC | 0.073 | 0.102 | 532 | 798 |
| AverageFA | 0.004 | 0.011 | 509 | 771 |

Table S55. Interaction of ideation group-by-type of psychiatric diagnosis on regional MD measures

| Region | P | FDR-P | Ideation | CLC |
|---|---|---|---|---|
| ACR | 0.003 | 0.009 | 528 | 808 |
| ALIC | 0.342 | 0.389 | 524 | 807 |
| BCC | 0.001 | 0.003 | 527 | 807 |
| CC | 0.002 | 0.009 | 527 | 810 |
| CGC | 0.008 | 0.019 | 526 | 810 |
| CGH | 0.888 | 0.888 | 527 | 810 |
| CR | 0.003 | 0.009 | 527 | 809 |
| CST | 0.051 | 0.086 | 523 | 805 |
| EC | 0.000 | 0.001 | 523 | 809 |
| FX | 0.000 | 0.000 | 528 | 799 |
| FXST | 0.364 | 0.395 | 528 | 809 |
| GCC | 0.000 | 0.003 | 529 | 809 |
| IC | 0.264 | 0.330 | 525 | 806 |
| IFO | 0.295 | 0.352 | 526 | 807 |
| PCR | 0.017 | 0.032 | 526 | 807 |
| PLIC | 0.583 | 0.608 | 523 | 808 |
| PTR | 0.013 | 0.027 | 525 | 808 |
| RLIC | 0.072 | 0.106 | 528 | 809 |
| SCC | 0.141 | 0.186 | 526 | 807 |
| SCR | 0.004 | 0.012 | 528 | 807 |
| SFO | 0.059 | 0.092 | 524 | 805 |
| SLF | 0.008 | 0.019 | 526 | 807 |
| SS | 0.027 | 0.047 | 525 | 807 |
| UNC | 0.000 | 0.003 | 521 | 794 |
| AverageMD | 0.098 | 0.136 | 500 | 769 |

Table S56. Interaction of ideation group-by-type of psychiatric diagnosis on regional RD measures

| Region | P | FDR-P | Ideation | CLC |
|---|---|---|---|---|
| ACR | 0.000 | 0.000 | 501 | 767 |
| ALIC | 0.258 | 0.281 | 499 | 768 |
| BCC | 0.002 | 0.006 | 499 | 769 |
| CC | 0.000 | 0.002 | 500 | 768 |
| CGC | 0.068 | 0.090 | 499 | 770 |
| CGH | 0.618 | 0.644 | 500 | 769 |
| CR | 0.000 | 0.000 | 500 | 767 |
| CST | 0.121 | 0.144 | 498 | 770 |
| EC | 0.000 | 0.001 | 500 | 767 |
| FX | 0.000 | 0.000 | 502 | 759 |
| FXST | 0.014 | 0.026 | 501 | 770 |
| GCC | 0.001 | 0.002 | 503 | 766 |
| IC | 0.021 | 0.035 | 498 | 769 |
| IFO | 0.657 | 0.657 | 500 | 765 |
| PCR | 0.007 | 0.014 | 500 | 769 |
| PLIC | 0.186 | 0.212 | 497 | 768 |
| PTR | 0.004 | 0.009 | 500 | 768 |
| RLIC | 0.057 | 0.079 | 501 | 768 |
| SCC | 0.040 | 0.059 | 501 | 766 |
| SCR | 0.001 | 0.002 | 501 | 767 |
| SFO | 0.116 | 0.144 | 498 | 770 |
| SLF | 0.001 | 0.002 | 502 | 768 |
| SS | 0.003 | 0.008 | 500 | 768 |
| UNC | 0.024 | 0.037 | 496 | 755 |
| AverageRD | 0.003 | 0.008 | 502 | 768 |

Table S57. Follow-up stratified analyses: association between group (clinical controls or recent suicidal ideation) and brain metrics that showed significant interaction effects in MDD patients only or in BD patients only.

| | | MDD only | | | BD only | | |
|---|---|---|---|---|---|---|---|
| Measure | Region | Cohen's D | SE | P-value | Cohen's D | SE | P-value |
| AD | FX | -0.119 | 0.064 | 0.065 | -0.127 | 0.134 | 0.351 |
| FA | ACR | -0.043 | 0.062 | 0.483 | 0.042 | 0.133 | 0.757 |
| FA | BCC | 0.042 | 0.062 | 0.499 | 0.039 | 0.134 | 0.774 |
| FA | CC | 0.009 | 0.062 | 0.885 | 0.000 | 0.134 | 1.000 |
| FA | CR | -0.089 | 0.062 | 0.149 | 0.041 | 0.133 | 0.762 |
| FA | FX | 0.060 | 0.062 | 0.328 | 0.253 | 0.134 | 0.063 |
| FA | FXST | -0.079 | 0.062 | 0.201 | -0.098 | 0.134 | 0.469 |
| FA | GCC | -0.008 | 0.062 | 0.903 | 0.007 | 0.134 | 0.956 |
| FA | PCR | -0.090 | 0.062 | 0.146 | -0.068 | 0.134 | 0.616 |
| FA | PTR | -0.125 | 0.062 | 0.043 | -0.107 | 0.134 | 0.433 |
| FA | SCC | 0.029 | 0.062 | 0.636 | -0.103 | 0.134 | 0.451 |
| FA | SCR | -0.064 | 0.062 | 0.299 | 0.080 | 0.133 | 0.556 |
| FA | SLF | -0.051 | 0.062 | 0.411 | -0.001 | 0.133 | 0.992 |
| FA | SS | -0.078 | 0.062 | 0.206 | -0.056 | 0.134 | 0.683 |
| FA | AverageFA | -0.076 | 0.064 | 0.236 | -0.001 | 0.133 | 0.997 |
| MD | ACR | 0.016 | 0.137 | 0.798 | 0.121 | 0.134 | 0.373 |
| MD | BCC | -0.090 | 0.032 | 0.149 | 0.186 | 0.135 | 0.176 |
| MD | CC | -0.086 | 0.036 | 0.168 | 0.101 | 0.134 | 0.458 |
| MD | CGC | 0.023 | 0.145 | 0.707 | 0.254 | 0.134 | 0.063 |
| MD | CR | 0.012 | 0.133 | 0.847 | 0.153 | 0.134 | 0.263 |
| MD | EC | -0.021 | 0.101 | 0.737 | 0.070 | 0.134 | 0.608 |
| MD | FX | -0.133 | -0.011 | 0.033 | -0.098 | 0.134 | 0.472 |
| MD | GCC | -0.035 | 0.086 | 0.573 | 0.132 | 0.134 | 0.331 |
| MD | PCR | -0.001 | 0.120 | 0.984 | 0.183 | 0.134 | 0.179 |
| MD | PTR | 0.022 | 0.144 | 0.720 | 0.218 | 0.134 | 0.109 |
| MD | SCR | 0.023 | 0.144 | 0.716 | 0.169 | 0.134 | 0.214 |
| MD | SLF | -0.009 | 0.113 | 0.884 | 0.169 | 0.134 | 0.216 |
| MD | SS | -0.023 | 0.099 | 0.711 | 0.143 | 0.134 | 0.293 |
| MD | UNC | -0.042 | 0.080 | 0.505 | 0.334 | 0.139 | 0.018 |
| RD | ACR | 0.050 | 0.064 | 0.440 | 0.095 | 0.134 | 0.486 |
| RD | BCC | -0.071 | 0.064 | 0.271 | 0.056 | 0.134 | 0.684 |
| RD | CC | -0.055 | 0.064 | 0.395 | 0.097 | 0.135 | 0.479 |
| RD | CR | 0.062 | 0.064 | 0.334 | 0.181 | 0.134 | 0.184 |

| | | | | | | | |
|---|---|---|---|---|---|---|---|
| RD | EC | 0.025 | 0.064 | 0.699 | 0.028 | 0.135 | 0.838 |
| RD | FX | -0.119 | 0.064 | 0.065 | -0.144 | 0.134 | 0.289 |
| RD | FXST | 0.110 | 0.064 | 0.087 | 0.194 | 0.134 | 0.154 |
| RD | GCC | 0.005 | 0.064 | 0.937 | 0.042 | 0.134 | 0.758 |
| RD | IC | 0.035 | 0.064 | 0.591 | 0.122 | 0.134 | 0.370 |
| RD | PCR | 0.063 | 0.064 | 0.329 | 0.149 | 0.134 | 0.275 |
| RD | PTR | 0.087 | 0.064 | 0.178 | 0.142 | 0.134 | 0.298 |
| RD | SCR | 0.079 | 0.064 | 0.217 | 0.091 | 0.134 | 0.505 |
| RD | SLF | 0.033 | 0.064 | 0.610 | 0.182 | 0.134 | 0.182 |
| RD | SS | 0.069 | 0.064 | 0.284 | 0.135 | 0.134 | 0.321 |
| RD | UNC | 0.018 | 0.064 | 0.786 | 0.223 | 0.138 | 0.112 |
| RD | AverageRD | 0.033 | 0.064 | 0.610 | 0.143 | 0.134 | 0.294 |
